# Genomics yields biological and phenotypic insights into bipolar disorder

**DOI:** 10.1101/2023.10.07.23296687

**Authors:** Kevin S. O’Connell, Maria Koromina, Tracey van der Veen, Toni Boltz, Friederike S. David, Jessica Mei Kay Yang, Keng-Han Lin, Xin Wang, Jonathan R. I. Coleman, Brittany L. Mitchell, Caroline C. McGrouther, Aaditya V. Rangan, Penelope A. Lind, Elise Koch, Arvid Harder, Nadine Parker, Jaroslav Bendl, Kristina Adorjan, Esben Agerbo, Diego Albani, Silvia Alemany, Ney Alliey-Rodriguez, Thomas D. Als, Till F. M. Andlauer, Anastasia Antoniou, Helga Ask, Nicholas Bass, Michael Bauer, Eva C. Beins, Tim B. Bigdeli, Carsten Bøcker Pedersen, Marco P. Boks, Sigrid Børte, Rosa Bosch, Murielle Brum, Ben M. Brumpton, Nathalie Brunkhorst-Kanaan, Monika Budde, Jonas Bybjerg-Grauholm, William Byerley, Judit Cabana-Domínguez, Murray J. Cairns, Bernardo Carpiniello, Miquel Casas, Pablo Cervantes, Chris Chatzinakos, Hsi-Chung Chen, Tereza Clarence, Toni-Kim Clarke, Isabelle Claus, Brandon Coombes, Elizabeth C. Corfield, Cristiana Cruceanu, Alfredo Cuellar-Barboza, Piotr M. Czerski, Konstantinos Dafnas, Anders M. Dale, Nina Dalkner, Franziska Degenhardt, J. Raymond DePaulo, Srdjan Djurovic, Ole Kristian Drange, Valentina Escott-Price, Ayman H. Fanous, Frederike T. Fellendorf, I. Nicol Ferrier, Liz Forty, Josef Frank, Oleksandr Frei, Nelson B. Freimer, John F. Fullard, Julie Garnham, Ian R. Gizer, Scott D. Gordon, Katherine Gordon-Smith, Tiffany A. Greenwood, Jakob Grove, José Guzman-Parra, Tae Hyon Ha, Tim Hahn, Magnus Haraldsson, Martin Hautzinger, Alexandra Havdahl, Urs Heilbronner, Dennis Hellgren, Stefan Herms, Ian B. Hickie, Per Hoffmann, Peter A. Holmans, Ming-Chyi Huang, Masashi Ikeda, Stéphane Jamain, Jessica S. Johnson, Lina Jonsson, Janos L. Kalman, Yoichiro Kamatani, James L. Kennedy, Euitae Kim, Jaeyoung Kim, Sarah Kittel-Schneider, James A. Knowles, Manolis Kogevinas, Thorsten M. Kranz, Kristi Krebs, Steven A. Kushner, Catharina Lavebratt, Jacob Lawrence, Markus Leber, Heon-Jeong Lee, Calwing Liao, Susanne Lucae, Martin Lundberg, Donald J. MacIntyre, Wolfgang Maier, Adam X. Maihofer, Dolores Malaspina, Mirko Manchia, Eirini Maratou, Lina Martinsson, Manuel Mattheisen, Nathaniel W. McGregor, Melvin G. McInnis, James D. McKay, Helena Medeiros, Andreas Meyer-Lindenberg, Vincent Millischer, Derek W. Morris, Paraskevi Moutsatsou, Thomas W. Mühleisen, Claire O’Donovan, Catherine M. Olsen, Georgia Panagiotaropoulou, Sergi Papiol, Antonio F. Pardiñas, Hye Youn Park, Amy Perry, Andrea Pfennig, Claudia Pisanu, James B. Potash, Digby Quested, Mark H. Rapaport, Eline J. Regeer, John P. Rice, Margarita Rivera, Eva C. Schulte, Fanny Senner, Alexey Shadrin, Paul D. Shilling, Engilbert Sigurdsson, Lisa Sindermann, Lea Sirignano, Dan Siskind, Claire Slaney, Laura G. Sloofman, Olav B. Smeland, Daniel J. Smith, Janet L. Sobell, Maria Soler Artigas, Dan J. Stein, Frederike Stein, Mei-Hsin Su, Heejong Sung, Beata Świątkowska, Chikashi Terao, Markos Tesfaye, Martin Tesli, Thorgeir E. Thorgeirsson, Jackson G. Thorp, Claudio Toma, Leonardo Tondo, Paul A. Tooney, Shih-Jen Tsai, Evangelia Eirini Tsermpini, Marquis P. Vawter, Helmut Vedder, Annabel Vreeker, James T. R. Walters, Bendik S. Winsvold, Stephanie H. Witt, Hong-Hee Won, Robert Ye, Allan H. Young, Peter P. Zandi, Lea Zillich, 23andMe Research Team, Estonian Biobank research team, Genoplan Research Team, HUNT All-In Psychiatry, PGC-FG Single cell working group, Genomic Psychiatry Cohort (GPC) Investigators, Rolf Adolfsson, Martin Alda, Lars Alfredsson, Lena Backlund, Bernhard T. Baune, Frank Bellivier, Susanne Bengesser, Wade H. Berrettini, Joanna M. Biernacka, Michael Boehnke, Anders D. Børglum, Gerome Breen, Vaughan J. Carr, Stanley Catts, Sven Cichon, Aiden Corvin, Nicholas Craddock, Udo Dannlowski, Dimitris Dikeos, Bruno Etain, Panagiotis Ferentinos, Mark Frye, Janice M. Fullerton, Micha Gawlik, Elliot S. Gershon, Fernando S. Goes, Melissa J. Green, Maria Grigoroiu-Serbanescu, Joanna Hauser, Frans A. Henskens, Jens Hjerling-Leffler, David M. Hougaard, Kristian Hveem, Nakao Iwata, Ian Jones, Lisa A. Jones, René S. Kahn, John R. Kelsoe, Tilo Kircher, George Kirov, Po-Hsiu Kuo, Mikael Landén, Marion Leboyer, Qingqin S. Li, Jolanta Lissowska, Christine Lochner, Carmel Loughland, Jurjen J. Luykx, Nicholas G. Martin, Carol A. Mathews, Fermin Mayoral, Susan L. McElroy, Andrew M. McIntosh, Francis J. McMahon, Sarah E. Medland, Ingrid Melle, Lili Milani, Philip B. Mitchell, Gunnar Morken, Ole Mors, Preben Bo Mortensen, Bertram Müller-Myhsok, Richard M. Myers, Woojae Myung, Benjamin M. Neale, Caroline M. Nievergelt, Merete Nordentoft, Markus M. Nöthen, John I. Nurnberger, Michael C. O’Donovan, Ketil J. Oedegaard, Tomas Olsson, Michael J. Owen, Sara A. Paciga, Christos Pantelis, Carlos N. Pato, Michele T. Pato, George P. Patrinos, Joanna M. Pawlak, Josep Antoni Ramos-Quiroga, Andreas Reif, Eva Z. Reininghaus, Marta Ribasés, Marcella Rietschel, Stephan Ripke, Guy A. Rouleau, Panos Roussos, Takeo Saito, Ulrich Schall, Martin Schalling, Peter R. Schofield, Thomas G. Schulze, Laura J. Scott, Rodney J. Scott, Alessandro Serretti, Jordan W. Smoller, Alessio Squassina, Eli A. Stahl, Hreinn Stefansson, Kari Stefansson, Eystein Stordal, Fabian Streit, Patrick F. Sullivan, Gustavo Turecki, Arne E. Vaaler, Eduard Vieta, John B. Vincent, Irwin D. Waldman, Cynthia S. Weickert, Thomas W. Weickert, Thomas Werge, David C. Whiteman, John-Anker Zwart, Howard J. Edenberg, Andrew McQuillin, Andreas J. Forstner, Niamh Mullins, Arianna Di Florio, Roel A. Ophoff, Ole A. Andreassen, the Bipolar Disorder Working Group of the Psychiatric Genomics Consortium

## Abstract

Bipolar disorder (BD) is a leading contributor to the global burden of disease^1^. Despite high heritability (60-80%), the majority of the underlying genetic determinants remain unknown^2^. We analysed data from participants of European, East Asian, African American and Latino ancestries (n=158,036 BD cases, 2.8 million controls), combining Clinical, Community, and Self-reported samples. We identified 298 genome-wide significant loci in the multi-ancestry meta-analysis, a 4-fold increase over previous findings^3^, and identified a novel ancestry-specific association in the East Asian cohort. Integrating results from fine-mapping and other variant-to-gene mapping approaches identified 36 credible genes in the aetiology of BD. Genes prioritised through fine-mapping were enriched for ultra-rare damaging missense and protein-truncating variations in BD cases^4^, highlighting convergence of common and rare variant signals. We report differences in genetic architecture of BD depending on the source of patient ascertainment and on BD-subtype (BDI and BDII). Several analyses implicate specific cell types in BD pathophysiology, including GABAergic interneurons and medium spiny neurons. Together, these analyses provide novel insights into the genetic architecture and biological underpinnings of BD.

## Main

Bipolar disorder (BD) is an often lifelong mood disorder that impairs quality of life, functional ability, and is associated with suicidality.^5^ Symptoms typically occur in early adulthood,^5^ with a similar prevalence and incidence rate across the world.^6^ Current treatment options include pharmacotherapies such as mood stabilisers, antipsychotics and antidepressants, preferably administered in conjunction with psychosocial interventions.^1,5^ However, approximately one third of patients relapse within the first year of treatment.^7^

The heterogeneous nature of the disorder is noted in the Diagnostic and Statistical Manual of Mental Disorders, fifth edition (DSM-5), which includes the category “bipolar and related disorders,” encompassing bipolar disorder type I (BDI), bipolar disorder type II (BDII) and cyclothymic disorders.^8^ The International Classification of Diseases, 11th Revision (ICD-11) also recognises BDI and BDII as distinct subtypes.^9^ BDI is characterised by episodes of both mania and depression, while BDII includes episodes of hypomania and depression. Advances in genetics and neuroimaging have begun to make inroads into the underlying pathophysiology of BD. The Psychiatric Genomics Consortium (PGC) Bipolar Disorder Working Group has spearheaded genetic discoveries in BD.^10,11^ A genome-wide association study (GWAS) of 41,917 BD cases and 371,549 controls identified 64 loci and highlighted calcium channel antagonists as potential targets for drug repurposing.^3^ Brain imaging studies have shown decreased cortical thickness, lower subcortical volume and disrupted white matter integrity associated with BD, as well as brain alterations associated with medication use.^12^ To date, this research has been conducted almost exclusively on individuals of European (EUR) ancestry.

Here, we present the largest to date multi-ancestry GWAS meta-analysis of 158,036 BD cases and 2,796,499 controls, combining Clinical, Community, and Self-reported samples. We identified 337 linkage disequilibrium (LD) independent genome-wide significant (GWS) variants that map to 298 loci. We hypothesised that differences in source of patient ascertainment, BD subtype, and genetic ancestry might lead to differences in genetic architecture, thus we also analysed these groups separately. We provide new insights into the genetic architecture and neurobiological mechanisms involved in BD, with the potential to inform the development of new treatments and precision medicine approaches.

### Study population

The current GWAS meta-analysis includes 79 cohorts. Case definitions were based on a range of assessment methods: (semi-)structured clinical interviews (Clinical), medical records, registries and questionnaire data (Community), and self-reported surveys (Self-reported). Details of the cohorts, including sample size, ancestry, and inclusion/exclusion criteria for cases and controls, are provided in Supplementary Tables 1 and 2 and the Supplementary Note. BD subtype data were available for a subset of individuals within the Clinical and Community groups. 82.5% of cases in the Clinical ascertainment group had BDI as did 68.7% of cases in the Community ascertainment group (X^2^=730, p < 2.2 ×▫10^−16^; Supplementary Table 2). The total number of samples available for analyses included 158,036 BD cases and 2,796,499 controls (effective n (Neff) =▫535,720; see Methods).

### Genetic architecture of BD differs by ascertainment and subtype

Given our hypothesis that samples ascertained and assessed by different methods could lead to differences in the genetic architecture, we performed meta-analyses separately for Clinical, Community and Self-reported samples. Using LDSC^13^ and assuming a population prevalence of 2%,^14^ BD ascertained from Clinical samples was more heritable (h^2^_SNP_ = 0.22; s.e. = 0.01) than BD ascertained from Community samples (h^2^_SNP_ = 0.05; s.e. = 0.003) or Self-report (h^2^_SNP_ = 0.08; s.e. = 0.003) (Supplementary Table 3). We used genetic correlation^13^ and MiXeR^15,16^ analyses to further investigate the genetic architecture of BD based on assessment. While there was a strong genetic correlation between Clinical and Community samples (*r*_*g*_ = 0.95; s.e. = 0.03), the genetic correlation for Self-reported BD was significantly greater (p = 7.4 ×□**1**0^−28^) with Community samples (*r*_*g*_ = 0.79; s.e. = 0.02) than with Clinical samples (*r*_*g*_ = 0.47; s.e. = 0.02) (Supplementary Figure 2).

MiXeR estimated the greatest polygenicity for BD ascertained from Self-reported samples, followed by Clinical and then Community samples (Figure 1, Supplementary Table 4). Almost all variants estimated to influence BD in Community samples were shared with BD ascertained from Clinical samples. The majority of Clinical and Community BD-influencing variants were also shared with Self-reported BD (Figure 1, Supplementary Figure 4). The mean correlation of variant effects in the shared components was high across all groups (Community and Self-reported r_g_shared_ = 0.95 (s.e. = 0.03), Community and Clinical r_g_shared_ = 0.99 (s.e. = 0.01) and Clinical and Self-reported r_g_shared_ = 0.74 (s.e. = 0.06) (Supplementary Table 4), supporting our decision to meta-analyse the three types of data sources.

**Figure 1.**
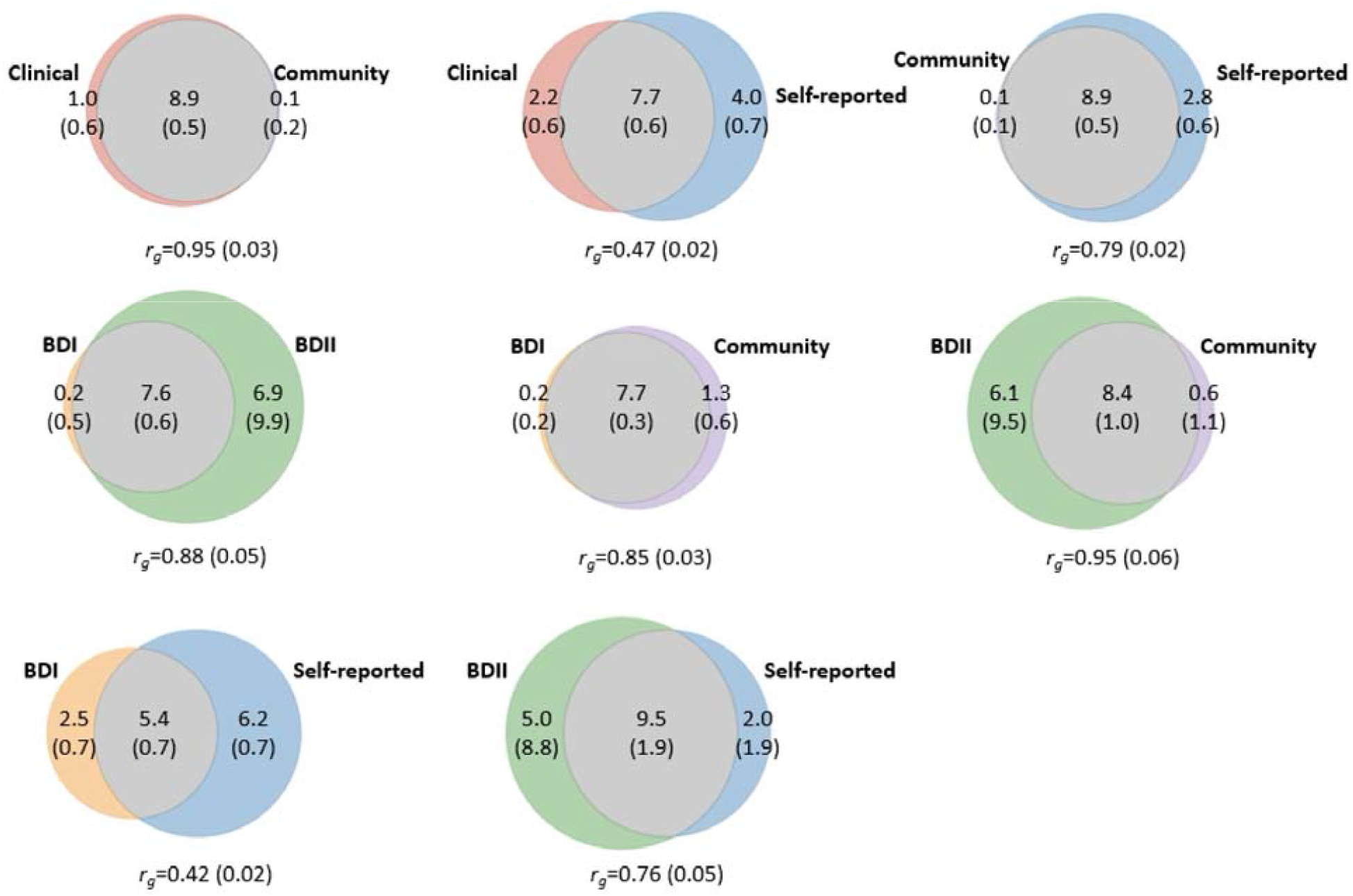
Genetic correlation and bivariate MiXeR estimates for the genetic overlap of BD ascertainment and subtypes. Trait-influencing genetic variants shared between each pair (grey) and unique to each trait (colours) are shown. The numbers within the Venn diagrams indicate the estimated number of trait-influencing variants (and standard errors) (in thousands) that explain 90% of SNP heritability in each phenotype. The size of the circles reflects the polygenicity of each trait, with larger circles corresponding to greater polygenicity. The estimated genetic correlation (rg) and standard error between BD and each trait of interest from LDSC is shown below the corresponding Venn diagram. Clinical and Community samples were stratified into bipolar I disorder (BDI) and bipolar II disorder (BDII) subtypes if subtype data were available. Model fit statistics indicated that MiXeR-modelled overlap for bivariate comparisons including the bipolar subtypes (BDI and BDII) were not distinguishable from minimal or maximal possible overlap, and therefore to be interpreted with caution (see Supplementary Table 4).

To analyse BD subtypes, we used available GWAS summary statistics for BDI (25,060 cases) and BDII (6,781 cases)^3^, which come from a subset of the Clinical and Community samples. Assuming a population prevalence of 1%,^17^ BDI was more heritable (h^2^_SNP_ = 0.21; s.e. = 0.01) than BDII (h^2^_SNP_ = 0.11; s.e. = 0.01). BDI and BDII were highly, but imperfectly, correlated (*r*_*g*_ = 0.88; s.e. = 0.05). The genetic correlations between both subtypes and the Community samples were high (BDI *r*_*g*_ = 0.85; s.e. = 0.03, BDII *r*_*g*_ = 0.95; s.e. = 0.06). In contrast, the genetic correlation between BDI and Self-reported BD (*r*_*g*_ = 0.42; s.e. = 0.02) was significantly lower (p = 7.1 ×□10^−13^) than between BDII and Self-reported BD (*r*_*g*_ = 0.76; s.e. = 0.05) (Supplementary Figure 2).

Given the difference in proportion of BDI and BDII cases within the Clinical and Community cohorts, we evaluated the genetic correlation between BD within Clinical and Community cohorts, and Self-reported BD, after conditioning on the genetic risk for BDI and BDII. After conditioning, the genetic correlation between Self-reported BD and BD within Community cohorts (*r*_*g*_ = 0.92; s.e. = 0.09) = was not significantly different (p = 0.10) than with BD in Clinical cohorts (*r*_*g*_ = 0.71; s.e. = 0.13).

### Ancestry-specific GWAS meta-analyses

We conducted separate meta-analyses in four ancestral groups. Because the self-reported data differed in genetic architecture from the clinical and community data, we performed separate meta-analyses with and without the inclusion of the self-reported data. Supplementary Table 2 provides a summary of the GWAS meta-analyses and details of associated loci are described in Supplementary Tables 5-7. Ancestry-specific estimates of SNP-heritability and cross-ancestry genetic correlations are provided in Supplementary Table 3.

We identified 261 independent GWS variants mapping to 221 loci associated with BD in EUR ancestry meta-analyses that included self-reported data, and 94 independent GWS variants mapping to 88 loci without self-reported data (Supplementary Tables 5 and 6). There were 92 of the 94 independent GWS variants available for meta-analysis in the Self-reported cohorts, of which 78 (85%) were concordant for direction of effect (Supplementary Table 6).

In the East Asian (EAS) ancestry meta-analysis we identified two BD-associated loci, one of which is novel with an ancestry-specific index variant (rs117130410, 4:105734758, build GRCh37; Supplementary Figure 1, Supplementary Table 7). While this variant had a frequency of 16% and 9% in EAS BD cases and controls, respectively, it is monomorphic in non-Asian populations. The second locus (rs174576, 11:61603510, build GRCh37; Supplementary Table 7) was only identified when the self-reported data were excluded from the meta-analysis since the index variant was not available in the self-reported data. This locus has been identified previously and implicates the *FADS1* and *FADS2* genes.^3,18^ No GWS loci were observed in the African American (AFR) or Latino (LAT) ancestry meta-analyses.

### Multi-ancestry meta-analysis

A multi-ancestry meta-analysis of all of the datasets identified 337 LD independent GWS variants mapping to 298 loci (Supplementary Figure 1, Supplementary Table 8). There was minimal test statistic inflation due to uncontrolled population stratification after correction for principal components in each dataset (LDSC intercept = 1.052 (se = 0.016), attenuation ratio=0.071 (s.e. = 0.013)).

Of the 298 loci identified in this multi-ancestry meta-analysis, 267 are novel for BD. Of the 64 previously reported BD-associated loci,^3^ 31 met GWS in the present analysis containing all samples, and of the 33 that did not, 25 met GWS in either the Clinical samples or in the meta-analysis that excluded Self-reported data (Supplementary Table 9). Moreover, the direction of association for all top SNPs (12,151 SNPs with p < 1 ×□10^−5^) from the previous GWAS was consistent with the direction of association in this multi-ancestry meta-analysis of all samples (Supplementary Table 9).

When considering the impact of ancestry on the discovery of these 298 loci, one locus (index SNP rs7248481, chr19:13079957-13122567) was most strongly associated in the EAS ancestry meta-analysis. For all other loci, the association was strongest in the EUR ancestry meta-analysis. The majority of the 298 loci were nominally significant (p < 0.05) within the AFR (290/298 loci), EAS (257/298 loci) and LAT (293/298 loci) ancestry-specific meta-analyses, highlighting consistency of signal across the ancestry groups (Supplementary Table 8).

We used GSA-MiXeR^19^ to estimate the proportion of SNP-heritability (SNP-h^2^) accounted for by SNPs within GWS loci. Compared to only 8.3% accounted for by SNPs within the 64 previously identified loci,^3^ SNPs within the 298 loci account for 18.5% of the SNP-h^2^ of BD (Supplementary Table 10). Moreover, SNPs within the 298 loci also accounted for higher proportions of SNP-h2 in the Clinical (64 loci: 8.5%; 298 loci: 17.8%), BDI (64 loci: 8.3%; 298 loci: 17.5%), Community (64 loci: 4.8%; 298 loci: 22.6%), and Self-reported (64 loci: 2.0%; 298 loci: 21.1%) samples.

We carried out sensitivity meta-analyses excluding the Self-reported samples (leaving 67,948 cases and 867,710 controls; N_eff_ = 191,722), and identified 116 independent GWS variants mapping to 105 loci (Supplementary Table 11). There was minimal test statistic inflation due to uncontrolled population stratification after correction for principal components in each dataset (LDSC intercept = 1.050; se = 0.012, attenuation ratio=0.086; s.e. = 0.018). Analysis of Self-report cohorts only (90,088 cases and 1,928,789 controls; N_eff_ = 344,088) identified 126 loci (Supplementary Table 12). Of the 116 independent GWS variants identified in the meta-analysis excluding the Self-report samples, 110 were available for meta-analysis in the Self-report samples, of which 96 (87%) were concordant (Supplementary Table 11).

### Genetic correlations with other traits

Genome-wide genetic correlations (*r*_*g*_) were estimated between EUR ancestry BD GWASs (with and without self-reported data, and when stratified by ascertainment and subtypes) and human diseases and traits via the Complex Traits Genetics Virtual Lab (CTG-VL; https://vl.genoma.io) web platform^20^ (Figure 2, Supplementary Tables 13-15). Most psychiatric disorders, including major depressive disorder (MDD), post-traumatic stress disorder (PTSD), attention deficit/hyperactivity disorder (ADHD), borderline personality disorder, and autism spectrum disorder (ASD), were more strongly correlated with the full meta-analysis, as well as with BDII, and BD in Community and Self-reported samples (Figure 2). In contrast, schizophrenia was more strongly genetically correlated with the full BD meta-analysis excluding self-reported data and with BDI and BD in clinical samples (Figure 2).

**Figure 2.**
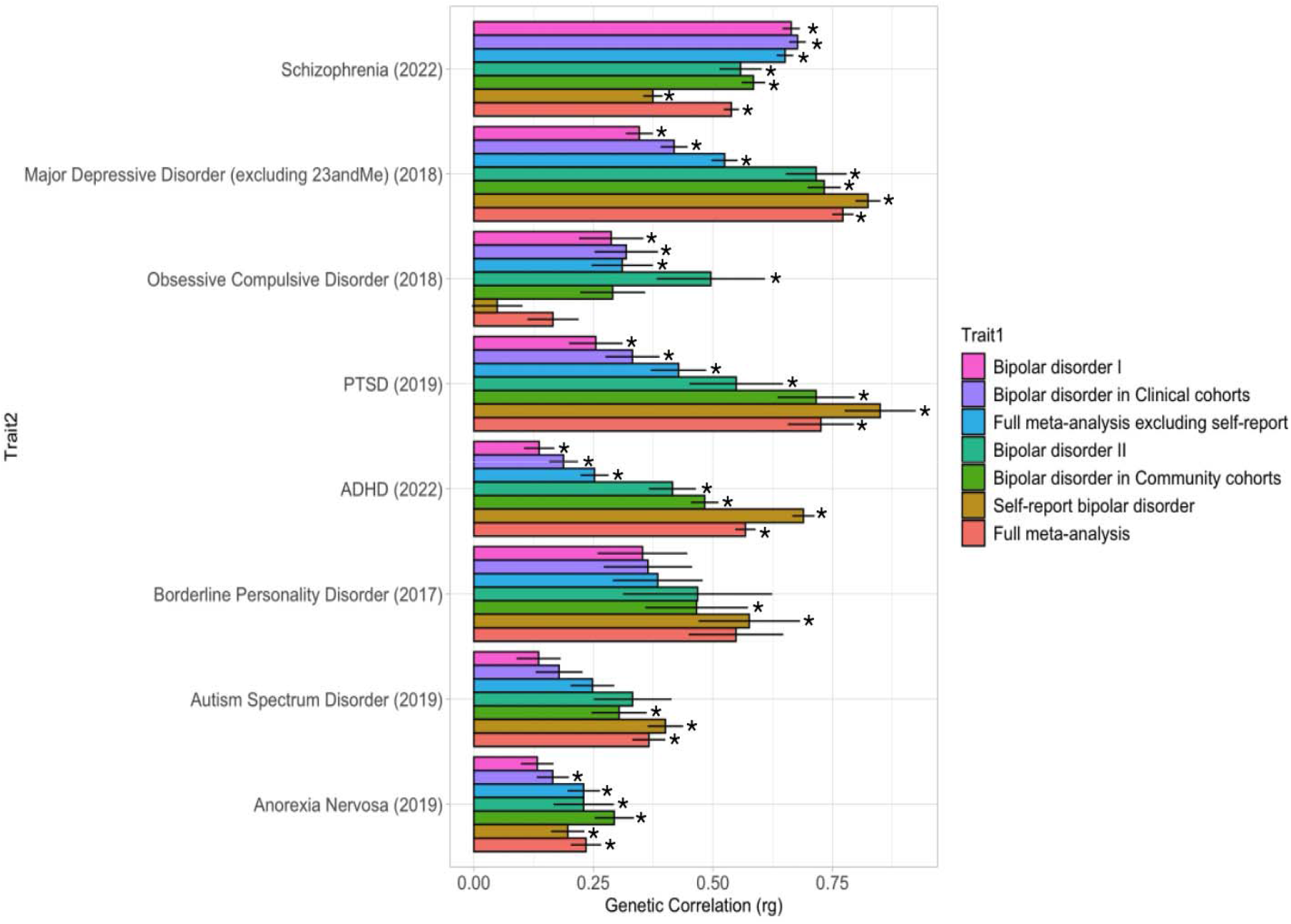
Genetic correlations between bipolar disorder and other psychiatric disorders. The y-axis (Trait2) is ordered based on the significance and magnitude of genetic correlation of each trait with bipolar disorder type I. Stars indicate results passing the Bonferroni corrected significance threshold of P < 3.6 ×□10^−5^. ADHD, attention deficit/hyperactivity disorder. PTSD, post-traumatic stress disorder. The year indicated in parentheses after each trait refers to the year in which the GWAS was published. Details are provided in Supplementary Table 13.

### Polygenic prediction of bipolar disorder

Polygenic risk score (PRS) analyses were performed using PRS-CS-auto^21^ in 55 EUR ancestry cohorts for which individual-level genotype and phenotype data were available (40,992 cases and 80,215 controls), as well as one cohort of AFR ancestry (347 cases and 669 controls) and three cohorts of EAS ancestry (4,473 cases and 65,923 controls) (Supplementary Tables 16-20). In the EUR ancestry cohorts, the variance explained by the multi-ancestry GWAS without the self-reported data (R^2^ = 0.090, s.e. = 0.019) was significantly greater than that explained by both the multi-ancestry GWAS including self-report data (R^2^ = 0.058, s.e. = 0.017, P = 2.72□×□10^−4^), and by the the EUR ancestry GWAS excluding the self-reported data (R^2^ = 0.084, s.e. = 0.018, P = 5.62 ×□10^−3^) (Figure 3A, Supplementary Tables 16 and 21). Individuals in the top quintile (top 20%) for this multi-ancestry GWAS without the self-reported data PRS had an odds ratio of 7.06 (95% confidence interval (CI) 3.9–10.4) of being affected with BD compared to individuals in the middle quintile. The corresponding median Area Under the Receiver Operating Characteristic Curve (AUC) was 0.70 (95% CI= 0.67-0.73). Therefore, the BD liability explained remains insufficient for diagnostic prediction in the general population.

**Figure 3.**
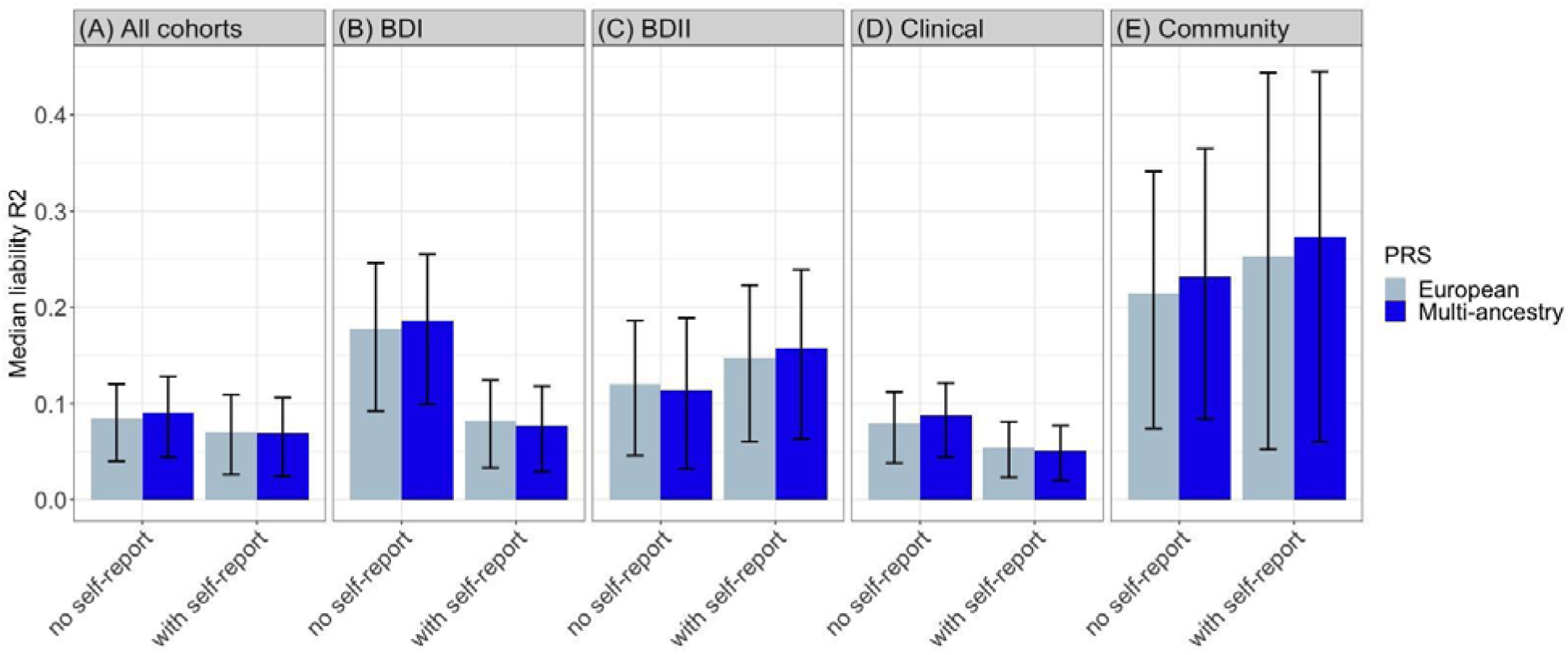
Phenotypic variance in bipolar disorder in European cohorts explained by polygenic risk scores derived from the multi-ancestry and European meta-analyses (with and without self-reported data).

Similarly, PRS derived from GWAS excluding self-reported data explained significantly more variance in cases of BDI (Figure 3B, Supplementary Tables 17) and in Clinical cohorts (Figure 3D, Supplementary Tables 19) than when self-reported data were included. Conversely, inclusion of the self-reported data yielded greater median R^2^ estimates for the PRS in cases of BDII (Figure 3C, Supplementary Tables 18) and in Community cohorts (Figure 3E, Supplementary Tables 20), although these differences were not significant.

Variance explained is presented on the liability scale, assuming a 2% population prevalence of bipolar disorder. The results in the first panel are the median weighted liability R^2^ values across all 55 European cohorts (40,992 cases, 80,215 controls, N_eff_ = 46,725). Similarly, the remaining panels show the results across 36 bipolar disorder I (BDI) cohorts (12,419 cases and 33,148 controls, N_eff_ = 14,607), 21 bipolar disorder II (BDII cohorts, 2,549 cases, 23,385 controls, N_eff_ = 4,021), 48 Clinical cohorts (27,833 cases, 46,623 controls, N_eff_ = 29,543), and 7 Community cohorts (13,159 cases, 36,592 controls, N_eff_ = 17,178). All analyses were weighted by the effective *n* per cohort. Error bars show 95% confidence intervals (CI). Comparison of variance explained was determined using 1- and 2-sample Wilcoxon tests (Supplementary Table 21)

PRS analysis of three clinically ascertained EAS cohorts revealed that the PRSs derived from GWAS excluding the self-reported data (Taiwan; EUR-PRS R2 = 0.069, Multi-PRS R2 = 0.075. Japan; EUR-PRS R2 = 0.027, Multi-PRS R2 = 0.025. Korea; EUR-PRS R2 = 0.016, Multi-PRS R2 = 0.022) performed better than those that included self-reported data (Taiwan; EUR-PRS R2 = 0.026, Multi-PRS R2 = 0.036. Japan; EUR-PRS R2 = 0.015, Multi-PRS R2 = 0.015. Korea; EUR-PRS R2 = 0.014, Multi-PRS R2 = 0.017) (Supplementary Table 22).

In a clinically ascertained AFR target cohort, the inclusion of self-reported data increased the explained variance by both the multi-ancestry PRS and the EUR ancestry PRS from 0.010 to 0.23 or 0.22, respectively (Supplementary Table 22).

### Pathway, tissue and cell type enrichment

Gene-set enrichment analyses were performed on the summary statistics derived from the multi-ancestry meta-analysis including self-reported data, using MAGMA.^22^ We identified significant enrichment of 6 gene-sets (Supplementary Table 23) related to the synapse and transcription factor activity. The association signal was enriched among genes expressed in the brain (Supplementary Table 24), and specifically in the early-to mid-prenatal stages of development (Supplementary Table 25). Single-cell enrichment analyses of brain cell types indicate involvement of neuronal populations from different brain regions, including hippocampal pyramidal neurons and interneurons of the prefrontal cortex and hippocampus (Supplementary Figure 5), and were largely consistent with findings from the previous PGC BD GWAS^3^. Similar patterns of enrichment were observed based on ascertainment and subtype (Supplementary Figure 6). In addition, GSA-MiXeR^19^ highlighted enrichment of specific dopamine- and calcium-related biological processes and molecular functions, as well as GABAergic interneuron development, respectively (Supplementary Table 26).

A recent study^23^ analysed single-nucleus RNA sequencing (snRNAseq) data of 3.369 million nuclei from 106 anatomical dissections within 10 brain regions and divided cells into 31 superclusters and 481 clusters, respectively, based on principal component analysis of sequenced genes. These superclusters were then annotated based on their regional composition within the brain (Figure 4). We used stratified LD score regression (S-LDSC)^24^ to estimate SNP-heritability enrichment for the top decile of expression proportion (TDEP) genes in each of the 31 superclusters and 481 clusters, as described previously.^25^ Heritability was significantly enriched in 9 of the 31 superclusters (Figure 4), and 49 of the 481 clusters (Supplementary Figure 7). No enrichment was seen in non-neuronal clusters. Interestingly, two clusters within the medium spiny neurons, not observed at the supercluster level, are significantly enriched further supporting the involvement of striatal processes in BD.

**Figure 4.**
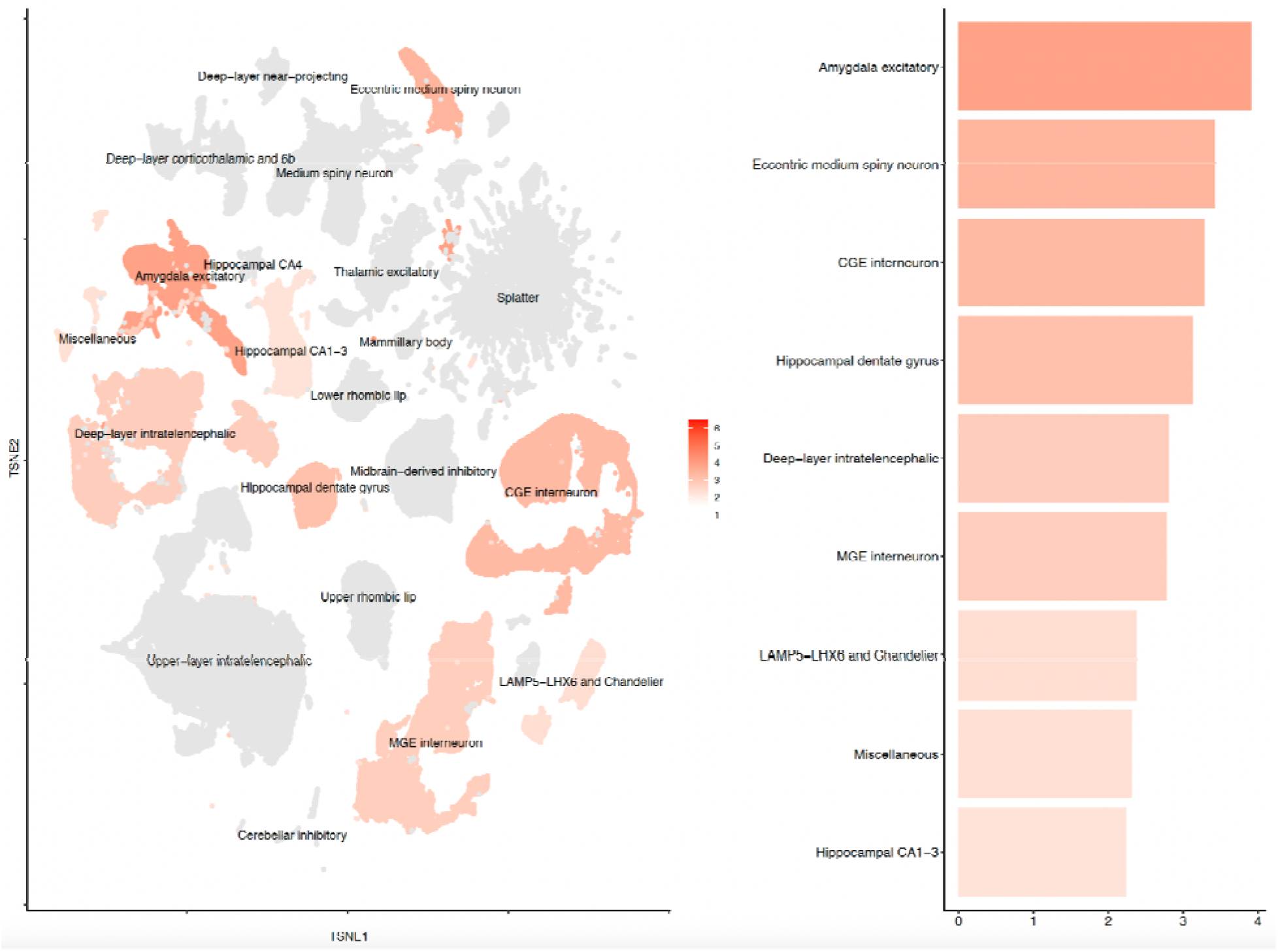
Supercluster-level SNP-heritability enrichment for bipolar disorder. The t-distributed stochastic neighbour embedding (tSNE) plot (from Siletti et al.^23^) (left) is coloured by the enrichment z-score. Grey indicates non-significantly enriched superclusters (FDR > 0.05). The barplot (right) shows the nine significantly enriched superclusters.

Single-cell enrichment analysis in 914 cell types across 29 non-brain murine tissues identified significant enrichment in the enteroendocrine cells of the large intestine and delta cells of the pancreas, which remained significant after cross-dataset conditional analyses with a murine brain tissue dataset (Supplementary Table 27).

### Fine-mapping

We performed functional fine-mapping using Polyfun+SuSiE (Supplementary Tables 28 and 29).^26^ At a threshold of PIP > 0.50, we identified 80 putatively causal fine-mapped SNPs for the multi-ancestry meta-analyses including self-reported data. At the more stringent threshold of PIP > 0.95 we identified 20 putatively causal SNPs. When comparing the number of SNPs within 95% credible sets, the inclusion of multi-ancestry and self-reported data led to smaller credible sets (i.e. credible sets with fewer numbers of SNPs). For example, we identified 175 95% credible sets of < 20 SNPs in the multi-ancestry dataset with self-reported data, compared to 122 in the European dataset with self-reported data (Supplementary Figure 8). Putatively causal SNPs with a PIP > 0.5 were mapped to genes by performing variant annotation with Variant Effect Predictor (VEP) (GRCh37) Ensembl release 109,^27^ based on their position relative to annotated Ensembl transcripts and known regulatory features. This analysis identified 71 unique genes annotated to fine-mapped SNPs from the multi-ancestry meta-analysis including self-reported data (Supplementary Table 29).

### Converging evidence of common and rare variation

Within loci associated with BD in the multi-ancestry meta-analysis, the 71 genes annotated to putatively causal fine-mapped SNPs (Supplementary Table 29) were enriched for ultra rare (<=5 minor allele count) damaging missense and protein-truncating variants in BD cases in the Bipolar Exome (BipEx) consortium dataset^4^ (Odds ratio (OR) = 1.16, 95% confidence interval (CI) = 1.05 -1.28, P = 0.002), and in schizophrenia cases in the Schizophrenia Exome Meta-analysis (SCHEMA) dataset^28^ (OR = 1.21, 95% CI = 1.02 - 1.43, P = 0.024). This enrichment is similar to that observed for schizophrenia^28^ and ADHD.^29^

### Identification of credible BD-associated genes

In addition to the 71 genes annotated to the fine-mapped putatively causal SNPs as described above, we annotated a further 45 genes to the 80 fine-mapped SNPs by SMR using eQTL and sQTL data, as well as by proximity, i.e. the nearest gene to each SNP (Supplementary Figure 9, Supplementary Tables 30 and 31). No genes were annotated to the CpGs identified by the mQTL analysis (Supplementary Table 30). We then determined if any of these 116 genes were also identified through the genome-wide gene-based analysis using MAGMA,^22^ eQTL analyses using TWAS as implemented in FUSION^30^ and isoTWAS,^31^ or through enhancer-promoter (E-P) interactions.^32,33^ This resulted in seven possible approaches by which loci could be mapped to genes including, eQTL evidence (eQTL or TWAS or FOCUS or isoTWAS), mQTL, sQTL, VEP, proximity, MAGMA and E-P interactions.

We integrated the results from the post-GWAS analyses described above and identified a credible set of 36 genes identified by at least three of the described approaches (Supplementary Table 31). The *SP4* gene was identified by six of these approaches, and astrocyte and GABAergic neuron specific regulation of *SP4*, by the GWS variant rs2107448, were identified from cell-type specific enhancer-promoter interaction results (Supplementary Table 31). Moreover, the *TTC12* and *MED24* genes were identified by five of the approaches. Eight of the 36 credible genes have synaptic annotations in the SynGO database.^34^ Three genes (*HTT, ERBB4* and *LR5NF*) were mapped to both postsynaptic and presynaptic compartments. One gene (*CACNA1B*) was mapped to only the presynapse and four genes (*SHANK2, OLFM1, SHISA9* and *SORCS3*) were mapped to only the postsynapse (Supplementary Table 32).

Based on the lifespan gene expression data from the Human Brain Transcriptome project (www.hbatlas.org),^35^ we identified two clusters of credible genes with distinct peaks in temporal expression (Supplementary Figure 10, Supplementary Table 31). The first cluster shows reduced prenatal gene expression, with gene expression peaking at birth and remaining stable over the life-course. Conversely, the second cluster shows a peak in gene expression during fetal development with a drop-off in expression before birth.

### Drug target analyses

Gene-set analyses were performed restricted to genes targeted by drugs, assessing individual drugs and grouping drugs with similar actions as described previously.^3,36^ Gene-level and gene-set analyses of the multi-ancestry GWAS summary statistics including self-report data were performed in MAGMA,^22^ and identified significant enrichment in the targets of anticonvulsant pregabalin (Supplementary Table 33). There was also significant enrichment in the targets of antipsychotics and anxiolytics (Supplementary Table 34).

Examination of the Drug Gene Interaction Database (DGIdb)^37^ to identify drug-gene interactions using the credible genes as input genes, showed that 15 out of 36 genes were interacting with a total number of 528 drugs. Gene-set enrichment analysis of these drug-gene interactions showed a significant enrichment (p<0.0001) for targets of the atypical antipsychotic drugs nemonapride and risperidone (Supplementary Table 35). However, after correction for the total number of drugs (N=69,018), the enrichment was non-significant (FDR>0.05). In addition, 16 of the 36 credible genes had evidence of tractability with a small molecule in the OpenTargets dataset, including *FURIN, MED24, THRA, ALDH2, ANKK1, ARHGAP15, CACNA1B, ERBB4, ESR1, FES, GPR139, HTT, MLEC, MSH6, PSMD14*, and *TOMM2*.

Among the 36 credible genes, two (*ALDH2* and *ESR1*) were within the list of 139 lithium target and interaction partner genes. The results of the network-based separation (S_AB_) analysis do not indicate a general overlap between the credible genes and lithium target genes in the human protein interactome (S_AB_=0.124, z-score=1.710, p-value=0.044). The positive S_AB_ value indicates that the lithium target genes and the 36 credible genes are separated from each other in the network of protein-protein interactions.

## Discussion

We performed the largest GWAS of BD, including diverse samples of EUR, EAS, AFR and LAT ancestry, resulting in an over four-fold increase in the number of BD-associated loci: 337 LD independent GWS variants mapping to 298 loci. In the meta-analysis of EUR, the largest ancestry group, we identified over 200 GWS loci. We also found a novel ancestral-specific association in the EAS cohort. We confirmed our hypothesis that differences in ascertainment and BD subtype might lead to differences in genetic architecture. Post-GWAS analyses provide novel insights into the biological underpinnings and genetic architecture of BD and highlight differences depending on ascertainment of participants and BD-subtype. We also showed that multi-ancestry data improved fine-mapping and polygenic prediction.

Enrichment of the common variant associations from this multi-ancestry meta-analysis highlights the synapse, interneurons of the prefrontal cortex and hippocampus, and hippocampal pyramidal neurons as particularly relevant. Exploratory analyses using GSA-MiXeR^19^ suggest enrichment of dopamine- and calcium-related biological processes and development of GABAergic interneurons. These findings were further corroborated by enrichment analyses in single-nucleus RNA-seq data from adult postmortem brain tissue, which highlighted specific clusters of interneurons derived from the caudal and medial ganglionic eminences and medium spiny neurons predominantly localised in the striatum. Medium spiny neurons are not enriched in depression using the same dataset.^25^ Although interneurons derived from ganglionic eminences were also enriched in schizophrenia, stronger signals were observed for amygdala excitatory and hippocampal neurons.^25^

A novel finding is that single-cell enrichment analysis of non-brain murine tissues identified significant enrichment in the enteroendocrine cells of the large intestine and delta cells of the pancreas. Conditional analyses suggest that this enrichment is independent of overlapping genes between these cell-types and those expressed in neurons. Stimulation of enteroendocrine cells by short-chain fatty acids (SCFAs) promotes serotonin production in the colon which leads to enhanced levels of serotonin in systemic circulation and in the brain, and is a proposed mechanism by which microbiota influence the gut-brain axis.^38,39^ Notably, lithium treatment is shown to upregulate SCFA-producing bacteria highlighting a potential mechanism of action.^40^

We mapped genes to the 80 putatively causal SNPs identified from fine-mapping based on seven complementary approaches and identified a subset of 36 credible genes implicated by at least three of these approaches. The top credible gene, identified by six gene-mapping approaches, was *SP4*, which has also been implicated in schizophrenia through both rare^28^ and common variation.^41^ Moreover, we clustered the credible genes based on similar patterns of temporal variation in expression over the lifespan and identified two clusters. The first cluster shows reduced gene expression pre-birth, with gene expression peaking at birth and remaining stable over the life-course, while the second cluster shows a peak in gene expression during fetal development aligning with the neurodevelopmental hypothesis of mental disorders.^42^ Genes prioritised through fine-mapping were shown to be enriched for ultra rare damaging missense and protein-truncating variation in the BipEx^4^ and SCHEMA^28^ datasets, respectively, highlighting convergence of common and rare variant signals as recently shown in schizophrenia.^41^

We identified differences in the genetic architecture of BD subtypes related to ascertainment. BD within Clinical and Community samples was highly but imperfectly correlated, with varying correlations with Self-reported BD. The low genetic correlation and minimal genetic overlap between cases ascertained through clinical studies and cases with self-reported BD is driven by a greater proportion of BDI within the Clinical and Community samples. In line with these results, PRS derived from meta-analyses excluding the self-reported data performed better in Clinical and BDI target samples, while the inclusion of self-reported data improved the PRS in Community and BDII target samples. Moreover, the pattern of correlations between BD and other psychiatric disorders differed with the inclusion of self-reported data. Schizophrenia had the highest genetic correlation with BD without the inclusion of the self-reported data, while major depressive disorder was most strongly correlated with BD after the inclusion of the self-reported data. These results suggest that the Self-reported samples may include a high proportion of people with BDII. Moreover, this is in line with recent findings in individuals diagnosed with BDII, which showed increasing polygenic scores for depression and ADHD and decreasing polygenic scores for BD over time.^43^ However, a diagnosis of BD in the outpatient setting may be overdiagnosed in people with conditions such as chronic depression or borderline personality disorder, highlighting a higher rate of comorbid disorders and potential for ‘overdiagnosis’ of BD within cohorts of this nature.^44,45^ We showed that the differences in genetic architecture and phenotypic proportions of the Clinical, Community and Self-reported BD cohorts impacted the replication of prior BD-associated loci. Previously associated loci that fell short of meeting GWS in the current study were GWS in the Clinical samples and in the meta-analyses that excluded Self-reported data, and all top SNPs (12,151 SNPs with p < 1 × 10−5) from the previous GWAS were consistent in direction of association in this multi-ancestry meta-analysis of all samples (Supplementary Table 9).

Investigation of the novel ancestral-specific association in the EAS ancestry meta-analysis in the GWAS catalog^46^ highlights overlaps with genome-wide significant loci for reduced sleep duration,^47^ and lower educational attainment,^48^ as well as a suggestive locus (p < 2 ×□10^−6^) for the interaction between cognitive function and MDD.^49^ These findings suggest a role for this genomic region in complex brain-related phenotypes.

The multi-ancestry PRS provided the greatest improvement over the EUR-PRS in two of the three EAS ancestry target cohorts (Korean and Taiwanese). More subtle improvements were seen when the EUR target cohorts were analysed. Multi-ancestry training data provided little improvement in the AFR target cohort, which may be due to the genetic heterogeneity of this target cohort.^50^ These results highlight the benefits of multi-ancestry representation in the PRS training data, in line with findings from other diseases.^51^ The predictive power of this BD PRS shows a substantial improvement compared to previous findings^3^ however, this BD PRS alone still falls short of clinical utility.^52^ The meta-analysis excluding self-reported data produced results with the most explanatory power; this is likely due to increased phenotypic heterogeneity when the self-reported data are included.

One limitation is the lack of in-sample LD estimates for all cohorts, due to a lack of in-house raw genotype data for some cohorts. For instance, analysis of the MHC/C4 locus was not considered since the number of samples for which individual-level genotype data were accessible did not increase much since the previous analysis^3^. We used a EUR LD reference panel to analyse the multi-ancestry meta-analyses^53^ where LD patterns and interindividual heterogeneity within the ancestry groups are not fully captured. Another limitation is the inclusion of samples with minimal phenotyping. Although this allowed us to achieve large sample sizes, especially in under-represented non-European ancestry cohorts, and greatly increase the number of loci identified, minimally-phenotyped samples have some shortcomings. For example, minimal phenotyping may result in low specificity association signals, as shown in major depression,^54,55^ and individuals in community-based biobanks may represent those less severely affected, as shown in schizophrenia.^56^

In conclusion, in this first large-scale multi-ancestry GWAS of BD, we identified 298 significant BD-associated loci, from which we demonstrate convergence of common variant associations with rare variant signals and highlight 36 genes credibly implicated in the pathobiology of the disorder. We identified differences in the genetic architecture of BD based on ascertainment and subtype, suggesting that stratification by subtype will be important in BD genetics moving forward. Several analyses implicate specific cell types in BD pathophysiology, including GABAergic interneurons and medium spiny neurons, as well as the enteroendocrine cells of the large intestine and delta cells of the pancreas. Enrichment of dopamine- and calcium-related biological processes were also identified, further contributing to our understanding of the biological aetiology of BD.

## Methods

### Sample description

Details of each of the cohorts, including sample size, ancestry, inclusion/exclusion criteria for cases and controls as well as citations, are provided in Supplementary Table 1 and the Supplementary Note. We included three types of samples: 1) samples where participants were assessed using semi-structured or structured interviews (Clinical), 2) samples where participants were assessed using medical records, registries and questionnaire data (Community) and 3) samples where participants self-report a diagnosis of bipolar disorder (Self-report). The Clinical samples included 55 cohorts, 46 of which were included in previous PGC-BD GWAS publications^3,10,11^. The Community samples included 20 cohorts, 11 of which were included in the previous PGC-BD GWAS^3^. Finally, we included four Self-report cohorts from 23andMe, Inc, in which individuals were classified as cases if they self-reported having received a clinical diagnosis or treatment for bipolar disorder in responses to web-based surveys (“Have you ever been diagnosed with, or treated for, bipolar disorder?”).

Individual-level genotype and phenotype data were shared with the PGC for 53 ‘internal’ cohorts, while the remaining 26 ‘external’ cohorts contributed summary statistics data.

The final multi-ancestry meta-analysis included up to 158,036 cases and 2,796,499 controls. The total effective n (N_eff_), equivalent to an equal number of cases and controls in each cohort (4□× □ncases□× □ncontrols/(ncases□+ □ncontrols) is 535,720 with 82.3% of participants (proportion of N_eff_) of EUR ancestry, 4.4% of AFR ancestry, 4.2% of EAS ancestry and 9.1% of LAT ancestry.

The majority of new cohorts included in this study were external community cohorts where subtype definitions were more difficult to determine, and as such the total number of BDI and BDII subtype cases does not differ remarkably from the previous PGC BD GWAS^3^ (Supplementary Table 1). Thus, the previous BDI (25,060 cases and 449,978 controls) and BDII (6,781 cases and 364,075 controls) GWAS summary statistics data were used for BDI and BDII analyses in this study.

### Genotyping, quality control and imputation

Technical quality control was performed separately on each cohort for which individual-level data were provided separately according to standards developed by the PGC^57^ including; SNP missingness□<□0.05 (before sample removal), subject missingness□< □0.02, autosomal heterozygosity deviation (Fhet□< □0.2), SNP missingness□<□0.02 (after sample removal), difference in SNP missingness between cases and controls□< □0.02, SNP Hardy–Weinberg equilibrium (P□> □1□× □10^−10^ in BD cases and P□> □1□× □10^−6^ in controls), and mismatches between pedigree and genetically-determined sex based on the F statistic of X chromosome homozygosity (female F < 0.2 and male F >0.8). In addition, relatedness was calculated across cohorts using identity by descent and one of each pair of related individuals (pi_hat□> □0.2) was excluded, prioritising exclusion of individuals related to the most others, controls over cases, and individuals from larger cohorts. Principal components (PCs) were generated using genotyped SNPs in each cohort separately using EIGENSTRAT v6.1.4^58^. Genotype imputation was performed using the prephasing/imputation stepwise approach implemented in Eagle v2.3.5^59^ and Minimac3^60^ to the Haplotype Reference Consortium (HRC) reference panel v1.0^61^. Data on the X chromosome were also available for all 53 internal cohorts and these were imputed to the HRC reference panel in males and females separately. The remaining 22 external cohorts were processed by the contributing collaborative teams using comparable procedures. Identical individuals between PGC processed cohorts and external cohorts with suspected sample overlap were detected using genotype-based checksums (https://personal.broadinstitute.org/sripke/share_links/zpXkV8INxUg9bayDpLToG4g58TMtjN_PGC_SCZ_w3.0718d.76) and removed from the PGC cohorts.

### Genome-wide association study (GWAS)

For internal cohorts, GWASs were conducted within each cohort using an additive logistic regression model in PLINK v1.90^62^, covarying for the first five PCs and any others as required, as previously described^3^. Analyses of the X chromosome were performed in males and females separately, with males scored 0 or 2 and females scored 0, 1 or 2. X chromosome analyses were performed only in individuals of EUR ancestry for which individual level data were available. For external cohorts, GWASs were conducted by the collaborating research teams using comparable procedures. To control test statistic inflation at SNPs with low minor allele frequency (MAF) in small cohorts, SNPs were retained only if cohort MAF was >1% and minor allele count was >10 in either cases or controls (whichever had smaller n).

Initially, meta-analysis of GWAS summary statistics was conducted using inverse-variance-weighted fixed-effects models in METAL (version 2011-03-25)^63^ across cohorts within ancestral groups. A GWS locus was defined as the region around a SNP with P < 5.0 × 10^−8^ with linkage disequilibrium (LD) r2 > 0.1, within a 3000 kb window, based on the LD structure of the ancestry matched HRC reference panel v1.0^61^, except LAT (EUR panel used). Multi-ancestry meta-analysis was similarly performed by combining cohorts with diverse ancestry using inverse-variance-weighted fixed-effects models in METAL^63^. Given that >80% of the included participants were of EUR ancestry, the LD structure of the EUR HRC reference panel was used to define GWS loci.

For all meta-analyses, SNPs present in <75% of total effective sample size (N_eff_) were removed from the meta-analysis results. In addition, we employed the DENTIST tool for summary data-based analyses, which leverages LD from a reference sample (ancestry matched HRC reference panel v1.0^61^, except LAT and multi-ancestry for which the EUR panel was used) to detect and filter out problematic variants by testing the difference between the observed z-score of a variant and a predicted z-score from the neighbouring variants^64^.

To identify independent association signals (P□< □5□× □10^−8^), the GCTA forward selection and backward elimination process (command ‘cojo-slct’) was applied using the summary statistics from the EAS, EUR and multi-ancestry meta-analysis (both including and excluding the self-report data), with the EAS and EUR HRC reference panels, respectively^65,66^.

The genetic correlation between meta-analyses based on all new cohorts (118,284 cases and 2,448,096 controls) and EUR cohorts from our previous PGC BD GWAS^3^ was *r*_*g*_ = 0.64 (se = 0.02), and *r*_*g*_ = 0.91 (se = 0.04) when excluding self-reported cohorts. Concordance of the direction of associations in the present GWAS with associations in the previously published BD data were evaluated as described previously.^67^

### Heritability and Genetic Correlation

LDSC^13^ was used to estimate the SNP-heritability (h^2^_SNP_) of BD from EUR GWAS summary statistics, including all cohorts as well as sub-groups by ascertainment and BD subtype. Popcorn was used to estimate h^2^_SNP_ of BD from non-EUR GWAS summary statistics.^68^ h^2^_SNP_ was converted to the liability scale using a lifetime BD prevalence of 2%. LDSC bivariate genetic correlations (*r*_*g*_) were also estimated between EUR BD GWASs (with and without self-report data) and eleven other psychiatric disorders as well, as 1390 human diseases and traits via the Complex Traits Genetics Virtual Lab (CTG-VL; https://vl.genoma.io) web platform.^20^ Adjusting for the number of traits tested, the Bonferroni corrected p-value was P < 3.569 ×□10^−5^. Cross-ancestry bivariate genetic correlations were estimated using Popcorn.^68^ Differences in *r*_*g*_ between phenotype pairs were tested as a deviation from 0 using the block jackknife approach implemented in LDSC.^69^

The results of the Clinical and Community cohort meta-analyses were conditioned on genetic risks for BDI and BDII, to account for differences in proportion of the BD subtypes within these cohorts. Conditioning was conducted using multitrait-based conditional and joint analysis using GWAS summary data (mtCOJO),^70^ implemented in GCTA.^65^ mtCOJO is robust to sample overlap between the GWASs of the exposure and outcome. The conditioned summary statistics were evaluated for genetic correlation with SElf-reported BD using LDSC.

### MiXeR

We applied causal mixture models (MiXeR)^15,16,71^ to investigate the genetic architecture of BD, specifically the overlap between Clinical, Community and Self-report samples, as well as BD subtypes. We first computed univariate analyses to estimate the polygenicity, discoverability and heritability of each trait. These were followed by bivariate analyses to compute the number of shared trait-influencing variants between pairs of traits, and finally trivariate analyses to compute the proportion of shared variants between all three traits analysed. We also determined the correlation of effect sizes of SNPs within the bivariate shared components.

### Polygenic prediction of bipolar disorder

We used PRS-CS-auto^21^ to compute polygenic risk scores in target cohorts, using a discovery GWAS where the target cohort was left out. Given that the majority of the individuals included in the meta-analysis were of EUR descent, we used the EUR LD reference panel based on UK BioBank data as provided by PRS-CS developers (https://github.com/getian107/PRScs). Raw scores were standardised to Z scores, and covariates including sex, the first five PCs and any others as required (as above for each cohort GWAS) were included in the logistic regression model, via the glm() function in R, with family=binomial and link=logit. The variance explained by PRS (R^2^) was first converted to Nagelkerke’s pseudo-R^2^ via the fmsb package in R, then converted to the liability scale to account for proportion of cases in each cohort and the population prevalence of BD.^72^ We provide R^2^ values for BD assuming a population prevalence of 2%, based upon a recent multinational survey.^14^ The weighted average R^2^ values were then calculated using the N_eff_ for each cohort. PRS-specific-medians and their confidence intervals were computed using nonparametric bootstrap replicates (10,000 resamples with replacement). The odds ratios for BD for individuals in the top quintile of PRS compared with those in the middle quintile were calculated for all cohorts. Similarly, the area under the curve (AUC) statistic was calculated via the pROC package in R^73^, for which we performed a training and testing procedure by taking 80% of the individuals in a given cohort on which to train the model, and tested the predictability in the remaining 20% of individuals. Ten random samplings of training and testing sets were performed in all cohorts, and the median AUC after all permutations is provided Supplementary Tables 16-22. The median confidence intervals for the AUC were similarly averaged across the ten random permutations. These AUC statistics were calculated based on the logistic regression model that includes the standardised PRS as a predictor and PC covariates. In order to assess the gain in AUC due to the PRS itself, we subtracted the median AUC of the model containing only the covariates from the full model, reported in Supplementary Tables 16-22 as AUC.

### Gene-Level and gene-set association analysis

Gene-level, gene-set and tissue-set associations were performed using a SNP-wise mean model (±10kb window) implemented in MAGMA^22^. Bonferroni correction was used to control for multiple testing. In addition, we performed gene-set analysis with GSA-MiXeR^19^, which quantifies partitioned heritability attributed to N=10,475 gene-sets from the GO^74^ and SynGO^34^ databases, alongside their fold enrichment with respect to a baseline model. The GSA-MiXeR full model incorporates 18,201 protein-coding genes, using a joint model to estimate heritability attributed to each gene based on GWAS summary statistics and HRC^59^ reference panel to account for LD between variants. GSA-MiXeR’s baseline model accounts for a set of 75 functional annotations^75^, as well as accounting for MAF- and LD-dependent genetic architecture. GSA-MiXeR’s heritability model is estimated using Adam (method for stochastic gradient-based optimization of the likelihood function)^76^. Standard errors of fitted parameters were estimated from the observed Fisher’s information matrix (the negative Hessian matrix of the log-likelihood function).

Identified credible genes were further assessed for enrichment in synaptic processes using the SynGO tool (https://www.syngoportal.org/) with default settings ^34^.

### Cell type specific enrichment analyses

Single-cell enrichment analyses of brain cell types were performed according to Mullins et al. (2021). Briefly, from five publicly available single-cell RNA sequencing datasets derived from human ^77,78^ and murine ^79–81^ brain tissues, the 10% of genes with highest gene expression specificity per cell type were extracted. After MAGMA^22^ gene analysis of the multi-ancestry GWAS summary statistics including self-report data using an annotation window of 35 kb upstream and 10 kb downstream of the gene boundaries and the 1000 Genomes phase 3 EUR reference panel, MAGMA gene-set analyses were conducted for all cell types in each dataset, respectively. Within each dataset, FDR-adjusted p-values below 0.05 were considered statistically significant.

In addition, we performed an exploratory single-cell enrichment analysis in 914 cell types across 29 non-brain murine tissues as implemented in FUMA^82^. Cell types with FDR-adjusted p-values below 0.05 were considered statistically significant. Moreover, to determine that identified enrichment was not due to overlapping genes with neuronal cell-types we performed cross-dataset conditional analyses of significantly enriched cell-types with murine brain tissue.

### Single-nucleus RNA-seq enrichment analyses

We used the Human Brain Atlas single-nucleus RNA-seq (snRNAseq) dataset^23^ consisting of 3.369 million nuclei sequenced using snRNAseq. The nuclei were from adult postmortem donors, and the dissections focused on 106 anatomical locations within 10 brain regions. Following quality control, the nuclear gene expression patterns allowed the identification of a hierarchy of cell types that were organized into 31 superclusters and 461 clusters. In the current paper we use the same naming system for the cell types and the brain regions as in Siletti et al^23^. We estimated SNP-heritability enrichment for the top decile of expression proportion (TDEP) genes (∼1,300 genes) in each of the 31 superclusters and 461 clusters, respectively, using stratified LD score regression (S-LDSC)^24^ as described previously.^25^ We used FDR correction (FDR < 0.05) to account for multiple comparisons.

### Fine-mapping

We performed functional fine-mapping of GWS loci via Polyfun-SuSiE^26^, using functional annotations of the baseline-LF2.2 UKB model and LD estimates from the Haplotype Reference Consortium (HRC) EUR (N = 21,265) reference panel. The maximum number of causal variants per fine-mapped region was adjusted accordingly based on the results from the conditional analysis. We excluded loci that fall within the MHC locus (6:28000000-34000000, build GRCh37) due to the known complexity of the LD architecture in that region. GWS loci ranges with a LD r2 above 0.1 were used as fine-mapping ranges. Putatively causal SNPs (PIP > 0.50 and part of 95% credible set) were mapped to genes by performing variant annotation with Variant Effect Predictor (VEP) (GRCh37) Ensembl release 75^27^.

### Convergence of common and rare variant signal

Data from the Bipolar Exome (BipEx) consortium^4^ (13,933 BD cases and 14,422 controls) were used to assess the convergence of common and rare variant signals, using a similar approach as previously used for schizophrenia^41^. This dataset includes approximately 8,2 k individuals with BDI and 3,4 k individuals with BDII, while the remainder of the sample lack BD sub-type information. Ultra rare variants (<=5 minor allele count) for damaging missense (missense badness, PolyPhen-2 and regional constraint (MPC) score >3) and protein-truncating variants (including: transcript ablation, splice acceptor variants, splice donor variants, stop gained and frameshift variants) were considered. An enrichment of rare variants in genes prioritised through fine-mapping in cases relative to controls were assessed using a Fisher’s Exact Test. Given the genetic overlap between bipolar disorder and schizophrenia, we repeated the analysis in data from the Schizophrenia Exome Meta-analysis (SCHEMA) cohort (24,248 schizophrenia cases and 97,322 controls)^28^. Using the same approach as taken in the SCHEMA^28^ and BipEx^4^ papers, background genes included all genes surveyed in each sequencing study, respectively.

### QTL integrative analysis

We conducted different QTL integration analyses to elucidate molecular mechanisms by which variants associated with BD might be linked to the phenotype. Summary data-based Mendelian randomization (SMR) (v1.3)^83^ with subsequent heterogeneity in dependent instruments (HEIDI)^70^ tests were performed for expression quantitative trait loci (eQTLs), splicing quantitative trait loci (sQTLs), and methylation quantitative trait loci (mQTLs). Data on eQTLs and sQTLs were obtained from the BrainMeta study v2 (n = 2865),^84^ while data on methylation quantitative trait loci (mQTLs) were obtained from the Brain-mMeta study v1 (n = 1160).^85^ Putatively causal SNPs identified from fine-mapping, as outlined above, were used as the QTL instruments for the SMR analyses. Using the BD GWAS and QTL summary statistics, each putative causal SNP was analysed as the target SNP for probes within a 2 Mb window on either side using the --extract-target-snp-probe option in SMR. The EUR HRC LD reference panel was used for the analyses of the multi-ancestry meta-analysis. A Bonferroni correction was applied for 2021 tests, i.e. SNP-QTL probe combinations, in the eQTL analysis (P_SMR_ < 2.47 ×□10^−5^), 6755 tests in the sQTL analysis (P_SMR_ < 7.40 ×□10^−6^) and 2222 tests in the mQTL analysis (P_SMR_ < 2.25 ×□10^−5^). The significance threshold for the HEIDI test (heterogeneity in dependent instruments) was P_HEIDI_ ≥ 0.01. Additional eQTL integration analyses were conducted using TWAS, FOCUS and isoTWAS. Details related to these analyses are provided in the Supplementary Note.

### Enhancer-promoter gene interactions

To investigate enhancer-promoter (E-P) interactions influenced by BD GWAS variants, we utilized cell-type-specific E-P maps from a multi-omics dataset, which included joint snATAC-seq & snRNA-seq and cell-specific Hi-C data from developing brains. We employed the activity-by-contact (ABC) model^32,33^ for this analysis. Following the authors’ guidelines, we excluded E-P interactions that (i) had an ABC score below 0.015, (ii) involved ubiquitously expressed genes or genes on the Y chromosome, or (iii) included genes not expressed in major brain cell types. Focusing on the BD GWAS, we selected only those E-P links that overlapped genome-wide significant SNPs (with peaks extended by 100 bp on both sides to increase overlap) or their LD buddies (R2 ≥ 0.8). This selection process yielded 11,023 E-P links. We then overlapped these putative disease-relevant variants with enhancer-promoter (E-P) links to prioritize causal genes.

To avoid multiple associations for a single variant, we applied the ABC-Max approach,^33^ retaining only the E-P links with the highest ABC score for each peak.

### Credible gene identification

We provide a set of credible genes by integrating information from various gene-mapping strategies, using a similar approach previously described (Supplementary Figure 9, Supplementary Table 31).^86^ First, genes identified through fine-mapping, and QTL (eQTL, mQTL and sQTL) analyses using SMR and proximity (nearest gene within 10 kb) to fine-mapped putatively causal SNPs were included. The identified set of 116 genes were then further assessed based on gene-level associations (MAGMA),^22^ additional integrative eQTL analyses^30,31^ and enhancer-promoter gene interactions.^32,33^ The criteria for filtering genes from the different eQTL methods were: (i) SMR adjusted p-value less than 0.05 and HEIDI test p-value greater than 0.01, (ii) TWAS adjusted p-values less than 0.05 and colocalization probability (COLOC.PP4) greater than 0.7, (iii) FOCUS posterior inclusion probability greater than 0.7 and within a credible set, (iv) isoTWAS permutation p-value less than 0.05, isoTWAS poster inclusion probability greater than 0.7 and within a credible set (Supplementary Figure 9). Genes annotated by at least one of these eQTL approaches were confirmed as having eQTL evidence (Supplementary Table 31). Thus, seven approaches were considered by which loci could be mapped to genes including, eQTL evidence (eQTL or TWAS or FOCUS or isoTWAS), mQTL, sQTL, VEP, proximity, MAGMA and E-P interactions.

### Clustering of credible genes by temporal variation

Lifespan gene expression from the Human Brain Transcriptome project (www.hbatlas.org)^35^ was used to cluster the list of credible genes based on their temporal variation. The gene expression and associated metadata were acquired from the gene expression omnibus (GEO; accession: GSE25219). The data consists of 57 donors aged 5.7 weeks post conception to 82 years old with samples extracted across regions of the brain. Prior to filtering gene expression for the list of credible genes, gene symbols of both credible genes and the gene expression dataset were harmonized using the “limma” package in R which updates any synonymous gene symbols to the latest Entrez symbol. Gene expression was available for 33 of the 36 credible genes. Within a given brain region, each gene’s expression was then mean-centered and scaled. Outliers in gene expression more than 4 standard deviations from the mean were removed. To generate a single gene expression profile for each gene across the lifespan, at a given age, the mean gene expression for a given gene was taken across brain regions, and in some cases across donors. This resulted in a matrix where each gene had a single expression value for each age across the lifespan. This gene-expression by age matrix was then used to cluster the credible genes by the lifespan expression profiles using the R package “TMixClust”. This method uses mixed-effects models with nonparametric smoothing splines to capture and cluster non-linear variation in temporal gene expression. We tested K=2 to K=10 clusters performing 50 clustering runs to analyse stability. The clustering solution with the highest likelihood (i.e., the global optimum using an expectation maximization technique) is selected as the most stable solution across the 50 runs for each of the trials testing 2-10 clusters. We compare the average silhouette width across the K=2 to K=10 clusters and select that with the maximum value as the optimal number of clusters.

### Drug enrichment analyses

Gene-set analyses were performed restricted to genes targeted by drugs, assessing individual drugs and grouping drugs with similar actions as described previously ^3,36^. Gene-level and gene-set analyses of the multi-ancestry GWAS summary statistics including self-report data were performed in MAGMA v1.10 ^22^, as outlined above for cell type specific enrichment.

Gene sets were defined comprising the targets of each drug in the Drug Gene Interaction database DGIdb v.5.0.6 ^37^; the Psychoactive Drug Screening Database Ki DB ^87^; ChEMBL v27 ^88^; the Target Central Resource Database v6.7.0 ^89^; and DSigDB v1.0 ^90^; all downloaded in October 2020. Multiple testing was controlled using a Bonferroni-corrected significance threshold of P□< □5.41□× □10^−5^ (924 drug-sets with at least ten valid drug gene sets) for drug-set analysis and P□ □ □5.49□× □10^−4^ (91 drug classes) for drug-class analysis, respectively. We also assessed whether any of the 36 credible genes were classified as druggable in the OpenTargets platform.

In addition, gene-set analyses were also performed to test the enrichment of drug-gene interactions on only credible genes as described above. Moreover, we investigated if any lithium target genes, as well as their interaction partners, were among the 36 credible genes using the latest version of the human protein interactome^91^. We calculated the network-based separation (S_AB_) between credible genes and lithium target genes, where a significant overlapping network neighbourhood would be indicative of functional similarity^92^.

## Supporting information

Cohort descriptions

Author list and affiliations

## Data Availability

The policy of the PGC is to make genome-wide summary results public. Summary statistics will be made available through the PGC website upon publication (https://www.med.unc.edu/pgc/results-and-downloads). Data are accessible with collaborative analysis proposals through the Bipolar Disorder Working Group of the PGC (https://www.med.unc.edu/pgc/shared-methods/how-to/). This study included some publicly available datasets accessed through dbGaP - PGC bundle phs001254.v1.p1. We will provide 23andMe summary statistics for the top 10,000 SNPs upon publication. Full GWAS summary statistics will be made available through 23andMe to qualified researchers under an agreement with 23andMe that protects the privacy of the 23andMe participants. Please visit https://research.23andme.com/collaborate/#dataset-access/ for more information and to apply to access the data.

## Data availability

Genome-wide association summary statistics for these analyses will be made available upon publication at https://www.med.unc.edu/pgc/download-results/. The full GWAS summary statistics for the 23andMe datasets will be made available through 23andMe to qualified researchers under an agreement with 23andMe that protects the privacy of the 23andMe participants. Please visit https://research.23andme.com/collaborate/#dataset-access for more information and to apply to access the data. After applying with 23andMe, the full summary statistics including all analysed SNPs and samples in the GWAS meta-analyses will be accessible to the approved researchers. Genotype data are available for a subset of cohorts, including dbGAP accession numbers and/or restrictions, as described in the ‘Cohort descriptions’ section of the supplementary materials.

## Code availability

Core analysis code for RICOPILI can be found at https://sites.google.com/a/broadinstitute.org/ricopili/. This wraps PLINK (https://www.cog-genomics.org/plink2/), EIGENSOFT (https://www.hsph.harvard.edu/alkes-price/software/), Eagle2 (https://alkesgroup.broadinstitute.org/Eagle/), Minimac3 (https://genome.sph.umich.edu/wiki/Minimac3), SHAPEIT3 (https://mathgen.stats.ox.ac.uk/genetics_software/shapeit/shapeit.html), METAL (https://genome.sph.umich.edu/wiki/METAL_Documentation) and LDSR (https://github.com/bulik/ldsc). For downstream analyses, MiXeR can be found at https://github.com/precimed/mixer, PRS-cs is at https://github.com/getian107/PRScs and GSA-MiXeR is available at https://github.com/precimed/gsa-mixer. MAGMA can be found at https://ctg.cncr.nl/software/magma. PolyFun+SuSiE (https://github.com/omerwe/polyfun), TWAS (http://gusevlab.org/projects/fusion/), isoTWAS (https://github.com/bhattacharya-a-bt/isotwas) and SMR (https://github.com/XudongHan-bio/SMR) are also freely available.

