## Supplementary material for "Genomics yields biological and phenotypic insights into bipolar disorder": Cohort descriptions

### **Sample descriptions**

We included data from 79 cohorts **(Supplementary Table 1)**, totaling 158,036 cases and 2,796,499 controls of European, East Asian, African American and Latinx descent. For 53 cohorts, raw genotype and phenotype data were shared with the Psychiatric Genomics Consortium (PGC). Cases were required to meet international consensus criteria (DSM-IV, ICD-9, or ICD-10) for a lifetime diagnosis of bipolar disorder (BD) established using structured diagnostic instruments from assessments by trained interviewers, clinician-administered checklists, or medical record review. Controls in most samples were screened for the absence of lifetime psychiatric disorders, as indicated. For the remaining 26 external cohorts, GWAS summary statistics for BD were shared with the PGC. Cases in these cohorts were largely defined using ICD codes ascertained from medical records. All samples in previous PGC BD GWAS papers were included^1–3^.

Below we describe the ascertainment and diagnosis of the participants in each individual cohort comprising this report. Most cohorts have been published on individually, and the primary report can usually be found using the PubMed identifiers provided. The lead PI of each sample warranted that their protocol was approved by their local Ethical Committee and that all participants provided written informed consent. **Supplementary Table 1** provides additional detail, including sample sizes and genotyping array. As the lifetime prevalence of BD is around 1-2%, some cohorts use controls that are not screened for BD^4,5^. The boldfaced first line for each sample indicates study PI, PubMed ID if published, country (study name), and the PGC internal tag or study identifier.

***========* PGC1 Samples *========***

**Rietschel, M; Nöthen, MM, Cichon, S | 21926972 [PGC1] | BOMA-Germany I | bip_bonn_eur**

Cases for the BOMA-Bipolar Study were ascertained from consecutive admissions to the inpatient units of the Department of Psychiatry and Psychotherapy at the University of Bonn and at the Central Institute for Mental Health in Mannheim, University of Heidelberg, Germany. DSM-IV lifetime diagnoses of bipolar I disorder were assigned using a consensus best-estimate procedure, based on all available information, including a structured interview with the SCID and SADS-L, medical records, and the family history method. In addition, the OPCRIT^6^ checklist was used for the detailed polydiagnostic documentation of symptoms. Controls were ascertained from three population-based studies in Germany (PopGen, KORA, and Heinz-Nixdorf-Recall Study). The control subjects were not screened for mental illness. Study protocols were reviewed and approved in advance by Institutional Review Boards of the participating institutions. All subjects provided written informed consent.

**Corvin, A | 18711365 [PGC1] | Ireland | bip_dub1_eur**

Samples were collected as part of a larger study of the genetics of psychotic disorders in the Republic of Ireland, under protocols approved by the relevant IRBs and with written informed consent that permitted repository use. Cases were recruited from Hospitals and Community psychiatric facilities in Ireland by a psychiatrist or psychiatric nurse trained to use the SCID. Diagnosis was based on the structured interview supplemented by case note review and collateral history where available. All diagnoses were reviewed by an independent reviewer. Controls were ascertained with informed consent from the Irish GeneBank and represented blood donors who met the same ethnicity criteria as cases. Controls were not specifically screened for psychiatric illness.

**Blackwood, D | 18711365 [PGC1] | Edinburgh, UK | bip_edi1_eur**

This sample comprised Caucasian individuals contacted through the inpatient and outpatient services of hospitals in South East Scotland. A BD-I diagnosis was based on an interview with the patient using the SADS-L supplemented by case note review and frequently by information from medical staff, relatives and caregivers. Final diagnoses, based on DSM-IV criteria, were reached by consensus between two trained psychiatrists. Ethnically-matched controls from the same region were recruited through the South of Scotland Blood Transfusion Service. Controls were not directly screened to exclude those with a personal or family history of psychiatric illness. The study was approved by the Multi-Centre Research Ethics Committee for Scotland and patients gave written informed consent for the collection of DNA samples for use in genetic studies.

**Kelsoe, J | 21926972 [PGC1] | USA (GAIN) | bip_gain_eur**

*Genetic Association Information Network (GAIN)/ The Bipolar Genome Study (BiGS)* The BD sample was collected under the auspices of the NIMH Genetics Initiative for BD (<http://zork.wustl.edu/nimh/>), genotyped as part of GAIN and analyzed as part of a larger GWAS conducted by the BiGS consortium. Approximately half of the GAIN sample was collected as multiplex families or sib pair families (waves 1-4), the remainder were collected as individual cases (wave 5). Subjects were ascertained at 12 sites: Indiana University, John Hopkins University, the NIMH Intramural Research Program, Washington University at St. Louis, University of Pennsylvania, University of Chicago, Rush Medical School, University of Iowa, University of California, San Diego, University of California, San Francisco, Howard University, and University of Michigan. All investigations were carried out after the review of protocols by the IRB at each participating institution. At all sites, potential cases were identified from screening admissions to local treatment facilities and through publicity programs or advocacy groups. Potential cases were evaluated using the DIGS^7^, FIGS^8^, and information from relatives and medical records. All information was reviewed through a best estimate diagnostic procedure by two independent and non-interviewing clinicians and a consensus best-estimate diagnosis was reached. In the event of a disagreement, a third review was done to break the tie. Controls were from the NIMH Genetic Repository sample obtained by Dr. P. Gejman through a contract to Knowledge Networks, Inc. Only individuals with complete or near-complete psychiatric questionnaire data who did not fulfill diagnostic criteria for major depression and denied a history of psychosis or BD were included as controls for BiGS analyses. Controls were matched for gender and ethnicity to the cases.

**Scott, L; Myer, RM; Boehnke, M | 19416921 [PGC1] | Michigan, USA (Pritzker and NIMH) | bip_mich_eur**

The Pritzker Neuropsychiatric Disorders Research Consortium (NIMH/Pritzker) case and control samples were from the NIMH Genetics Initiative Genetics Initiative Repository. Cases were diagnosed according to DMS-III or DSM-IV criteria using diagnostic interviews and/or medical record review. Cases with low confidence diagnoses were excluded. From each wave 1-5 available non-Ashkenazi European-origin family, two BD1 siblings were included when possible and the proband was preferentially included if available (n=946 individuals in 473 sibling pairs); otherwise a single BD1 case was included (n=184). The bipolar sibling pairs were retained within the NIMH/Pritzker sample when individuals in more than one study were uniquely assigned to a study set. Controls had non-Ashkenazi European-origin, were aged 20-70 years and reported no diagnosis with or treatment for BD or schizophrenia, and that they had not heard voices that others could not hear. Individuals with suspected major depression were excluded based on answers to questions related to depressive mood. NIMH controls were further selected as the best match(es) to NIMH cases based on self-reported ancestry.

**Sklar, P; Smoller, J | 18317468 [PGC1] | USA (STEP1) | bip_stp1_eur**

The Systematic Treatment Enhancement Program for Bipolar Disorder (STEP-BD) was a seven-site, national U.S., longitudinal cohort study designed to examine the effectiveness of treatments and their impact on the course of BD that enrolled 4,361 participants who met DSM-IV criteria for BD1, BD2, bipolar not otherwise specified (NOS), schizoaffective manic or bipolar type, or cyclothymic disorder based on diagnostic interviews. From the parent study, 2,089 individuals who were over 18 years of age with BD1 and BD2 diagnoses consented to the collection of blood samples for DNA. BD samples with a consensus diagnosis of BD1 were selected for inclusion in STEP1. Two groups of controls samples from the NIMH repository were used. One comprised DNA samples derived from US Caucasian anonymous cord blood donors. The second were controls who completed the online self-administered psychiatric screen and were ascertained as described above, by Knowledge Networks Inc. For the second sample of controls only those without a history of schizophrenia, psychosis, BD or major depression with functional impairment were used.

**Sklar, P; Smoller, J | 18711365 [PGC1] | USA (STEP2) | bip_stp2_eur**

The STEP2 sample included BD-1 and BD-2 samples from the STEP-BD study described above along with BD-2 subjects from UCL study also described above. The controls samples for this study were from the NIMH repository as described above for the STEP1 study.

**Andreassen, OA | PMID:21926972 [PGC1], PMID:20451256 | Norway (TOP) | bip_top7_eur**

In the TOP study (Tematisk omrade psykoser), cases of European ancestry, born in Norway, were recruited from psychiatric hospitals in the Oslo region. Patients were diagnosed according to the SCID^9^ and further ascertainment details have been reported. Healthy control subjects were randomly selected from statistical records of persons from the same catchment area as the patient groups. The control subjects were screened by interview and with the Primary Care Evaluation of Mental Disorders (PRIME-MD)^10^. None of the control subjects had a history of moderate/severe head injury, neurological disorder, mental retardation or an age outside the age range of 18-60 years. Healthy subjects were excluded if they or any of their close relatives had a lifetime history of a severe psychiatric disorder. All participants provided written informed consent and the human subjects protocol was approved by the Norwegian Scientific-Ethical Committee and the Norwegian Data Protection Agency.

**McQuillin, A; Gurling, H | 18317468 [PGC1] | UCL (University College London), London, UK | bip_uclo_eur**

The UCL sample comprised Caucasian individuals who were ascertained and received clinical diagnoses of bipolar 1 disorder according to UK National Health Service (NHS) psychiatrists at interview using the categories of the International Classification of Disease version 10. In addition bipolar subjects were included only if both parents were of English, Irish, Welsh or Scottish descent and if three out of four grandparents were of the same descent. All volunteers read an information sheet approved by the Metropolitan Medical Research Ethics Committee who also approved the project for all NHS hospitals. Written informed consent was obtained from each volunteer. The UCL control subjects were recruited from London branches of the National Blood Service, from local NHS family doctor clinics and from university student volunteers. All control subjects were interviewed with the SADS-L to exclude all psychiatric disorders.

**Craddock, N, Jones, I, Jones, L | 17554300 | WTCCC | bip_wtcc_eur_sr-qc**

Cases were all over the age of 17 yr, living in the UK and of European descent. Recruitment was undertaken throughout the UK and included individuals who had been in contact with mental health services and had a lifetime history of high mood. After providing written informed consent, participants were interviewed by a trained psychologist or psychiatrist using a semi-structured lifetime diagnostic psychiatric interview (Schedules for Clinical Assessment in Neuropsychiatry) and available psychiatric medical records were reviewed. Using all available data, best-estimate life-time diagnoses were made according to the RDC^12^. In the current study we included cases with a lifetime diagnosis of RDC bipolar 1 disorder, bipolar 2 disorder or schizo-affective disorder, bipolar type.

Controls were recruited from two sources: the 1958 Birth Cohort study and the UK Blood Service (blood donors) and were not screened for history of mental illness.

All cases and controls were recruited under protocols approved by the appropriate IRBs. All subjects gave written informed consent.

***========* PGC2 Samples *========***

**Adolfsson, R | Not published | Umeå, Sweden | bip_ume4_eur**

Clinical characterization of the patients included the Mini-International Neuropsychiatric Interview (MINI^11^), the Diagnostic Interview for Genetic Studies (DIGS^7^), the Family Interview for Genetic Studies (FIGS^8^) and the Schedules for Clinical Assessment in Neuropsychiatry (SCAN)^12^. The final diagnoses were made according to the DSM-IV-TR and determined by consensus of 2 research psychiatrists. The unrelated Swedish control individuals, consisting of a large population-based sample representative of the general population of the region, were randomly selected from the ‘Betula study’.

**Alda, M; Smoller, J | Not published | Nova Scotia, Canada; I2B2 controls | bip_hal2_eur**

The case samples were recruited from patients longitudinally followed at specialty mood disorders clinics in Halifax and Ottawa (Canada). Cases were interviewed in a blind fashion with the Schedule of Affective Disorders and Schizophrenia-Lifetime version (SADS-L)^13^ and consensus diagnoses were made according to DSM-IV^14^ and Research Diagnostic Criteria (RDC)^15^. Protocols and procedures were approved by the local Ethics Committees and written informed consent was obtained from all patients before participation in the study. Control subjects were drawn from the I2B2 (Informatics for Integrating Biology and the Bedside) project^16^. The study consists of de-identified healthy individuals recruited from a healthcare system in the Boston, MA, US area. The de-identification process meant that the Massachusetts General Hospital Institutional Review Board elected to waive the requirement of seeking informed consent as detailed by US Code of Federal Regulations, Title 45, Part 46, Section 116 (46.116).

**Andreassen, OA | Not published | Norway (TOP) | bip_top8_eur**

The TOP8 bipolar disorder cases and controls were ascertained in the same way as the bip_top7_eur (TOP7) samples described above, and recruited from hospitals across Norway.

**Biernacka, JM; Frye, MA | 27769005 | Mayo Clinic, USA | bip_may1_eur**

Bipolar cases were drawn from the Mayo Clinic Bipolar Biobank^17^. Enrolment sites included Mayo Clinic, Rochester, Minnesota; Lindner Center of HOPE/University of Cincinnati College of Medicine, Cincinnati, Ohio; and the University of Minnesota, Minneapolis, Minnesota. Enrolment at each site was approved by the local Institutional Review Board, and all participants consented to use of their data for future genetic studies. Participants were identified through routine clinical appointments, from in-patients admitted in mood disorder units, and recruitment advertising. Participants were required to be between 18 and 80 years old and be able to speak English, provide informed consent, and have DSM-IV-TR diagnostic confirmation of type 1 or 2 bipolar disorder or schizoaffective bipolar disorder as determined using the SCID. Controls were selected from the Mayo Clinic Biobank^18^. Potential controls with ICD9 codes for bipolar disorder, schizophrenia or related diagnoses in their electronic medical record were excluded.

**Breen, G; Vincent, JB | 24387768; 19416921; 21926972 [PGC1] |London, UK; Toronto, Canada [BACC] |bip_bac1_eur**

The total case/control cohort (N=1922) includes 871 subjects from Toronto, Canada (N=431 cases (160 male; 271 female); N=440 controls (176 male; 264 female)), 1051 subjects from London, UK (N=538 cases (180 male; 358 female); N=513 controls (192 male; 321 female)). A summary of mean and median age at interview, age of onset (AOO), diagnostic subtypes (BD 1 versus BD 2), presence of psychotic symptoms, suicide attempt and family history of psychiatric disorders has been provided previously for both the Toronto and London cohorts^19^. From the Toronto site (Centre for Addiction & Mental Health (CAMH)), BD individuals and unrelated healthy controls matched for age, gender and ethnicity were recruited. Inclusion criteria for patients: a) diagnosed with DSMIV/ICD 10 BD 1 or 2; b) 18 years old or over; c) Caucasian, of Northern and Western European origin, and three out of four grandparents also N.W. European Caucasian. Exclusion criteria include: a) Use of intravenous drugs; b) Evidence of intellectual disability; c) Related to an individual already in the study; d) Manias that only ever occurred in relation to or resulting from alcohol or substance abuse/dependence, or medical illness; e) Manias resulting from non-psychotropic substance usage. The SCAN interview (Schedule for Clinical Assessments in Neuropsychiatry) was used for subject assessment^20^. Using the SCAN interview along with case note review, each case was assigned DSM-IV and ICD 10 diagnoses by two independent diagnosticians, according to lifetime consensus best-estimate diagnosis. Lifetime occurrence of psychiatric symptoms was also recorded using the OPCRIT checklist, modified for use with mood disorders. Similar methods and criteria were also used to collect a sample of 538 BD cases and 513 controls for the London cohort (King’s College London; KCL)^21^.

Both studies were approved by respective institutional research ethics committees (the CAMH Research Ethics Board (REB) in Toronto, and the College Research Ethics Committee (CREC) at KCL), and informed written consent was obtained from all participants. GWAS results have previously been published for the entire KCL/CAMH cohort^22^.

**Rietschel, M; Nöthen, MM; Schulze, TG; Reif, A; Forstner, AJ | 24618891 | BOMA-Germany II | bip_bmg2_eur**

Cases were recruited from consecutive admissions to psychiatric in-patient units at the University Hospital Würzburg. All cases received a lifetime diagnosis of BD according to the DSM-IV criteria using a consensus best-estimate procedure based on all available information, including semi-structured diagnostic interviews using the Association for Methodology and Documentation in Psychiatry^23^, medical records and the family history method. In addition, the OPCRIT system was used for the detailed polydiagnostic documentation of symptoms.

Control subjects were ascertained from the population-based Heinz Nixdorf Recall (HNR) Study^24^. The controls were not screened for a history of mental illness. Study protocols were reviewed and approved in advance by Institutional Review Boards of the participating institutions. All subjects provided written informed consent.

**Rietschel, M; Nöthen, MM; Schulze, TG; Bauer, M; Forstner, AJ; Müller-Myhsok, B | 24618891 | BOMA-Germany III | bip_bmg3_eur**^25^

Cases were recruited at the Central Institute of Mental Health in Mannheim, University of Heidelberg, and other collaborating psychiatric hospitals in Germany. All cases received a lifetime diagnosis of BD according to the DSM-IV criteria using a consensus best-estimate procedure based on all available information including structured diagnostic interviews using the AMDP, Composite International Diagnostic Screener (CID-S)^26^, SADS-L and/or SCID, medical records, and the family history method. In addition, the OPCRIT system was used for the detailed polydiagnostic documentation of symptoms.

Controls were selected randomly from a Munich-based community sample and recruited at the Max-Planck Institute of Psychiatry. They were screened for the presence of anxiety and mood disorders using the CID-S. Only individuals without mood and anxiety disorders were collected as controls. Study protocols were reviewed and approved in advance by Institutional Review Boards of the participating institutions. All subjects provided written informed consent.

**Hauser, J; Lissowska, J; Forstner, AJ | 24618891 | BOMA-Poland | bip_bmpo_eur**

Cases were recruited at the Department of Psychiatry, Poznan University of Medical Sciences, Poznan, Poland. All cases received a lifetime diagnosis of BD according to the DSM-IV criteria on the basis of a consensus best-estimate procedure and structured diagnostic interviews using the SCID. Controls were drawn from a population-based case-control sample recruited by the Cancer-Center and Institute of Oncology, Warsaw, Poland and a hospital-based case-control sample recruited by the Nofer Institute of Occupational Medicine, Lodz, Poland. The Polish controls were produced by the International Agency for Research on Cancer (IARC) and the Centre National de Génotypage (CNG) GWAS Initiative for a study of upper aerodigestive tract cancers. The controls were not screened for a history of mental illness. Study protocols were reviewed and approved in advance by Institutional Review Boards of the participating institutions. All subjects provided written informed consent.

**Rietschel, M; Nöthen, MM; Rivas, F; Mayoral, F; Kogevinas, M; others | 24618891 | BOMA-Spain | bip_bmsp_eur**

Cases were recruited at the mental health departments of the following five centers in Andalusia, Spain: University Hospital Reina Sofia of Córdoba, Provincial Hospital of Jaen; Hospital of Jerez de la Frontera (Cádiz); Hospital of Puerto Real (Cádiz); Hospital Punta Europa of Algeciras (Cádiz); and Hospital Universitario San Cecilio (Granada). Diagnostic assessment was performed using the SADS-L; the OPCRIT; a review of medical records; and interviews with first and/or second degree family members using the Family Informant Schedule and Criteria (FISC)^27^. Consensus best estimate BD diagnoses were assigned by two or more independent senior psychiatrists and/or psychologists, and according to the RDC, and the DSM-IV. Controls were Spanish subjects drawn from a cohort of individuals recruited in the framework of the European Community Respiratory Health Survey (ECRHS, http://www.ecrhs.org/). The controls were not screened for a history of mental illness. Study protocols were reviewed and approved in advance by Institutional Review Boards of the participating institutions. All subjects provided written informed consent.

**Fullerton, J.M.; Mitchell, P.B.; Schofield, P.R.; Martin N.G.; Cichon, S. | 24618891 | BOMA-Australia | bip_bmau_eur**

Cases were recruited at the Mood Disorder Unit, Prince of Wales Hospital in Sydney. All cases received a lifetime diagnosis of BD according to the DSM-IV criteria on the basis of a consensus best-estimate procedure^19^ and structured diagnostic interviews using the DIGS, FIGS, and the SCID. Controls were parents of unselected adolescent twins from the Brisbane Longitudinal Twin Study. The controls were not screened for a history of mental illness. Study protocols were reviewed and approved in advance by Institutional Review Boards of the participating institutions. All subjects provided written informed consent.

**Grigoroiu-Serbanescu, M; Nöthen, MM | 21353194 | BOMA-Romania | bip_rom3_eur**

Cases were recruited from consecutive admissions to the Obregia Clinical Psychiatric Hospital, Bucharest, Romania. Patients were administered the DIGS^28^ and FIGS^8^ interviews. Information was also obtained from medical records and close relatives. The diagnosis of BP-I was assigned according to DSM-IV criteria using the best estimate procedure. All patients had at least two hospitalized illness episodes. Population-based controls were evaluated using the DIGS to exclude a lifetime history of major affective disorders, schizophrenia, schizoaffective disorders, and other psychoses, obsessive-compulsive disorder, eating disorders, and alcohol or drug addiction.

**Kelsoe, J; Sklar, P; Smoller, J | [PGC1 Replication] | USA (FAT2; FaST,** **BiGS, TGEN) | bip_fat2_eur**

Cases were collected from individuals at the 11 U.S. sites described for the GAIN sample. Eligible participants were age 18 or older meeting DSM-IV criteria for BD-I or BD-II by consensus diagnosis based on interviews with the Affective Disorders Evaluation (ADE) and MINI. All participants provided written informed consent and the study protocol was approved by IRBs at each site. Collection of phenotypic data and DNA samples were supported by NIMH grants MH063445 (JW Smoller); MH067288 (PI: P Sklar), MH63420 (PI: V Nimgaonkar) and MH078151, MH92758 (PI: J. Kelsoe). The control samples were NIMH controls that were using the methods described in that section. The case and control samples were independent of those included in the GAIN sample.

**Kirov, G | 25055870 | Bulgarian trios | bip_butr_eur**

All cases were recruited in Bulgaria from psychiatric inpatient and outpatient services. Each proband had a history of hospitalisation and was interviewed with an abbreviated version of the SCAN. Consensus best-estimate diagnoses were made according to DSM-IV criteria by two researchers. All participants gave written informed consent and the study was approved by local ethics committees at the participating centers.

**Kirov, G | 25055870 | UK trios | bip_uktr_eur**

The BD subjects were recruited from lithium clinics and interviewed in person by a senior psychiatrist, using the abbreviated version of the SCAN. Consensus best-estimate diagnoses were made based on the interview and hospital notes. Ethics committee approval for the study was obtained from the relevant research ethics committees and all individuals provided written informed consent for participation.

**Landén, M; Sklar, P | [ICCBD] | Sweden (ICCBD) | bip_swa2_eur**

The BD subjects were identified using the Swedish National Quality Register for Bipolar Disorders (BipoläR) and the Swedish National Patient Register (using a validated algorithm^29^ requiring at least two hospitalizations with a BD diagnosis). A confirmatory telephone interview with a diagnostic review was conducted. Additional subjects were recruited from the St. Göran Bipolar Project (Affective Center at Northern Stockholm Psychiatry Clinic, Sweden), enrolling new and ongoing patients diagnosed with BD using structured clinical interviews. Diagnoses were made according to the DSM-IV criteria (BipoläR and St. Göran Bipolar Project) and ICD-10 (National Patient Register). The control subjects used were the same as for the SCZ analyses described above. All ascertainment procedures were approved by the Regional Ethical Committees in Sweden.

**Landén, M; Sklar, P | [ICCBD] | Sweden (ICCBD) | bip_swei_eur**

The cases and controls in the bip_swei_eur sample were recruited using the same ascertainment methods described for the bip_swa2_eur sample.

**Leboyer, M |**^30^**; [PGC1 replication] | France | bip_fran_eur**

Cases with BD1 or BD2 and control samples were recruited as part of a large study of genetics of BD in France (Paris-Creteil, Bordeaux, Nancy) with a protocol approved by relevant IRBs and with written informed consent. Cases of French descent for more than 3 generations were assessed by a trained psychiatrist or psychologist using structured interviews supplemented by medical case notes, mood scales and self-rating questionnaire assessing dimensions.

**Li, Q | 24166486; 27769005** **| USA (Janssen), SAGE controls | bip_jst5_eur**

The study included unrelated patients with bipolar 1 disorder from 6 clinical trials (IDs: NCT00253162, NCT00257075, NCT00076115, NCT00299715, NCT00309699, and NCT00309686). Participant recruitment was conducted by Janssen Research & Development, LLC (formerly known as Johnson & Johnson Pharmaceutical Research & Development, LLC) to assess the efficacy and safety of risperidone. Bipolar cases were diagnosed according to DSM-IV-TR criteria. The diagnosis of bipolar disorder was confirmed by the Schedule for Affective Disorders and Schizophrenia for School-Age Children-Present and Lifetime Version (K-SADS-PL) in NCT00076115, by the SCID in NCT00257075 and NCT00253162, or by the MINI in NCT00299715 and NCT00309699, and NCT00309686, respectively. Additional detailed descriptions of these clinical trials can be found at ClinicalTrials.gov. Only patients of European ancestry with matching controls were included in the current analysis. Controls subjects were drawn from the Study of Addiction: Genetics and Environment (SAGE, dbGaP Study Accession: phs000092.v1.p1). Control subjects did not have alcohol dependence or drug dependence diagnoses; however, mood disorders were not an exclusion criterion.

**Craddock, N; Jones, I; Jones, L | [ICCBD] | Cardiff and Worcester, UK (ICCBD-BDRN) | bip_icuk_eur**

Cases were all over the age of 17 yr, living in the UK and of European descent. Cases were recruited via systematic and not systematic methods as part of the Bipolar Disorder Research Network project ([www.bdrn.org](http://www.bdrn.org)), provided written informed consent and were interviewed using a semi-structured diagnostic interview, the Schedules for Clinical Assessment in Neuropsychiatry. Based on the information gathered from the interview and case notes review, best-estimate lifetime diagnosis was made according to DSM-IV. Inter-rater reliability was formally assessed using 20 randomly selected cases (mean ĸ Statistic = 0.85). In the current study we included cases with a lifetime diagnosis of DSM-IV bipolar disorder or schizo-affective disorder, bipolar type. The BDRN study has UK National Health Service (NHS) Research Ethics Committee approval and local Research and Development approval in all participating NHS Trusts/Health Boards.Controls were part of the Wellcome Trust Case Control Consortium common control set, which comprised healthy blood donors recruited from the UK Blood Service and samples from the 1958 British Birth Cohort. Controls were not screened for a history of mental illness. All cases and controls were recruited under protocols approved by the appropriate IRBs. All subjects gave written informed consent.

**Ophoff, RA | Not Published | Netherlands | bip_ucla_eur**

The case sample consisted of inpatients and outpatients recruited through psychiatric hospitals and institutions throughout the Netherlands. Cases with DSM-IV bipolar disorder, determined after interview with the SCID, were included in the analysis. Controls were collected in parallel at different sites in the Netherlands and were volunteers with no psychiatric history after screening with the (MINI^11^). Ethical approval was provided by UCLA and local ethics committees and all participants gave written informed consent.

**Paciga, S | [PGC1] | USA (Pfizer) | bip_pf1e_eur**

This sample comprised Caucasian individuals recruited into one of three Geodon (ziprasidone) clinical trials (NCT00141271, NCT00282464, NCT00483548). Subjects were diagnosed by a clinician with a primary diagnosis of Bipolar 1 Disorder, most recent episode depressed, with or without rapid cycling, without psychotic features, as defined in the DSM-IV-TR (296.5x) and confirmed by the MINI (version 5.0.0). Subjects also were assessed as having a HAM-D-17 total score of >20 at the screening visit. The trials were conducted in accordance with the protocols, International Conference on Harmonization of Good Clinical Practice Guidelines, and applicable local regulatory requirements and laws. Patients gave written informed consent for the collection of blood samples for DNA for use in genetic studies.

**Pato, C | [ICCBD] | Los Angeles, USA (ICCBD-GPC)| bip_usc2_eur**

Genomic Psychiatry Consortium (GPC) cases and controls were collected via the University of Southern California healthcare system, as previously described^31^. Using a combination of focused, direct interviews and data extraction from medical records, diagnoses were established using the OPCRIT and were based on DSM-IV-TR criteria. Age and gender-matched controls were ascertained from the University of Southern California health system and assessed using a validated screening instrument and medical records.

***========* PGC2 Followup Samples *========***

**Kelsoe, J | [PGC1] | USA (BiGS/TGEN1) | TGEN1_eur**

Cases and controls for this sample were ascertained using the same procedures applied for the bip_gain_eur sample described above. These samples formed a distinct PCA cluster from the samples described above and were therefore analysed separately.

**Li, Q | 24166486 | various Eastern Europe, shared T. Esku controls | JJ_EAST_eur**

The cases were drawn from the same six clinical studies described for bip_jst5_eur except that only patients of east European ancestry with matching controls were included in this cohort. Most of the Eastern European controls were from the Estonian Biobank project (EGCUT)^32^ and were ancestrally matched with cases.

**Schulze, T | [ConLiGen] | Germany | BIP_KFO_eur**

The KFO sample was derived from the Clinical Research Group 241 (KFO241 consortium; [www.kfo241.de](http://www.kfo241.de)) and the PsyCourse consortium ([www.psycourse.de](http://www.psycourse.de)). The samples form part of a multi-site German/Austrian longitudinal study. Diagnoses were made according to DSM-IV. German Red Cross controls were collected by the Central Institute for Mental Health in Mannheim, University of Heidelberg, Germany. Volunteers who gave blood to the Red Cross were asked whether they would be willing to participate in genetic studies of psychiatric disorders. Control subjects were not selected on the basis of mental health screening.

***========* External studies PGC3 *========***

**Mortensen, P; Borglum, A | Not published | [iPsych] | NA**

The iPSYCH2015 bipolar disorder sample is a nationwide population based case-cohort sample derived from the Danish Bloodspot resource^33^. In 1981, Denmark began storing neonatal bloodspots and collected samples have been subsequently linked to the Danish Psychiatric Central Research Register (DPCRR). The iPSYCH sample includes practically all individuals diagnosed with bipolar disorder who were born in Denmark between 1981 and 2008. Cases were diagnosed clinically by a psychiatrist at in- or out-patient psychiatric hospitals according to ICD10 as recorded in DPCRR (ICD10 codes F30-F31). Diagnoses were given in 2013 or earlier for persons not less than 10 years old. Controls were randomly selected from the same national birth cohort and not diagnosed with bipolar disorder.

The iPSYCH2015 cohort consists of the initial case-cohort sample iPSYCH2012^34^ and the recent extension iPSYCH2015i.^35^

DNA was prepared as described previously^36^ and genotyping was done using the PsychChip array from Illumina (CA, San Diego, USA) for iPSYCH2012 and the Global Screening Array v2 with a multi disease drop in (Illumina, San Diego, California) according to the manufacturer’s protocols. Genotypes of iPSYCH2012 and iPSYCH2015i were processed separately using the Ricopili pipeline and imputated using the downloadable version of the Haplotype Reference Consortium (HRC) (accession number: EGAD00001002729)^37^ as reference. Genetic outliers were excluded based on principal component analysis.. A total of 2118 female cases and 19496 female controls as well as 1228 male cases and 20083 male controls were included, respectively. Processing and analysis of genotype data were performed at the secured, national high performance-computing cluster *GenomeDK* (<http://genome.au.dk>). The study was approved by the Danish Data Protection Agency and the Scientific Ethics Committee in Denmark.

**Stefánsson, H | [PGC1 replication] | Iceland (deCODE genetics) | deCODE**

The Icelandic sample consisted of 2,908 subjects with BD (1661 SNP typed) and 344,848 controls (141,854 SNP typed). DNA was isolated from blood samples provided by patients and controls that were recruited throughout Iceland. Approval for the study was granted by the National Bioethics Committee of Iceland and the Icelandic Data Protection Authority and informed consent was obtained for all participants providing a sample for the study. Diagnoses were assigned according to Research Diagnostic Criteria^38^ through the use of the SADS-L^39^ for 303 subjects. DSM-IV BD diagnoses were obtained through the use of the Composite International Diagnostic Interview (CIDI-Auto) for 82 subjects. The remaining BD subjects were diagnosed by ICD 9 or ICD 10 at Landspitali University Hospital in the years 1987-2018. Controls were recruited as a part of various genetic programs at deCODE and were not screened for psychiatric disorders. Whole genome sequencing was performed on samples from 541 BD cases and 26,014 controls. Two types of imputations were performed; into SNP-typed individuals based on long-range phasing, followed by a familial imputation step into un-typed relatives of SNP-typed individuals^40^. Cases of bipolar I disorder were defined using ICD-10 codes 31.1 and 31.2 and ICD-9 codes 296.0 and 296.2. Cases of bipolar II disorder were defined using the ICD-10 code 31.0 in the absence of ICD-10 codes F31.1 and F31.2 and ICD-9 codes 296.0 and 296.2.

**Milani L | 24518929 | Estonia (Estonian Biobank) | EstonianBiobank**

The Estonian Biobank (EstBB) is a population-based cohort of 200,000 participants with a rich variety of phenotypic and health-related information collected for each individual^32^. At recruitment, all participants signed a consent to allow follow-up linkage of their electronic health records (EHR), thereby providing a longitudinal collection of phenotypic information. Health records have been extracted from the national Health Insurance Fund Treatment Bills (from 2004), Tartu University Hospital (from 2008), and North Estonia Medical Center (from 2005). The diagnoses are coded in ICD-10 format and drug dispensing data include drug ATC codes, prescription status and purchase date (if available). For the current study, cases of bipolar disease were determined by searching the EHRs for data on F31* ICD-10 diagnosis. All remaining participants who did not have any ICD-10 F* group diagnoses were defined as controls. Cases with bipolar I disorder were those with ICD codes of F31.1 and F31.2.

**Zwart JA | Unpublished | Norway (the Trøndelag Health Study) | HUNT**

The HUNT sample consisted of 905 subjects with BD and 41,914 population controls^41^. Patients and controls were of European ancestry and were recruited from the Nord-Trøndelag County, Norway. Diagnoses were assigned according to ICD-9 or ICD-10. The controls included individuals not diagnosed with substance use disorders, schizophrenia, bipolar disorder, major depressive disorder, anxiety disorders, eating disorders, personality disorders, or ADHD in hospitals (ICD-9 or ICD-10) or general practice (ICPC2). They also were >40 years of age, had low self-reported levels of anxiety and depression (HADS-A and HADS-D ≤ 11), and reported no use of antidepressants, anxiolytics, or hypnotics. Approval for the study was granted by the Data Inspectorate of Norway, the Health Directorate and the Regional Committee for Medical and Health Research Ethics. Cases of bipolar I disorder were those with ICD codes of F31.1, F31.2 or F31.6 and individuals with an ICD-9 code of 295 or ICD-10 codes F20-F29 were excluded. Cases of bipolar II disorder were those with ICD codes of F31.8 and individuals with an ICD-9 code of 295 or ICD-10 codes F20-F29, F31.1-.2 or F31.6 were excluded.

**Breen G | 30305743 | UK (UK Biobank) | UKBiobank**

The UK Biobank is a prospective cohort study of 501,726 individuals, recruited at 23 centres across the United Kingdom^42^. Extensive phenotypic data are available for UK Biobank participants from health records and questionnaires. Participants were classified as having bipolar disorder if they had a reported clinical diagnosis of bipolar disorder (all primary and secondary ICD10 F31 code diagnoses in hospital inpatient records data; UK Biobank category 2002; <http://biobank.ctsu.ox.ac.uk/showcase/label.cgi?id=2002>; N = 777) or if they self-reported bipolar disorder during an interview with a nurse at baseline recruitment (UK Biobank data-field 20002; <http://biobank.ctsu.ox.ac.uk/showcase/field.cgi?id=20002>; N = 1,116; union N = 1,454). The selection of control participants has been described previously^43^. Control participants did not meet case criteria, did not report the use of any psychiatric medication at baseline (UK Biobank data-field 20003; <http://biobank.ctsu.ox.ac.uk/showcase/field.cgi?id=20003>), and did not self-report any history of mental health disorder in the online mental health questionnaire (UK Biobank category 136; <http://biobank.ctsu.ox.ac.uk/showcase/label.cgi?id=136>; N = 58113).

***========* PGC PsychChip Samples *========***

**Pato, C | Not published | [PGC Psychchip] | gpcw1**

The cases and controls in this study were ascertained in the same manner as those described above for bip_usc2_eur.

**Reif, A | Not published | [PGC Psychchip] | germ1**

Cases were recruited in the same manner as those described above for BOMA-Germany II | bip_bmg2_eur. Control subjects were healthy participants who were recruited from the community of the same region as cases. They were of Caucasian descent and fluent in German. Exclusion criteria were manifest or lifetime DSM-IV axis I disorder, severe medical conditions, intake of psychoactive medication as well as alcohol abuse or abuse of illicit drugs. Absence of DSM-IV axis I disorder was ascertained using the German versions of the Mini International Psychiatric Interview. IQ was above 85 as ascertained by the German version of the Culture Fair Intelligence Test 2^44^. Study protocols were reviewed and approved by the ethical committee of the Medical Faculty of the University of Würzburg. All subjects provided written informed consent.

**Serretti, A, Vieta E, Ribases M | Not published | [PGC Psychchip] | spsp3**

The sample includes 267 BD subjects (Spanish Wave2 Serretti PsychChip QC Summary), of which 180 Spanish and 87 Italian. Spanish sample: 180 subjects were enrolled in a naturalistic cohort study, consecutively admitted to the out-patient Bipolar Disorders Unit, Hospital Clinic, University of Barcelona. This is a systematic cross-sectional analysis deeply described in a previous paper on the same sample investigating rs10997870 SIRT1 gene variant^45^. Inclusion criteria were a diagnosis of Bipolar Disorder (type 1 or 2) according to DSM-IV TR criteria and age of 18 years or older. The study was approved by the local ethical committee and carried out in accordance with the ethical standards laid down in the Declaration of Helsinki. Signed informed consent was obtained from all participants after a detailed and extensive description of the study and patient’s confidentiality was preserved. The current and lifetime diagnoses of mental disorders were formulated by independent senior psychiatrists (diagnostic concordance: Kappa=0.80) according to DSM-IV TR clinical criteria and confirmed through the semi-structured interviews for Axis I disorders according to DSM IV TR criteria (SCID I). Furthermore, all available clinical data coming from follow-up at our unit and collateral information concerning illness history were cross-referred in order to ensure accuracy and obtain complete clinical information. Specific psychopathological dimensions were assessed by means of rating scales and clinical questionnaires administered by clinicians, adequately trained to enhance inter-rater reliability. Mood episodes were defined according to DSM-IV TR criteria and their severity was measured through the administration of the 21-item Hamilton Depression Rating Scale (HDRS-21, Spanish version). The most severe depressive episode was defined on the basis of the severity at the HDRS (total score > 14) and clinical judgment. Italian sample: 87 subjects with bipolar depression were enrolled into the study when admitted at the Department of Psychiatry, University of Bologna, Italy. A description of the subjects has been previously reported when analyzing clinical features^46^. Inclusion criteria were: a diagnosis of bipolar disorder, most recent episode depressive as assessed by DSM-IV-TR criteria; Young Mania Rating Scale (YMRS) score <12; Hamilton Depression Rating Scale (HAM-D) <12. Exclusion criteria were: presence of a bipolar disorder, most recent episode manic or hypomanic; presence of severe medical conditions; presence of moderate to severe dementia (Mini Mental State Examination score <20). The following scales were administered biweekly during the hospitalization: HAM-D, Hamilton Anxiety Rating Scale (HAM-A), YMRS and Dosage Record and Treatment Emergent Symptom Scale (DOTES). Written informed consent was obtained for each patient recruited. The study protocol was approved by the local Ethical Committee and it has been performed in accordance with the ethical standards laid down in the 1975 Declaration of Helsinki.

The Spanish controls were part of the Mental-Cat clinical sample or the INSchool population-based cohort. A total of 1,774 controls from the Mental-Cat cohort (60.5% males) were evaluated and recruited prospectively from a restricted geographic area at the Hospital Universitari Vall d’Hebron of Barcelona (Spain) and consisted of unrelated healthy blood donors. The INSchool sample consisting of 771 children (76.2% males) from schools in Catalonia were involved for screening using the Achenbach System of Empirically Based Assessment (ASEBA) with the Child Behavior Checklist CBCL/4-18 (completed by parents or surrogates), the Teacher Report Form TRF/5-18 (completed by teachers and other school staff) and the Youth Self-Report YSR/11-18 (completed by youths); the Strengths and Difficulties Questionnaire (SDQ) and the Conner’s ADHD Rating Scales (Parents and Teachers). Genomic DNA samples were obtained either from peripheral blood lymphocytes by the salting out procedure or from saliva using the Oragene DNA Self-Collection Kit (DNA Genotek, Kanata, Ontario Canada). DNA concentrations were determined using the Pico- Green dsDNA Quantitation Kit (Molecular Probes, Eugene, OR) and genotyped with the Illumina Infinium PsychArray-24 v1.1 at the Genomics Platform of the Broad Institute. The study was approved by the Clinical Research Ethics Committee (CREC) of Hospital Universitari Vall d'Hebron, all methods were performed in accordance with the relevant guidelines and regulations and written informed consent was obtained from participant parents before inclusion into the study. Detailed information has been published previously^47^.

**Perlis, R; Sklar, P; Smoller, J, Goes F, Mathews CA, Waldman I | Not published | [PGC Psychchip] | usaw4**

Perlis, R; Sklar, P; Smoller, J: EHR data were obtained from a health care system of more than 4.6 million patients^48^ spanning more than 20 years. Experienced clinicians reviewed charts to identify text features and coded data consistent or inconsistent with a diagnosis of bipolar disorder. Natural language processing was used to train a diagnostic algorithm with 95% specificity for classifying bipolar disorder. Filtered coded data were used to derive three additional classification rules for case subjects and one for control subjects. The positive predictive value (PPV) of EHR-based bipolar disorder and subphenotype diagnoses was calculated against diagnoses from direct semistructured interviews of 190 patients by trained clinicians blind to EHR diagnosis. The PPV of bipolar disorder defined by natural language processing was 0.86. Coded classification based on strict filtering achieved a value of 0.84, but classifications based on less stringent criteria performed less well. No EHR-classified control subject received a diagnosis of bipolar disorder on the basis of direct interview (PPV=1.0). For most subphenotypes, PPV exceeded 0.80. The EHR-based classifications were used to accrue bipolar disorder cases and controls for genetic analyses. Samples were genotyped on the Psychchip array.

Goes, FS: Cases represented independent probands from a European-American family sample that was collected at Johns Hopkins University from 1988-2010. Families had at least 2 additional relatives with a major mood disorder (defined as bipolar disorder type 1, bipolar type 2 or recurrent major depressive disorder). Diagnostic interviews were performed using the Schedule for Affective Disorders and Schizophrenia-Lifetime Version (N=81) and the Diagnostic Instrument for Genetics Studies (N=161). All cases underwent best-estimate diagnostic procedures. After genotyping quality control there were 242 cases, of which 240 were diagnosed as Bipolar Disorder type 1 and 2 as Schizoaffective Disorder, bipolar type. Diagnoses were based on DSM-III and DSM-IV criteria. Probands from this sample have been previously studied in family based linkage and exome studies.^49–51^

Mathews CA: Control samples were ascertained as part of ongoing genetic and neurophysiological studies of hoarding, obsessive compulsive and tic disorders. Controls reported no current or lifetime history of mania or hypomania at the time of ascertainment. Sixty-two of the 104 controls were screened for psychiatric illness using the Structured Clinical Interview for DSM-IV TR diagnoses and diagnoses of bipolar disorder, lifetime or current, were ruled out through a best estimate consensus diagnosis. Other psychiatric diagnoses were not excluded. The remaining 42 participants were not formally screened, but reported no lifetime or current history of bipolar disorder, obsessive compulsive, hoarding, or tic disorders. Samples were genotyped on the Psychchip array. Ethical approvals were obtained from the University of Florida Human Subjects Review Board.

Waldman I: Control samples were ascertained as part of an ongoing genetic study of ADHD and other Externalizing disorders (I.e., Oppositional Defiant Disorder and Conduct Disorder). Controls reported no current diagnoses of Externalizing or Internalizing disorders at the time of ascertainment. Controls were assessed for psychiatric conditions using the Emory Diagnostic Rating Scale (EDRS)^52^, a questionnaire that assessed parent ratings of symptoms of common DSM-IV Externalizing and Internalizing disorders (e.g., Major Depressive Disorder and various anxiety disorders). Samples were genotyped on the Psychchip array. Ethical approvals were obtained from the Emory University and University of Arizona Human Subjects Review Boards.

**Baune, BT; Dannlowski, U | Not published | [PGC Psychchip] | bdtrs**

The Bipolar Disorder treatment response Study (BP-TRS) comprises BD inpatient cases and screened controls of Caucasian background. Psychiatric diagnosis of Bipolar Disorders was ascertained using SCID or MINI 6.0 using DSM-IV criteria in a face-to-face interview by a trained psychologist / psychiatrist for both cases and controls. Healthy controls were included if no current or lifetime psychiatric diagnosis was identified. Cases were included if current or lifetime diagnosis of bipolar disorder was ascertained by structured diagnostic interview. Cases and controls are of similar age range (>=18 yrs of age) and were collected from the same geographical areas. Other assessments including symptom ratings, psychiatric history, treatment history, treatment response were based on interview, and carried out by trained psychologists/psychiatrists. Samples were genotyped on the Psychchip array. Ethical approval was obtained from the University of Münster Human Ethics Committee, Münster, Germany.

**Ophoff R, Posthuma D, Lochner C, Franke B | Not published | [PGC Psychchip] | dutch**

Ophoff R: Cases and controls were collected using the same protocol as described above for the “ucla” sample.

Lochner C: Controls include South African Caucasian population based-controls ascertained from blood banks and controls recruited through university campuses and newspaper advertisements, who underwent a psychiatric interview and had no current or lifetime psychiatric disorder^53^^,^^54^.

Franke B: The controls included are healthy individuals from the Dutch part of the International Multicenter ADHD Genetics (IMAGE) project^55^^,^^56^.

Posthuma D: Data were provided for 960 unscreened Dutch population controls from the Netherlands Study of Cognition, Environment and Genes (NESCOG)^57^. The study was approved by the institutional review board of Vrije Universiteit Amsterdam and participants provided informed consent.

**Gawlik M | Not published | [PGC Psychchip] | gawli**

Patients were recruited at the Department of Psychiatry, Psychosomatics and Psychotherapy, University of Würzburg, Germany. Diagnosis according to DSM-IV (Diagnostic and Statistical Manual of Mental Disorders-fourth edition) was made by the best estimate lifetime diagnosis method, based on all available information, including medical records, and the family history method.

**Fullerton J, Mitchell PB, Schofield PR, Green MJ, Weickert CS, Weickert TW, The Australian Schizophrenia Research Bank | Not published | [PGC Psychchip] | neuc1**

The NeuRA collection comprised BD cases from three cohorts ascertained in Australia: the bipolar high risk study^58^ (n=97), the Imaging Genetics in Psychosis Study (IGP; n=47)^59^ and a clinic sample (n=109) recruited via the Sydney Bipolar Disorders Clinic^60^. The clinic sample used the same ascertainment procedures as described for the bip_bmau_eur sample. The bipolar high risk study is a collaborative study with 4 US and one Australian groups, with young participants aged 12-30. The IGP sample was recruited from outpatient services of the South Eastern Sydney-Illawarra Area Health Service (SESIAHS), the Sydney Bipolar Disorders Clinic and the Australian Schizophrenia Research Bank. Healthy controls were sourced from the high risk, IGP and the Cognitive and Affective Symptoms of Schizophrenia Intervention (CASSI) trial^61^ studies, and were recruited from the community, had no personal lifetime history of a DSM-IV Axis-I diagnosis as determined by psychiatric interview, and no history of psychotic disorders among first-degree biological relatives. Additional controls were recruited as part of the strategy to develop an Australian Schizophrenia Research Biobank for studies into the genetics of this disease. The ascertainment of these controls has been previously described^62^.

**Landen M, Hillert J, Alfredsson L | Not published | [PGC Psychchip] |** **swed1**

The cases in the swed1 sample were recruited using the same ascertainment methods described for the bip_swa2_eur sample. Population-based healthy controls, randomly selected from the Swedish national population register, were collected as part of two case-control studies of multiple sclerosis: GEMS (Genes and Environment in Multiple Sclerosis) and EIMS (Epidemiological Investigation of Multiple Sclerosis)^63^.

**Di Florio A, McQuillin A, McIntosh A, Breen G | Not published | [PGC Psychchip] | ukwa1**

McQuillin A: BD cases were recruited using the same protocol as the bip_uclo_eur described above. A subset (n=448) of the control subjects were random UK blood donors obtained from the ECACC DNA Panels (<https://www.phe-culturecollections.org.uk/products/dna/hrcdna/hrcdna.jsp>). The remaining control subjects (n=814) had been screened for an absence of mental illness in using the same protocol as the bip_uclo_eur described above.

Di Florio A: Cases were recruited across the United Kingdom in the same manner as described for the bip_wtcc_eur and bip_icuk_eur samples.

McIntosh AM: BD cases were recruited from the clinical case loads of treating psychiatrists from Edinburgh and across the central belt of Scotland. Controls were identified from non-genetic family members and from the extended networks of the participants themselves. All participants were of European ancestry and diagnosis was confirmed using an established battery developed for ICCCBD. Breen G: Controls were drawn from blood donors to the UK Motor Neuron Disease Association DNA Biobank^64^

**Perlis, R; Sklar, P; Smoller, J, Nievergelt C, Kelsoe J | Not published | [PGC Psychchip] | usaw5**

Kelsoe, J: The Pharmacogenomics of Bipolar Disorder (PGBD) study was a prospective assessment of lithium response in BDI patients. The goal was to identify genes for lithium response. Subjects were recruited from clinics at 11 international sites and followed for up to 2.5 years. Diagnosis was obtained by DIGS interview and medical records reviewed by blind experienced clinicians. As the comparison was between lithium responders and non-responders, no controls were collected. All subjects provided written informed consent.

Perlis R: Cases of bipolar disorder were Individuals treated with lithium drawn from the Partners Healthcare electronic health record (EHR) database, which spans two large academic medical centers, Massachusetts General Hospital and Brigham and Women’s Hospital in addition to community and specialty outpatient clinics^65^. Any patients aged 18 years or older with at least one lithium prescription between 2006 and 2013 based on e-prescribing data were included. The Partners Institutional Review Board approved all aspects of this study. Individuals with a diagnosis of schizophrenia based on ICD9 codes were excluded.

Smoller J: Cases and controls were recruited in the same manner as described above for “usaw4”.

***========* PGC3 Samples *========***

**Rietschel M, Nöthen MM, Forstner AJ, Streit F, Babadjanova G |24618891| Russia (BOMA-Russia) | bmrus**

Patients were recruited from consecutive admissions to the psychiatric inpatient units of the Russian State Medical University, Moscow. Unrelated controls were recruited from the general population. All protocols and procedures were approved by the respective local Ethics Committees. Written informed consent was obtained from all study participants before the study participation. All patients were assigned a lifetime diagnosis of BPAD type I or type II. This was based on Diagnostic and Statistical Manual of Mental Disorders-IV criteria and a consensus best-estimate procedure, including a structured interview-I, review of medical records, the family history method and the Operational Criteria Checklist for Psychotic Illness OPCRIT system.

**Ferentinos P, Dikeos D, Patrinos G | Not published | Greece (Attikon General Hospital) | greek**

All adult patients with a DSM-IV-TR/DSM-5 diagnosis of Bipolar Disorder hospitalized at the inpatient unit or followed-up at the specialized ‘Affective disorders and Suicide’ outpatient clinic of the 2nd Department of Psychiatry, National and Kapodistrian University of Athens, Attikon General Hospital, Athens, Greece from 2012 to 2017 were recruited for the current study. Patients were referred to the specialized ‘Affective disorders and Suicide’ outpatient clinic either from the inpatient unit after hospitalization or from the community. Diagnosis was established and demographic (age, gender, family status, profession, employment status, education) and relevant clinical features (e.g. age at onset, polarity of first and most recent episode, number of lifetime depressive and manic/hypomanic episodes, number of hospitalizations, lifetime suicidality, lifetime psychosis) were extracted through a M.I.N.I.-5.0.0-based semi-structured diagnostic interview, which was administered during patients’ initial clinical assessment and regularly updated ever since, interviews of primary caregivers and inspection of medical records. Lifetime presence of any DSM-IV-TR axis I psychiatric comorbidities (dysthymia, panic disorder, agoraphobia, social phobia, generalized anxiety disorder, obsessive-compulsive disorder, post-traumatic stress disorder, alcohol and substance abuse and dependence, anorexia nervosa, bulimia nervosa) was similarly extracted. Family history of major psychiatric disorders and suicidality in first and second degree relatives was recorded with a specific questionnaire based on the Family Interview for Genetic Studies. Medical comorbidities were recorded with the Cumulative Illness Rating Scale, completed on the basis of interview with patient and primary caregivers, inspection of patient's medical records and laboratory exams (basic or specific, if considered necessary). Presence of selected medical diseases was specifically recorded.

Control (unaffected) participants were a convenient sample drawn from the same geographic area as case participants, either within health care facilities or as community volunteers. All of them went through a brief clinical interview including items on psychiatric and medical history, psychiatric family history, past and current medical or psychiatric therapies, and a brief mental state examination. Only participants found to be free of lifetime major mental disorders (MDD, BD, schizophrenia, or other psychotic disorders) and with no family history of major mental disorder in their first-degree relatives were recruited as controls.

All cases and controls were native Greek speakers. All participants provided written informed consent before being included in the study and the study protocol was approved by the Research Ethics Committee of Attikon General Hospital.

**Andreassen, OA | Not published | Norway (TOP) | norgs**

The NORGS bipolar disorder cases and controls were ascertained in the same way as the bip_top7_eur (TOP7) samples described above, and recruited from hospitals across Norway.

**Andreassen, OA | Not published | Norway (TOP) | noroe**

The NOROE bipolar disorder cases and controls were ascertained in the same way as the bip_top7_eur (TOP7) samples described above, and recruited from hospitals across Norway.

**Reininghaus EZ | Not published| Austria (Medical University of Graz) | graza**

Univ. Prof. DDr. Eva Reininghaus, Priv.Doz. DDr. Susanne Bengesser, Priv.Doz. Dr. Nina Dalkner, Dr. Frederike Fellendorf and further team members of the special outpatients department for bipolar affective disorders at the Department of Psychiatry and Psychotherapeutic Medicine, Medical University of Graz, Austria: Cases with bipolar affective disorder (type I and II) and healthy controls were recruited at the Department of Psychiatry and Psychotherapeutic Medicine at the Medical University of Graz (MUG), Austria. Study protocols were approved by the ethics committee of the Medical University of Graz. Patients and healthy controls gave written informed consent and the study was conducted according to the declaration of Helsinki. All patients received a clinical interview by a psychiatrist or psychologist and a diagnosis according to DSM-IV with the SCID-I (Structured clinical interview). Healthy controls did not have a history of a psychiatric disorder. Furthermore, healthy controls did not have any first or second degree relatives with a psychiatric disorder. The PGC-Graz sample (n= 244; 114 males, 130 females) includes 167 cases with bipolar disorder and 77 healthy controls genotyped with Omniexpress 1.2 by Illumina.

**Grigoroiu-Serbanescu M | 31791676; 26806518 | Romania (BOMA-Romania) | bmrom**

This sample includes the BOMA-Romania sample and additional cases from the ConLiGen-Romania sample. For the BOMA-Romania sample, unrelated BP-I patients were recruited from consecutive admissions in the Obregia Psychiatric Hospital of Bucharest, Romania. All participants provided written informed consent following a detailed explanation of the study aims and procedures. The study was performed in accordance with the Code of Ethics of the World Medical Association (Declaration of Helsinki). All participants were of Romanian descent according to self-reported ancestry. Genealogical information about parents and all four grandparents was obtained through direct interview of the subjects.

The patients were investigated with the Diagnostic Interview for Genetic Studies (DIGS)^28^ and the Family Interview for Genetic Studies (FIGS)^8^ The diagnosis of BP-I was assigned according to DSM-IV criteria on the basis of both the DIGS and medical records. Patients were included in the sample if they had at least two documented hospitalized illness episodes (one manic/mixed and one depressive or two manic episodes) and no residual mood incongruent psychotic symptoms during remissions. This information was also confirmed by first degree relatives for 64% of the cases. The illness age-of-onset was defined as the age at which the proband first met DSM-IV criteria for a manic, mixed, or major depressive episode. Family history of psychiatric illness was obtained with FIGS administered both to the patients and to all available relatives.

Cases in the ConLiGen-Romania study were ascertained in the same manner as for BOMA-Romania. Cases were required to have taken lithium for at least two years and lithium treatment response was evaluated with the Alda scale^66^**.**

Population-based controls were evaluated using the DIGS and FIGS to screen for a lifetime history of major affective disorders, schizoaffective disorders, SCZ and other psychoses, obsessive-compulsive disorder, eating disorders, and alcohol or drug addiction. Unaffected individuals were included as controls in the present study.

***========* PGC4 Samples *========***

**Grigoroiu-Serbanescu M | PMID : 31791676| Romania (BOMA-Romania) | rom4**

Cases were recruited from consecutive admissions to the Obregia Clinical Psychiatric Hospital, Bucharest, Romania. Patients were administered the DIGS^28^ and FIGS^8^ interviews. Information was also obtained from medical records and close relatives. The diagnosis of BP-I was assigned according to DSM-IV-R criteria using the best estimate procedure. All patients had at least two hospitalized illness episodes. Population-based controls were evaluated using the DIGS to exclude a lifetime history of major affective disorders, schizophrenia, schizoaffective disorders, and other psychoses, obsessive-compulsive disorder, eating disorders, and alcohol or drug addiction.

**McQuillin A | PMID: 37643680 | UCL (University College London), London, UK | amq1**

Case and controls were collected using the protocol described above for bip_uclo_eur.

**Squassina A, | PMID: 21961650 | Italy | ital1**

Patients with bipolar I or bipolar II disorder were recruited at the outpatient unit (Lithium Clinic) of the Clinical Psychopharmacology Centre at the Department of Biomedical Science, Section of Neuroscience & Clinical Pharmacology, University of Cagliari, University Hospital Agency of Cagliari, Italy. Clinical assessments followed a strict procedure. After providing informed consent, participants were interviewed using one of the structured or semistructured interviews SADS-L. Clinical diagnosis was confirmed by DSM-IV criteria. We also used available medical records, narrative summaries of all interviews, and details such as baseline assessments, clinical course, response to treatment, treatment adherence, psychiatric and medical comorbidities, history of suicidal behavior, and symptom profiles in OPCRIT format.^6^

For uniform evaluation of treatment response, we used all available information including data from clinical records, diagnostic interviews, and prospective follow-up assessed by NIMH Life- Chart Method^67^. We used the Alda scale to assess lithium response^66^.

**Manchia M, Carpiniello B, Squassina A | PMID: 35566641 | Italy | ital2**

The case samples were recruited among patients attending the outpatient clinic of the community mental health center of the Unit of Clinical Psychiatry within the University Hospital of Cagliari, Italy. Patients were enrolled in the genetic study if they met the following inclusion criteria: diagnosis of either Bipolar I or Bipolar II disorder according to DSM 5^68^ criteria validated through the Italian version of the SCID-5-CV (Structured Clinical Interview for DSM-5 Clinical Version); being in euthymic phase.
All patients provided a written consent form regarding the use of their biological and clinical data for research purposes. Blood samples were gathered at the beginning of the study along with the relevant demographic and biometric data. All the clinical documents are stored in an anonymized database, accessible only by authorized personnel.

The recruited subjects were phenotypically characterized with the use of the following standardized tests:

·	Brief Assessment of Cognition in Affective Disorders (BACA)

·	Brief Assessment of Cognition in Schizophrenia to assess baseline cognitive capacities

·	Hamilton Depression Rating Scale (HDRS)

·	Young Mania Rating Scale (YMRS)

·	Hamilton Anxiety Rating Scale (HAM-A)

·	Barratt Impulsivity scale (BIS)

·	Clinical Global Impression Scale – Severity (CGI-S)

·	Alda score for Lithium response (clinical response defined as a score >7)

·	OPCRIT

**Tondo L, Squassina A | PMID: 20348464 | Italy | ital3**

Our sample population encompasses a cohort of patients followed at the Mood Disorder Lucio Bini Center in Cagliari (Italy), a specialized outpatient clinic for the diagnosis, treatment and research of affective disorders. Since the founding of this outpatient clinic in 1977, all demographic and clinical information about patients have been recorded systematically by means of semi-structured initial and follow-up interviews, a life chart, extensive clinical evaluation and repeated assessments with standard rating scales for mood such as the Hamilton Depression Rating Scale (HDRS)^69^, and Young Mania Rating Scale^70^, typically every 4–6 weeks. Diagnoses were updated to meet the Diagnostic and Statistical Manual of Mental Disorders (DSM)-5 criteria^68^ after the year 2013. Written informed consent was obtained for collection and analysis of patient data to be presented anonymously in aggregate form, in accordance with the requirements of Italian law and following review by a local ethical committee. Required data were entered into a computerized database in coded form to protect subject identity.

Patients were included in the study if they had at least 12 months of treatment with lithium and if they had a diagnosis of bipolar disorder (BD) or major depressive disorder (MDD) according to DSM-5. The clinical response to lithium treatment was characterized using the "Retrospective Criteria of Long-Term Treatment Response in Research Subjects with Bipolar Disorder" scale, also known as Alda Scale^66^.

**Kircher T, Dannlowski U | PMID 30267149| Germany | FOR 2107**

FOR 2107 is a longitudinal cohort study aiming to integrate clinical and neurobiological associations of genetic and environmental risk factors and their interaction involved in the aetiology, onset and course of affective disorders^71^. Participants of the present study were part of the bi-center “Marburg Münster Affective Cohort Study” (MACS) and were recruited from in- and out-patient departments of the universities of Marburg and Münster, Germany, local psychiatric hospitals (Vitos Marburg, Gießen, Herborn, and Haina, LWL Münster, Germany), and via postings in local newspapers and flyers.

Lifetime BD was assessed using a semi-structured interview according to DSM-IV-TR (Diagnostic and Statistical Manual of Mental Disorders)^72^ applied by trained staff and according to detailed SOPs. Controls did not report any lifetime history of psychiatric diagnosis or treatment as determined in line with DSM-IV-TR. All raters underwent standardized training, and interviews were regularly supervised and videotaped.

All procedures were approved by the local Ethics Committees of Marburg (AZ:07/14) and Münster (AZ:2014-422-b-S), Germany, according to the Declaration of Helsinki. Participants gave written informed consent prior to study participation and received financial compensation.

The study is funded by the German Research Foundation (Research Unit FOR 2107). Principal investigators are Tilo Kircher (KI588/14-1, KI588/14-2, KI588/20-1, KI588/22-1), Udo Dannlowski (DA 1151/5-1, DA 1151/5-2, DA1151/6-1), Axel Krug (KR3822/5-1, KR3822/7-2), Igor Nenadic (NE2254/1-2,NE2254/3-1,NE2254/4-1), Carsten Konrad (KO4291/3-1), Marcella Rietschel (RI 908/11-1, RI 908/11-2), Markus Nöthen (NO 246/10-1, NO 246/10-2), Stephanie Witt (WI 3439/3-1, WI 3439/3-2). We are deeply indebted to all study participants and staff. A list of acknowledgments can be found here: www.for2107.de/acknowledgements. Tilo Kircher received unrestricted educational grants from Servier, Janssen, Recordati, Aristo, Otsuka, neuraxpharm. Further information on the FOR 2107 data used for this article can be found here: www.for2107.de. Qualified researchers can request access to FOR 2107 data through contacting the principal investigators.

**Alda M | Not published | Nova Scotia, Canada | hal3**

The case samples were recruited from patients longitudinally followed at a specialty mood disorders clinic in Halifax (Canada). Cases were interviewed in a blind fashion with the Schedule of Affective Disorders and Schizophrenia-Lifetime version (SADS-L)^13^ by pairs of clinician researchers (psychiatrists and/or nurses). The interviews together with medical records were subsequently reviewed in a blind fashion by a panel of senior clinical researchers. Consensus diagnoses were made according to DSM-IV^14^ and Research Diagnostic Criteria (RDC)^15^ Protocols and procedures were approved by the local Ethics Committees and written informed consent was obtained from all patients before participation in the study.

***========* External Samples PGC4 *========***

**Iwata N | PMID: 28115744 | Japan (advanced COSMO and Biobank Japan)**

A detailed description of the sample information, genotyping, quality control and imputation procedures is reported elsewhere^73^. The diagnosis for each case subject followed the DSM-IV-TR criteria for BD and schizoaffective disorder and was reached by the consensus of at least two experienced psychiatrists, based on unstructured interviews with the subject and their family, as well as a review of the subject's medical records. For the comparison subjects, we used GWAS data for subjects in the BioBank Japan project collected as case subjects for non-psychiatric disorders. These subjects were not psychiatrically evaluated.

**Hong-Hee Won, Woojae Myung, Heon-Jeong Lee, Genoplan Research Team | Not published | South Korea**

We recruited 201 and 347 patients with bipolar disorder from Seoul National University Bundang Hospital (SNUBH) and Korea University College of Medicine, respectively. We included 5,462 individuals with no history of mental illness in the community-based cohort as controls. DNA samples were genotyped using the Illumina Asian Screening Array or the Affymetrix Axiom Korea Biobank Array 1.0. We checked data quality at sample level and at variant level prior to analysis. For quality control (QC), we excluded duplicate variants or variants with missingness >1%, minor allele frequency <1%, or Hardy-Weinberg equilibrium P <1e-06. After variant level QC, we excluded one of genetically related samples, samples with missingness >5%, heterozygosity rate beyond 5 standard deviations, or sex mismatch. After removing these samples from the raw genotype data, we conducted variant level QC again in the same way as above. Genotype imputation was performed using Haplotype Reference Consortium (HRC) reference data on the Michigan Imputation Server, and we removed imputed variants with R2 <0.8 and minor allele frequency <1%. After the QC process, a total of 6,002 individuals (541 cases and 5,461 controls) and 4,851,098 variants remained. We performed a genome-wide association analysis adjusted for sex and 10 PCs using the firth model of the PLINK software.

Additional 355 patients with bipolar disorder from SNUBH and 3,550 controls were used in the PRS analysis. Quality control (QC) measures were stringently applied to these samples. Using Plink, we excluded variants with missingness > 0.05, a Hardy-Weinberg equilibrium p-value < 10^-6^, or minor allele frequency (MAF) < 0.01. We excluded samples with missingness > 0.05 or outliers out of three standard deviations from the mean for the first and second principal components of genetic ancestry. We also excluded six samples that were out of three standard deviations from the mean of the heterozygosity rate. Prior to imputation, quality checks were conducted for strand, alleles, position, and reference and alternative allele assignments, as well as frequency differences, utilizing tools provided by the McCarthy group (www.well.ox.ac.uk/~wrayner/tools/). The 1000 Genomes Project Phase 3 data was used as an imputation reference panel. Phasing was conducted with Eagle version 2.4.1, and imputation was performed using Minimac 4. After imputation, we excluded variants with an imputation R^2^ value < 0.3. Additional QC was performed with the same stringent criteria applied in the pre-imputation phase, including genotype missingness, sample missingness, Hardy-Weinberg equilibrium test, and MAF filtering. Ethical approval for this study was obtained from the Institutional Review Board (IRB) of Bundang Seoul National University Hospital (Approval No. X-2301-807-903 and B-2407-912-109). This study was conducted with bioresources from National Biobank of Korea, the Korea Disease Control and Prevention Agency, Republic of Korea (KBN-2021-030).

**Po-Hsiu Kuo, Hsi-Chung Chen |PMID 35321763, 27450446, and 23653667 | Taiwan**

The samples were from the GREAT cohort (Genomic Research and Epidemiology Studies of Affective Disorders in Taiwan), which has been collecting data on mood disorder patients and control groups since 2008. This cohort included a total of 1228 patients with bipolar disorder (comprising both type I and type II) who had genotyping information available, and their ages ranged from 18 to 70 years old. These patients were referred consecutively by psychiatrists from various collaborating hospitals in Taiwan, based on the diagnostic criteria outlined in DSM-IV-TR. Approximately 80% of the patients had undergone detailed interviews, utilizing tools such as the Composite International Diagnostic Interview or the Schedule for Affective Disorders and Schizophrenia questionnaire. DNA samples were genotyped using several arrays, including the Illumina Human Omni Express Beadchip, Illumina Human Omni Express Exome Beadchip, Affymetrix Axiom Genome-Wide CHB Array, and Affymetrix Axiom Genome-Wide TWB 2.0 Array. Imputation was performed using the TopMed imputation server, and quality control measures were implemented, excluding data with a genotype missing rate above 5%, a Hardy-Weinberg equilibrium p-value less than 1e-06, and a minor allele frequency below 1%. More detailed information on the imputation and quality control procedures utilized in our earlier studies, please refer to the following reference (PMID: 27450446).

**GAIN (admixed African American) (USA)**

Genetic Association Information Network (GAIN)/ The Bipolar Genome Study (BiGS) Data from the existing National Institutes of Health Genetic Association Information Network (GAIN) study of bipolar disorder was obtained through dbGap: phs000017.v3.p1. The GAIN study was multi-site and informed consent and institutional review board approval were obtained and details are described above for the GAIN-European data. Bipolar I diagnosis was confirmed with the structured Diagnostic Interview for Genetic Studies (DIGS) for the assessment of major mood and psychotic disorders and their spectrum conditions. The admixed African American (AA) bipolar patient data used in this study were from unrelated individuals in multiplex families and assessed with DIGS version 4. The genotyping has been described previously^74^.

**Andreassen OA | Norwegian Mother, Father and Child Cohort Study (MoBa) (Norway) | 27063603**

MoBa is a longitudinal prospective pregnancy cohort including approximately 114,500 children, 95,200 mothers, and 75,200 fathers^75,76^. Between 1999 and 2008 pregnant women were recruited to the study and gave written informed consent to participation in 41% of the pregnancies. Blood samples were obtained from both parents during pregnancy and from mothers and children (umbilical cord) at birth^77^. The establishment of MoBa and initial data collection was based on a license from the Norwegian Data Protection Agency and approval from The Regional Committees for Medical and Health Research Ethics. MoBa is currently regulated by the Norwegian Health Registry Act (14140 and 2016/1226). We extracted diagnostic outcomes from the Norwegian Patient Registry which contains information on diagnoses from the International Classification of Diseases, Tenth Revision (ICD-10) on in-and outpatients reported from all hospitals and specialized health care services in Norway from 2008-2019^78^. Cases of bipolar disorder were those with F31 ICD codes. The genotyping and quality control has been described previously^79^.

**FINNGEN (Finland) | 36653562**

The FinnGen study (<https://www.finngen.fi/en>) is an ongoing research project that utilizes samples from a nationwide network of Finnish biobanks and digital healthcare data from national health registers. The definitions of FinnGen disease end points and their respective controls for each release are available at https://www.finngen.fi/en/researchers/clinical-endpoints, and FinnGen end points can also be browsed at <https://risteys.finregistry.fi/>. Inclusion and exclusion criteria for individuals with bipolar disorder, and corresponding controls, can be found at <https://risteys.finregistry.fi/endpoints/F5_BIPO>.

**Medland SE | 37655588 | Australian Genetics of Bipolar Disorder Study (GBP)**

**Whiteman DC | 22933644 | QSkin Sun and Health Study (QSkin)**

Cases were recruited from the Australian Genetics of Bipolar Disorder Study (GBP). A detailed description of the GBP study recruitment, self-report online survey and bipolar disorder diagnosis is reported elsewhere^80^. Briefly, bipolar disorder diagnoses were determined using the Mood Disorder Questionnaire (MDQ) and additional items corresponding to DSM-5 BD diagnostic criteria or self-reported bipolar disorder BD diagnosis when MDQ/DSM-5 data were not available. Unrelated controls were recruited from the general population in Queensland, Australia by the QSkin Sun & Health Study^81^. QSkin participants who did not report lifetime diagnoses for any of the following mental health conditions were included as controls: anxiety, depression, postnatal depression, bipolar disorder, schizophrenia, attention-deficit/hyperactivity disorder, autism, anorexia, bulimia and obsessive-compulsive disorder. All protocols and procedures were approved by the respective local Ethics Committees. Written informed consent was obtained from all study participants before the study participation.

**Department of Veterans Affairs Cooperative Studies Program (CSP) (USA) | 33169155**

VA-Cooperative Studies Program (CSP) #572 is a cohort of approximately 9300 veterans with schizophrenia or bipolar 1 disorder who received detailed in-person assessments of clinical diagnosis using the Structured Clinical Interview for DSM (SCID), assessments of performance-based neurocognition and everyday functioning, and suicidality. CSP #572 is a companion study to the Million Veteran Program (MVP), and was genotyped together with MVP on a custom Axiom Biobank array. Control participants were drawn from the MVP roster, excluding participants with lifetime or self-reported history of schizophrenia, bipolar disorder, or depression, or treatment with antipsychotics, mood stabilizers, or antidepressants.

**Genomic Psychiatry Cohort (GPC) (USA) | 33169155**

Details of ascertainment and diagnosis, genotyping and quality control have been described in detail previously^82^. Briefly, cases were ascertained using the Diagnostic Interview for Psychosis and Affective Disorders (DI-PAD), a semi-structured clinical interview administered by mental health professionals, which was developed specifically for the GPC study. Individuals reporting no lifetime symptoms indicative of psychosis or mania and who have no first-degree relatives with these symptoms are included as control participants.

**23andMe Inc (USA)**

23andMe is a company providing direct-to-consumer genetic testing as a paid service to its customers. 23andMe customers are invited to answer research questionnaires and, subject to participants’ informed consent, their health data and genotyping results are included in genetic analyses. Subject to agreements between 23andMe and the PGC, summary findings of genetic analyses were shared with the PGC for inclusion in the current GWAS meta-analysis. The full GWAS summary statistics for the 23andMe discovery data set will be made available through 23andMe to qualified researchers under an agreement with 23andMe that protects the privacy of the 23andMe participants. Datasets will be made available at no cost for academic use. Please visit <https://research.23andme.com/collaborate/#dataset-access/> for more information and to apply to access the data. Case Ascertainment: Phenotypic status was based on responses to web-based surveys, with individuals that self-reported as having received a clinical diagnosis or treatment for bipolar disorder classified as cases. Control Ascertainment: Control status was based on responses to web-based surveys, and individuals that didn’t self-report as having received a clinical diagnosis or treatment for bipolar disorder were classified as controls.
