## Supplementary Figures for "Genomics yields biological and phenotypic insights into bipolar disorder"

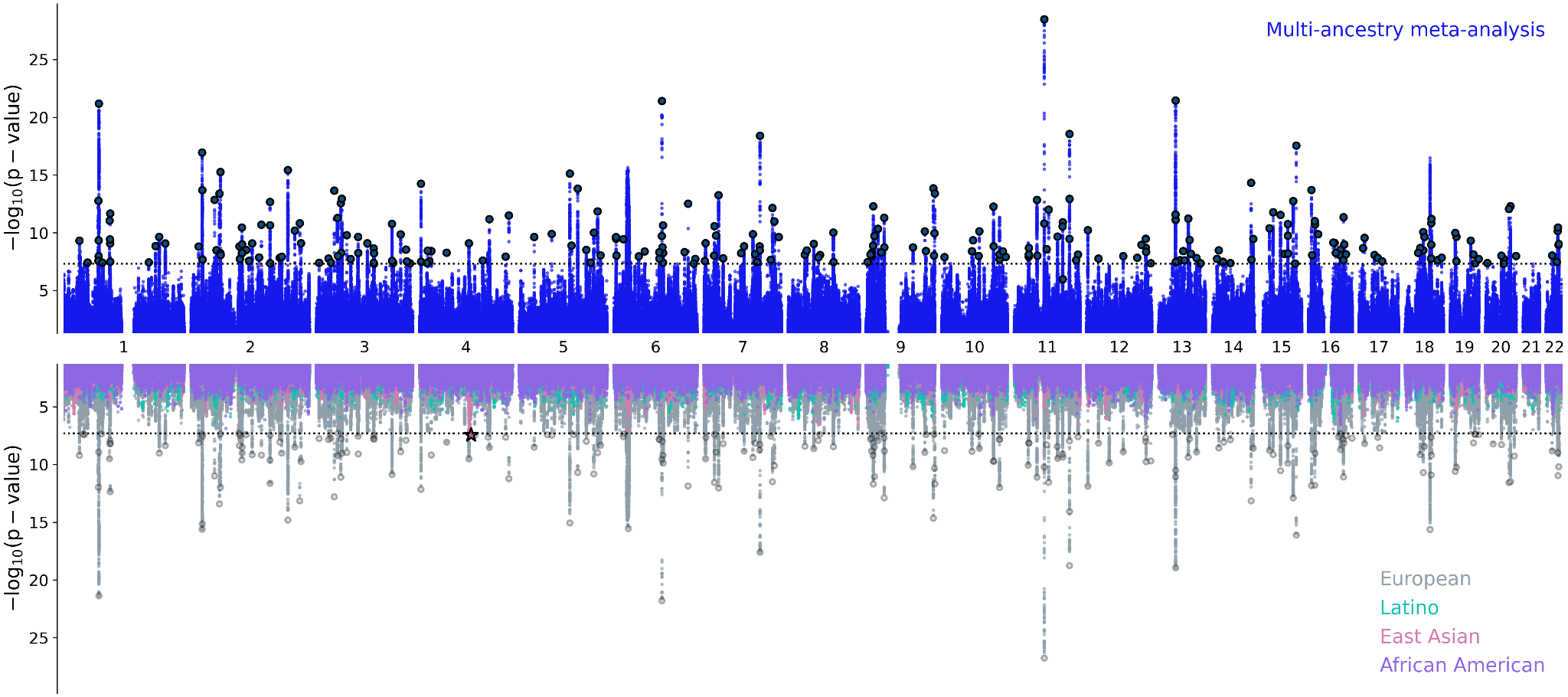

**Supplementary Figure 1. Miami plot for BD genome-wide meta-analyses, including all cohorts.** Upper panel: the multi-ancestry meta-analysis identified 298 genome-wide significant (GWS) loci. Lower panel: porcupine plot showing the results of the Latino (0 GWS loci), African American (0 GWS loci), East Asian (1 GWS locus) and European (229 GWS loci) meta-analyses. The x-axes show genomic position (chromosomes 1–22), and the y axes show statistical significance as –log10[p-value]. P-values are two-sided and based on an inverse-variance-weighted fixed-effects meta-analysis. The dashed black lines show the GWS threshold (P < 5 × 10^−8^). The star indicates the position of the East Asian GWS locus (rs117130410, 4:105734758, build GRCh37).

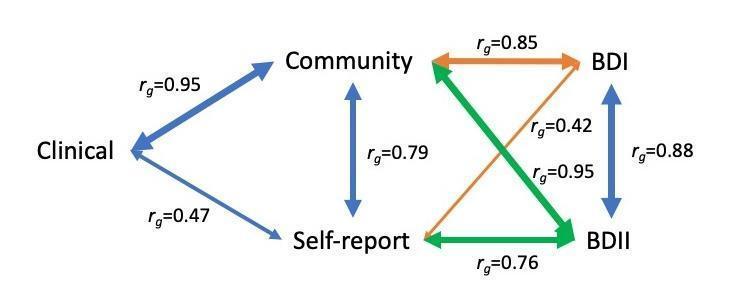

**Supplementary Figure 2. Network diagram of the genetic correlations between BD ascertained from Clinical, Community and Self-report samples, as well as BD-subtypes (BDI and BDII).**

The line widths are proportional to the strength of the correlations between pairs. BDI: bipolar disorder I, BDII: bipolar disorder II

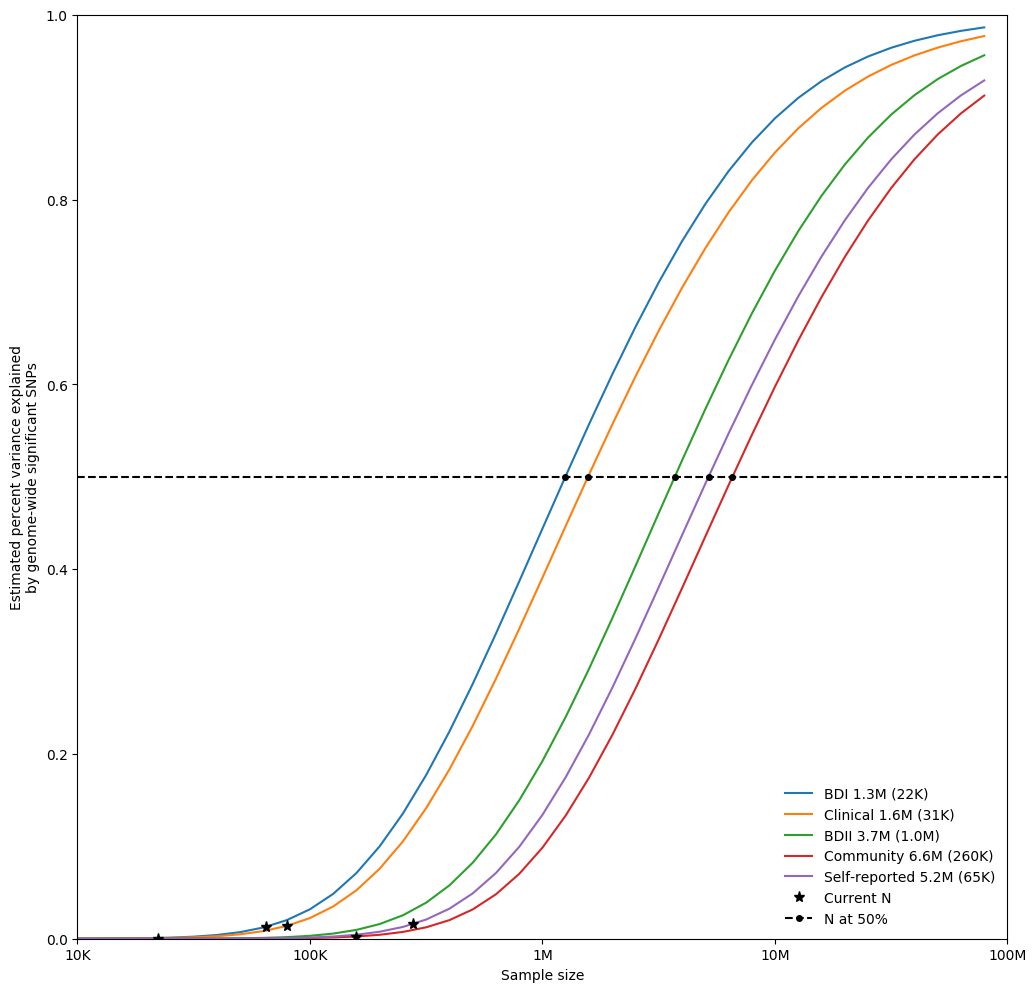

**Supplementary Figure 3: Univariate MiXeR estimates of the required effective sample size needed to capture 50% of the genetic variance (horizontal dashed line) associated with each BD ascertainment and subtype.**

N and Sample size refer to the effective sample size. The estimated effective sample size (and standard errors) are given in the legend alongside each trait name.

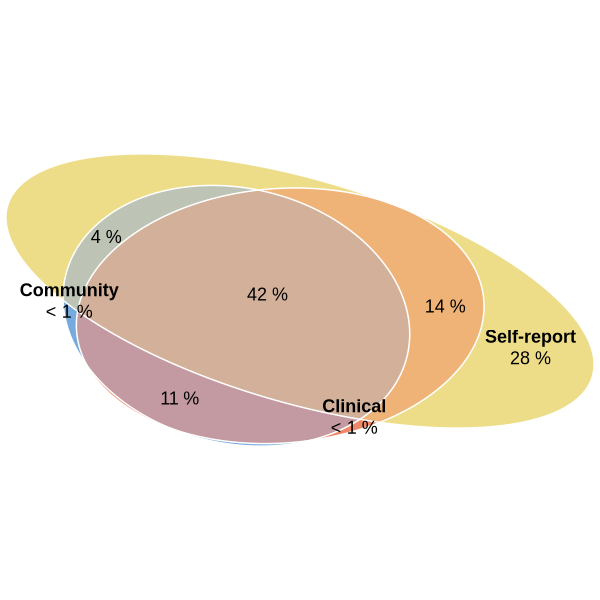

**Supplementary Figure 4. Trivariate MiXeR estimates for the genetic overlap of BD from Clinical, Community and Self-report samples.**

The percentages show the proportion of trait-influencing variants within each section of the Venn diagram relative to the sum of all trait-influencing variants across all samples. The size of the circles reflects the polygenicity of each trait.

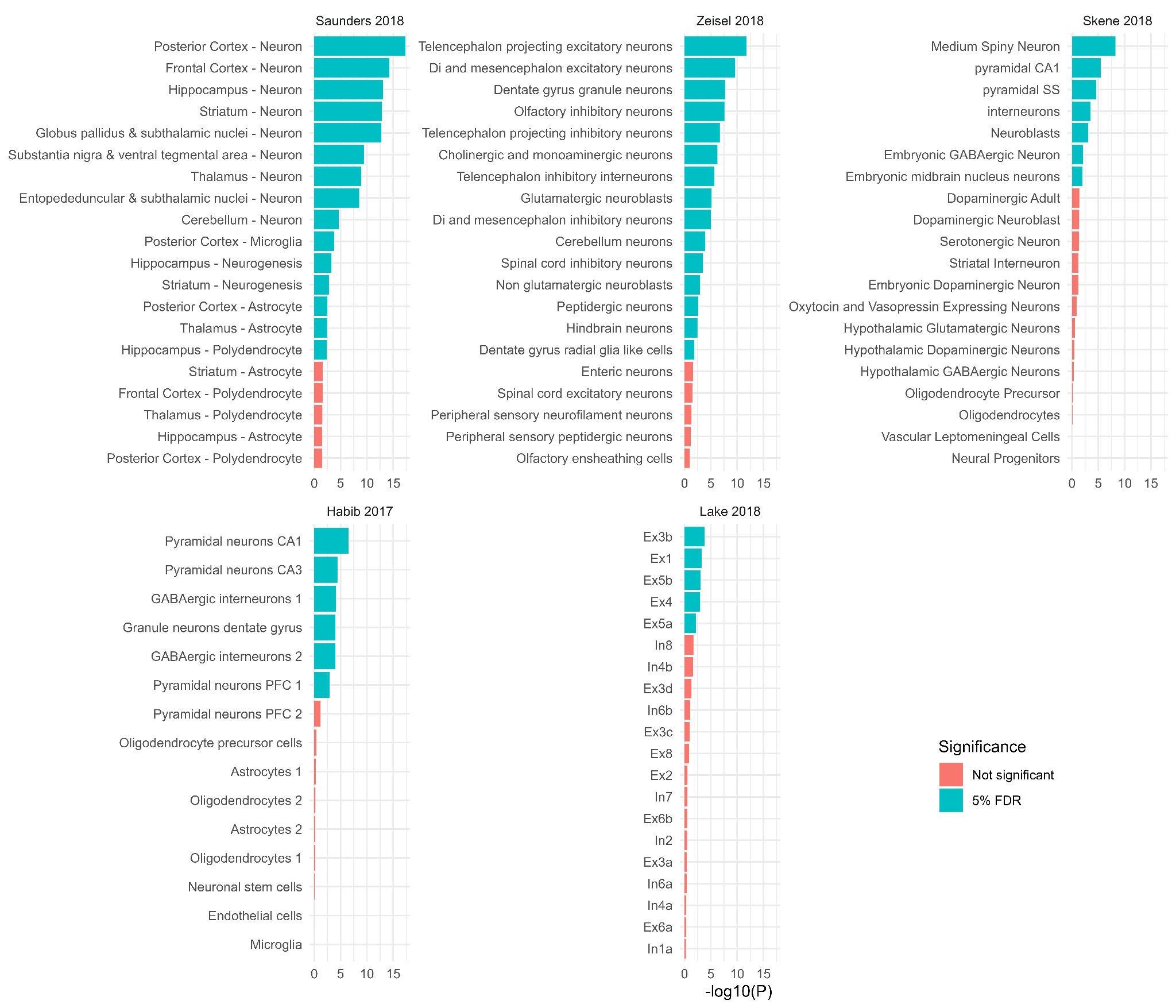

**Supplementary Figure 5. Top brain cell types enriched for bipolar disorder association signal**

The significance level (-log_10_(p value)) of MAGMA is shown for the top 20 most associated cell types in diverse datasets. The genes tested for each cell type are the top 10% of genes most specifically expressed in that cell type compared with all other cell types in the dataset. The color indicates whether the cell type is enriched for BD association signal based on the multi-ancestry meta-analysis including self-reported data at a 5% false discovery rate (FDR) within a gene expression dataset. P values are based on a linear regression and are uncorrected and one-sided. The Zeisel, Saunders and Skene datasets are derived from mouse samples, while the Habib and Lake datasets are derived from human samples. CA - CA region of the hippocampus, PFC - prefrontal cortex, SS - somato-sensory cortex, Ex - excitatory, In - inhibitory, Oli - oligodendrocyte. The numbers after the cell types refer to the cluster of cells with a similar gene expression profile, defined using clustering algorithms in the original publications.

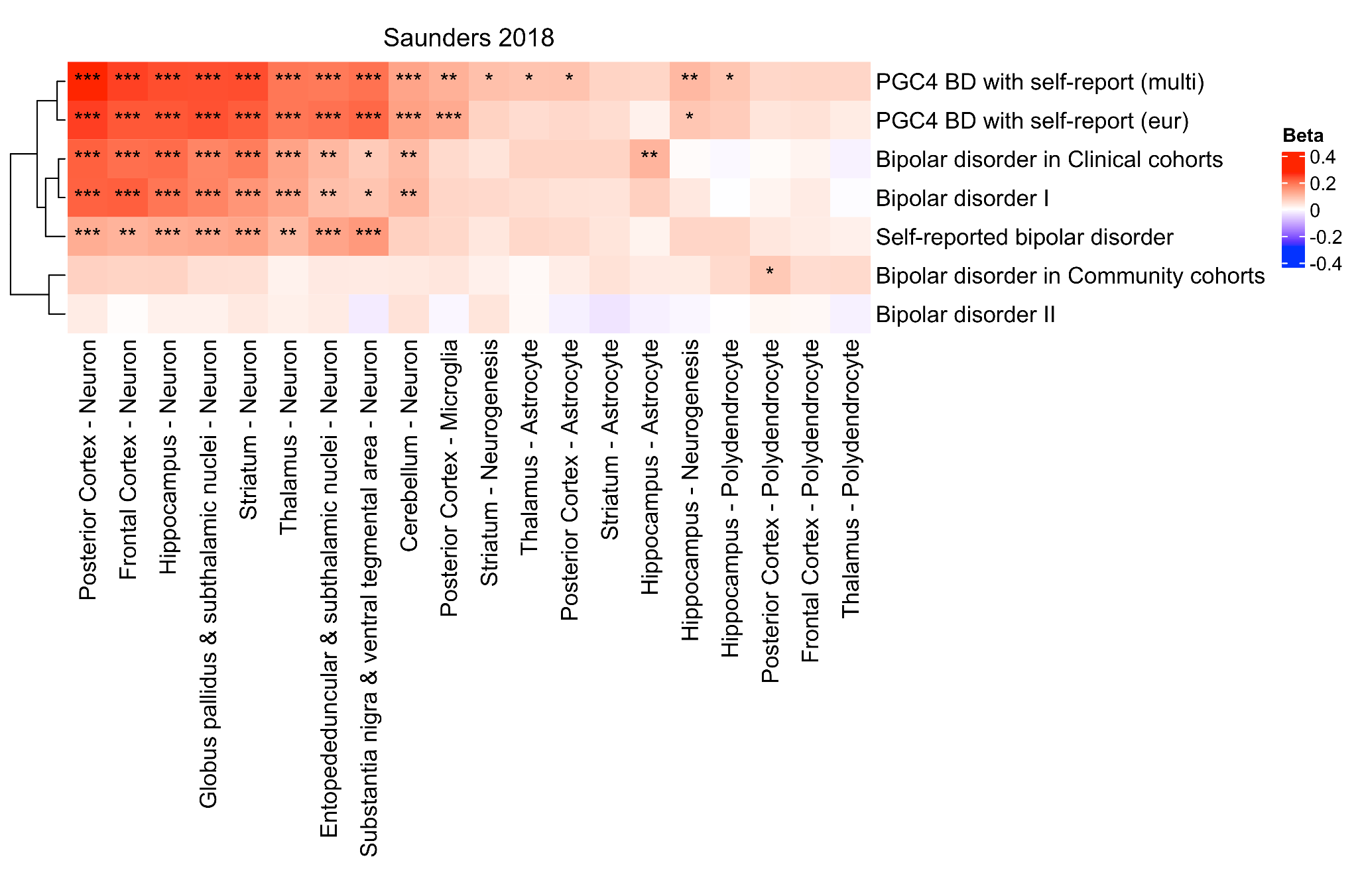

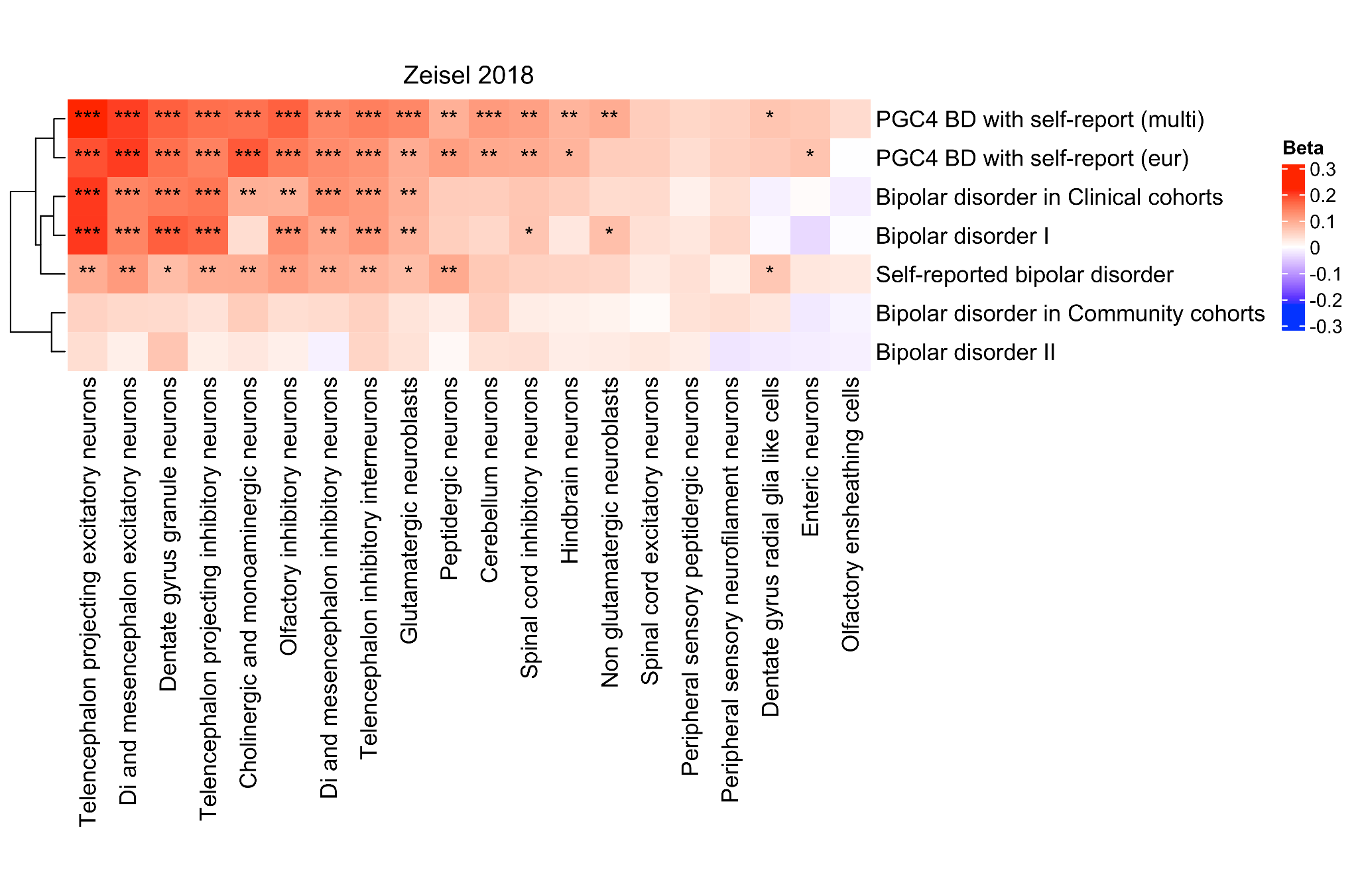

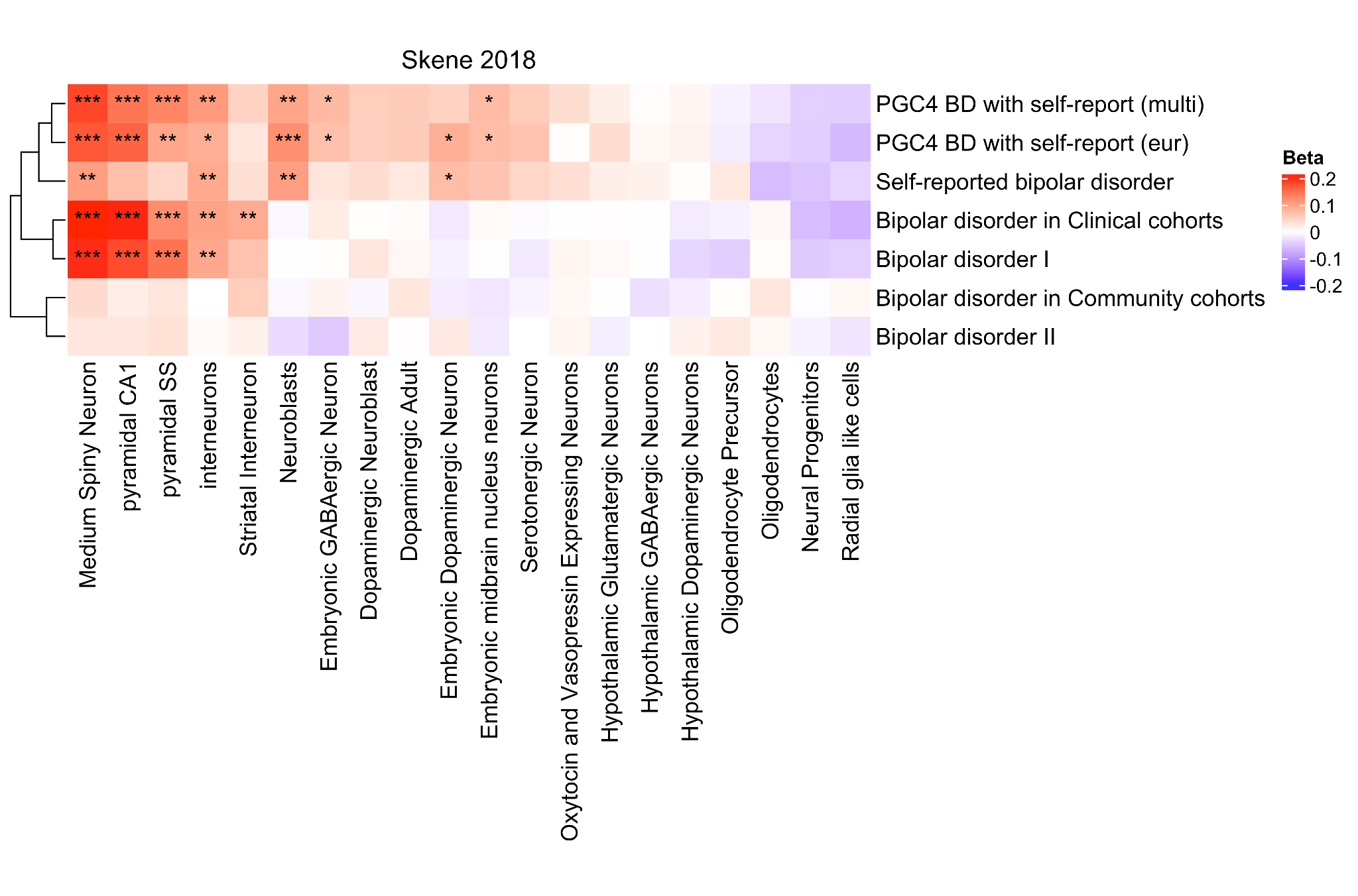

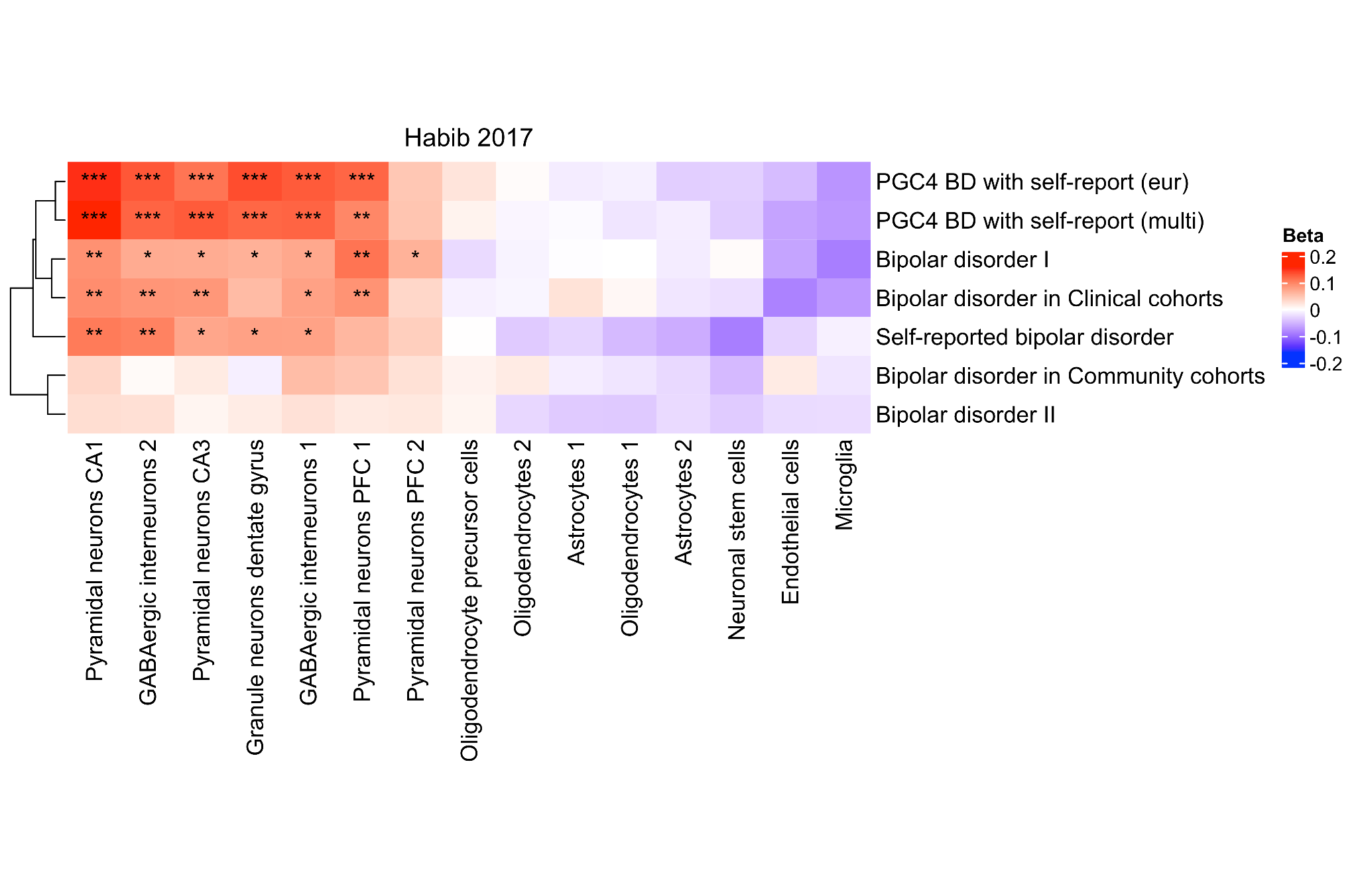

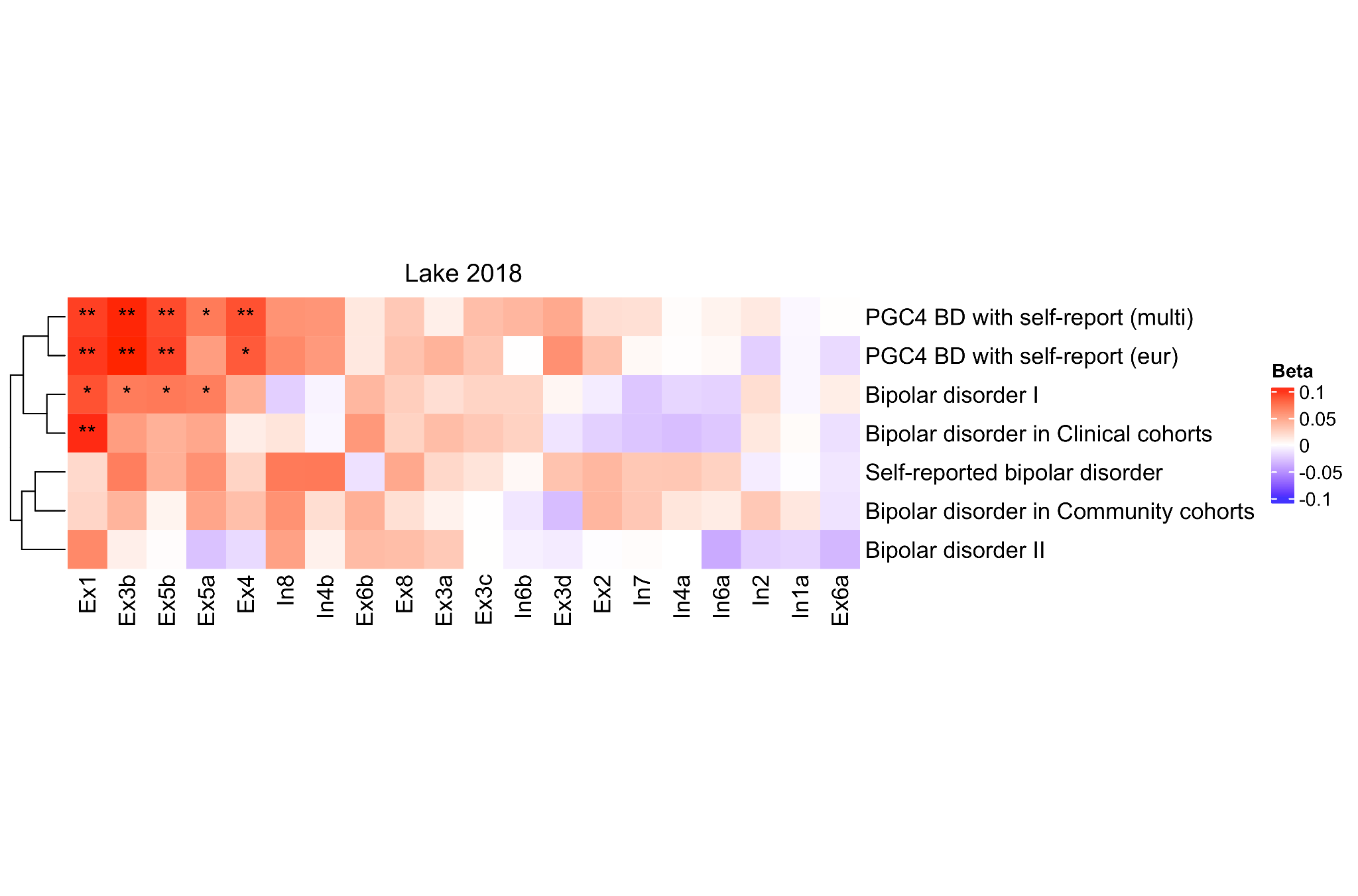

**Supplementary Figure 6. Heatmaps of the cell-type enrichment signal for bipolar disorder, including ascertainment and subtype**

The beta values (Beta) from MAGMA are shown for the top 20 cell types with lowest association p-value based on the multi-ancestry meta-analysis including self-reported data (Supplementary Figure 3). The genes tested for each cell type are the top 10% of genes most specifically expressed in that cell type compared with all other cell types in the dataset. Rows, corresponding to GWAS stratified by ascertainment and BD subtype, and columns, corresponding to cell types, were clustered based on the respective beta values. The color indicates whether the effect is positive or negative. Stars indicate the significance level of enrichment (* - FDR-adj. p < 0.05, ** - FDR-adj. p < 0.01, *** - FDR-adj. p < 0.001). The Zeisel, Saunders and Skene datasets are derived from mouse samples, while the Habib and Lake datasets are derived from human samples. CA - CA region of the hippocampus, FDR-adj. p - false discovery rate-adjusted p-value, PFC - prefrontal cortex, SS - somato-sensory cortex, Ex - excitatory, In - inhibitory, Oli - oligodendrocyte. The numbers after the cell types refer to the cluster of cells with a similar gene expression profile, defined using clustering algorithms in the original publications.

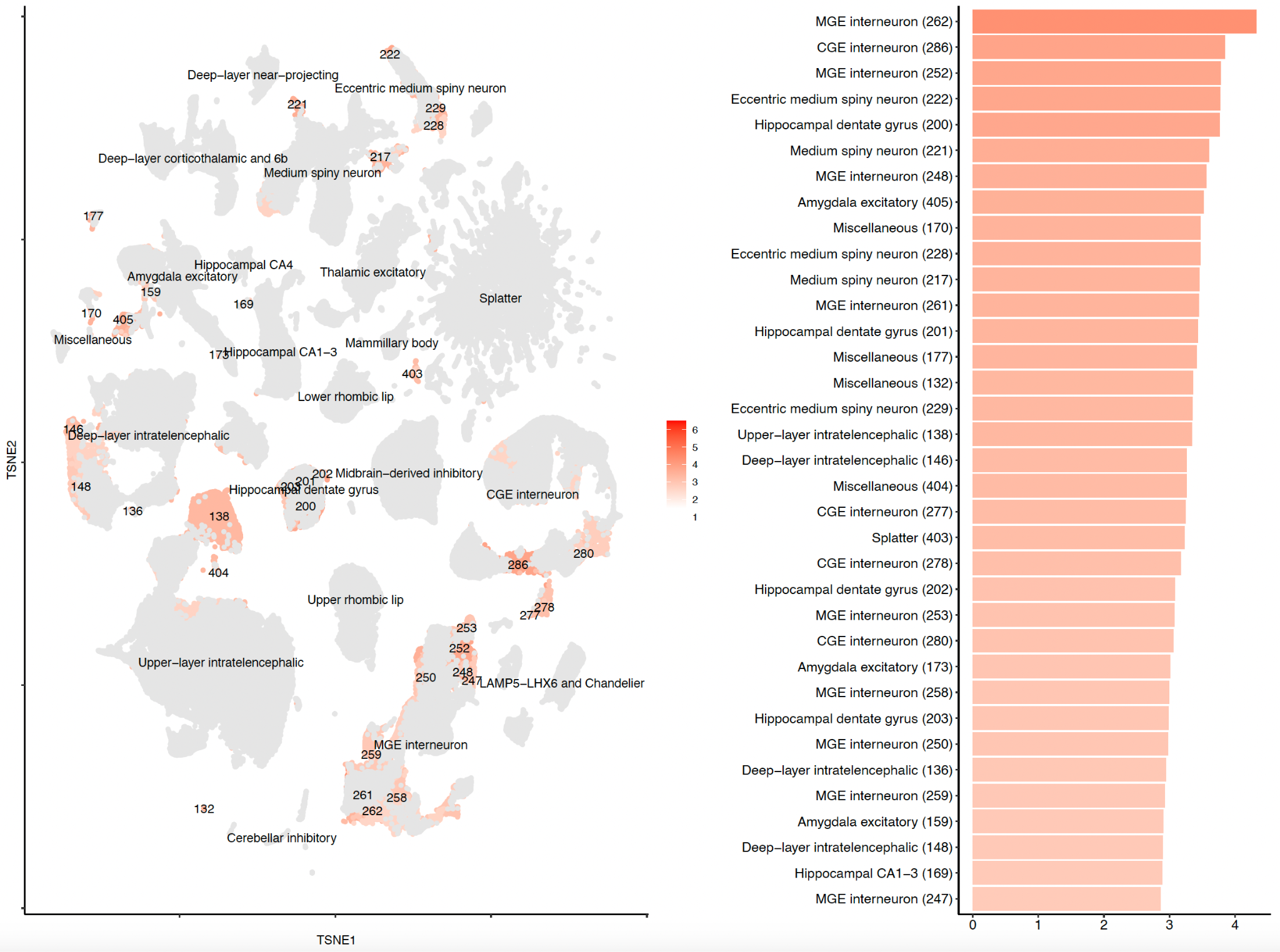

**Supplementary Figure 7. Cluster-level SNP-heritability enrichment for bipolar disorder.**

The t-distributed stochastic neighbor embedding (tSNE) plot (left) (from Siletti et al.^23^) is coloured by the enrichment z-score. Grey indicates non-significantly enriched superclusters (FDR > 0.05). The barplot (right) shows the top 35 significantly enriched clusters. The numbers in parentheses on the y-axis indicate the cell type clusters as defined in Siletti et al.^23^

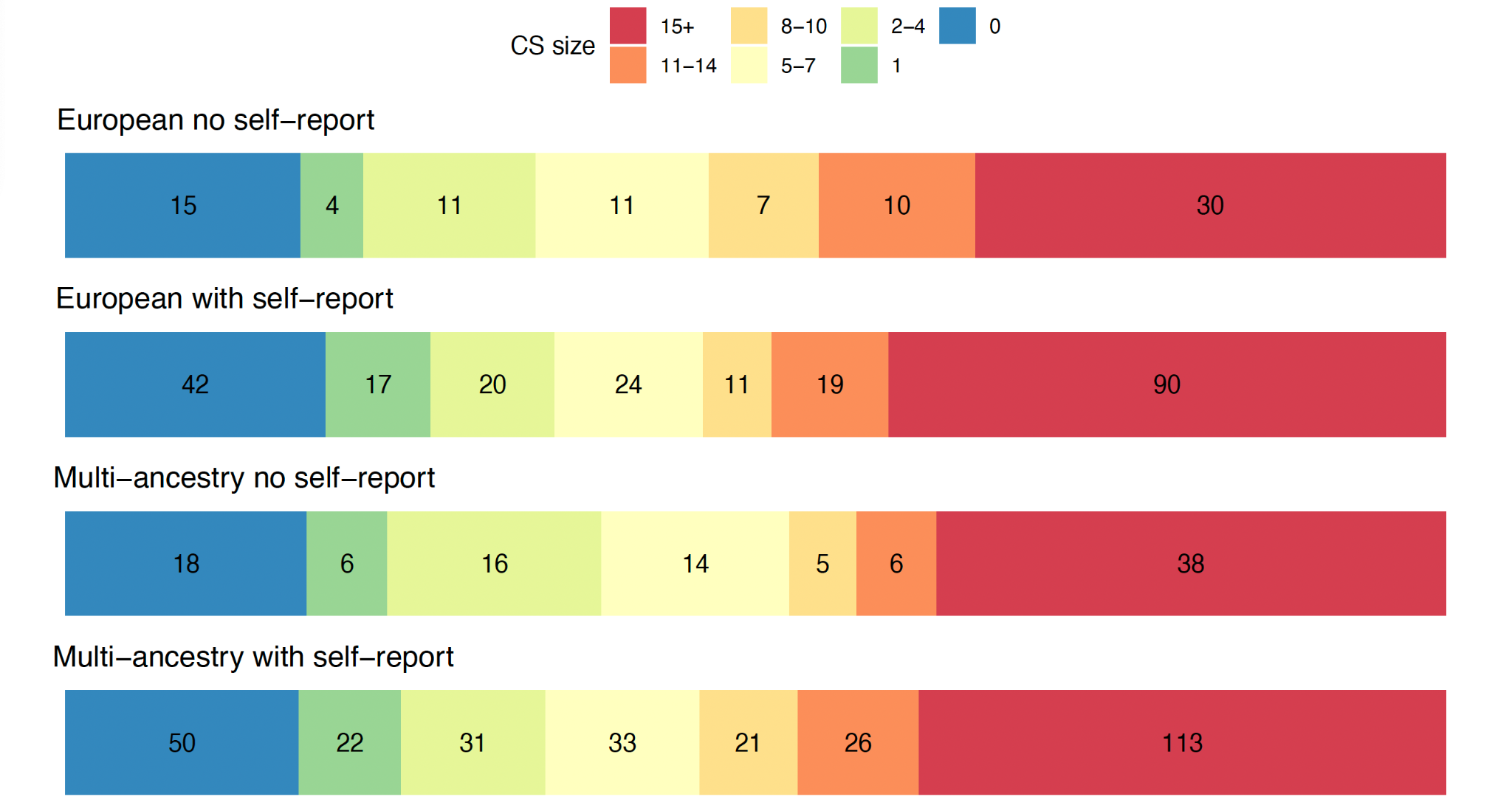

**Supplementary Figure 8. Number of SNPs within the smallest 95% credible sets (CS) from meta-analysis of European and multi-ancestry meta-analyses when excluding and including self-report data.**

Colours represent CS of varying size, with blue CS containing 0 SNPs and red CS containing 15+ SNPs. All fine-mapped SNPs regardless of their PIP were used to assess the size of the 95% credible sets.

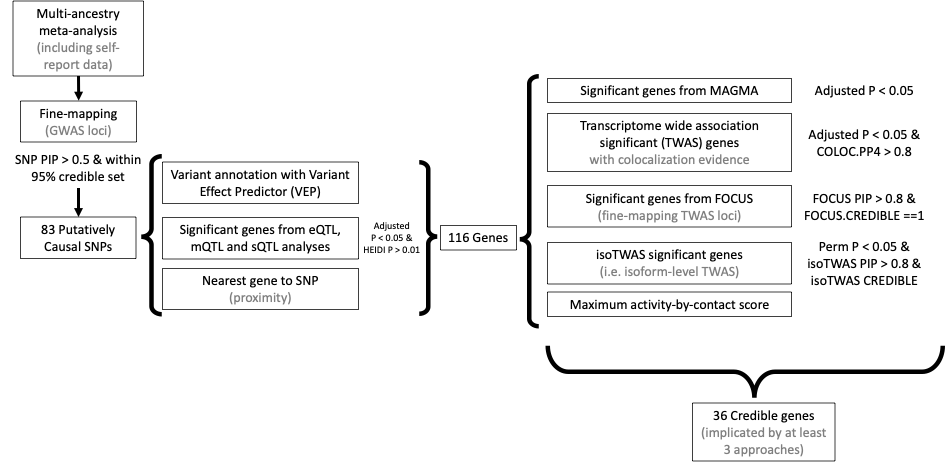

**Supplementary Figure 9. Methods and criteria for credible gene identification.**

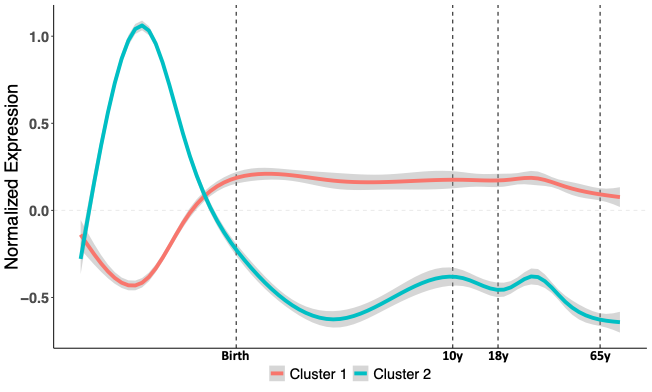

**Supplementary Figure 10. Clustering of patterns of temporal variation in expression of 34 credible genes.**

Cluster 1 shows reduced prenatal gene expression, with gene expression peaking at birth and remaining stable over the life-course. Cluster 2 includes genes with a peak gene expression during fetal development with a drop-off in expression before birth. Genes within each cluster are described in Supplementary Table 31.
