## Supplementary material for "Genomics yields biological and phenotypic insights into bipolar disorder": Author list and affiliations

Kevin S. O’Connell^1,2,*^, Maria Koromina^3,4,5,273^, Tracey van der Veen^6,273^, Toni Boltz^7,273^, Friederike S. David^8,9,273^, Jessica Mei Kay Yang^10,273^, Keng-Han Lin^11,273^, Xin Wang^11,273^, Jonathan R. I. Coleman^12,13,273^, Brittany L. Mitchell^14,15,273^, Caroline C. McGrouther^16,273^, Aaditya V. Rangan^16,17,273^, Penelope A. Lind^14,15,18,273^, Elise Koch^1,2,273^, Arvid Harder^19,273^, Nadine Parker^1,2,273^, Jaroslav Bendl^3,5,20,21,273^, Kristina Adorjan^22,23,24^, Esben Agerbo^25,26,27^, Diego Albani^28^, Silvia Alemany^29,30,31^, Ney Alliey-Rodriguez^32,33^, Thomas D. Als^25,34,35^, Till F. M. Andlauer^36^, Anastasia Antoniou^37^, Helga Ask^38,39^, Nicholas Bass^6^, Michael Bauer^40^, Eva C. Beins^8^, Tim B. Bigdeli^41,42,43,44^, Carsten Bøcker Pedersen^25,26,27^, Marco P. Boks^45^, Sigrid Børte^46,47,48^, Rosa Bosch^29,49^, Murielle Brum^50^, Ben M. Brumpton^51^, Nathalie Brunkhorst-Kanaan^50^, Monika Budde^22^, Jonas Bybjerg-Grauholm^25,52^, William Byerley^53^, Judit Cabana-Domínguez^29,30,31^, Murray J. Cairns^54,55^, Bernardo Carpiniello^56^, Miquel Casas^49,57,58^, Pablo Cervantes^59^, Chris Chatzinakos^41,43^, Hsi-Chung Chen^60,61^, Tereza Clarence^3,5,20,21^, Toni-Kim Clarke^62^, Isabelle Claus^8^, Brandon Coombes^63^, Elizabeth C. Corfield^38,64,65^, Cristiana Cruceanu^59,66^, Alfredo Cuellar-Barboza^67,68^, Piotr M. Czerski^69^, Konstantinos Dafnas^37^, Anders M. Dale^70^, Nina Dalkner^71^, Franziska Degenhardt^8,72^, J. Raymond DePaulo^73^, Srdjan Djurovic^74,75^, Ole Kristian Drange^2,76^, Valentina Escott-Price^10^, Ayman H. Fanous^77,78,79^, Frederike T. Fellendorf^71^, I. Nicol Ferrier^80^, Liz Forty^10^, Josef Frank^81^, Oleksandr Frei^1,47^, Nelson B. Freimer^82,83^, John F. Fullard^3,5,20,21^, Julie Garnham^84^, Ian R. Gizer^85^, Scott D. Gordon^86^, Katherine Gordon-Smith^87^, Tiffany A. Greenwood^88^, Jakob Grove^25,89,90,91^, José Guzman-Parra^92^, Tae Hyon Ha^93,94^, Tim Hahn^95^, Magnus Haraldsson^96,97^, Martin Hautzinger^98^, Alexandra Havdahl^38,39,64^, Urs Heilbronner^22^, Dennis Hellgren^19^, Stefan Herms^8,99,100^, Ian B. Hickie^101^, Per Hoffmann^8,99,100^, Peter A. Holmans^10^, Ming-Chyi Huang^102^, Masashi Ikeda^103,104^, Stéphane Jamain^105^, Jessica S. Johnson^3,5,106^, Lina Jonsson^107^, Janos L. Kalman^22,23^, Yoichiro Kamatani^108,109^, James L. Kennedy^110,111,112,113^, Euitae Kim^93,94,114^, Jaeyoung Kim^93,115^, Sarah Kittel-Schneider^116,117^, James A. Knowles^118^, Manolis Kogevinas^119^, Thorsten M. Kranz^50^, Kristi Krebs^120^, Steven A. Kushner^121^, Catharina Lavebratt^122,123^, Jacob Lawrence^124^, Markus Leber^125^, Heon-Jeong Lee^126^, Calwing Liao^127,128^, Susanne Lucae^129^, Martin Lundberg^122,123^, Donald J. MacIntyre^130^, Wolfgang Maier^131^, Adam X. Maihofer^88,132^, Dolores Malaspina^3,5^, Mirko Manchia^56,133^, Eirini Maratou^134^, Lina Martinsson^135,136^, Manuel Mattheisen^25,34,35,117,137^, Nathaniel W. McGregor^138^, Melvin G. McInnis^139^, James D. McKay^140^, Helena Medeiros^141^, Andreas Meyer-Lindenberg^142,143^, Vincent Millischer^122,123,144,145^, Derek W. Morris^146^, Paraskevi Moutsatsou^134^, Thomas W. Mühleisen^99,147^, Claire O’Donovan^84^, Catherine M. Olsen^148^, Georgia Panagiotaropoulou^149^, Sergi Papiol^22,23,29^, Antonio F. Pardiñas^10^, Hye Youn Park^93,94^, Amy Perry^87^, Andrea Pfennig^40^, Claudia Pisanu^150^, James B. Potash^73^, Digby Quested^151,152^, Mark H. Rapaport^153^, Eline J. Regeer^154^, John P. Rice^155^, Margarita Rivera^156,157,158^, Eva C. Schulte^8,22,131^, Fanny Senner^22,23^, Alexey Shadrin^1,2,159^, Paul D. Shilling^88^, Engilbert Sigurdsson^96,97^, Lisa Sindermann^8^, Lea Sirignano^81^, Dan Siskind^160^, Claire Slaney^84^, Laura G. Sloofman^3,5^, Olav B. Smeland^1,2^, Daniel J. Smith^161^, Janet L. Sobell^162^, Maria Soler Artigas^29,30,31,163^, Dan J. Stein^164^, Frederike Stein^9^, Mei-Hsin Su^165^, Heejong Sung^166^, Beata Świątkowska^167^, Chikashi Terao^109^, Markos Tesfaye^1,2,75^, Martin Tesli^1,2,168^, Thorgeir E. Thorgeirsson^169^, Jackson G. Thorp^14^, Claudio Toma^170,171,172^, Leonardo Tondo^173^, Paul A. Tooney^174^, Shih-Jen Tsai^175,176^, Evangelia Eirini Tsermpini^84^, Marquis P. Vawter^177^, Helmut Vedder^178^, Annabel Vreeker^45,179,180^, James T. R. Walters^10^, Bendik S. Winsvold^48,181,182^, Stephanie H. Witt^81^, Hong-Hee Won^115,183^, Robert Ye^127,128^, Allan H. Young^184,185^, Peter P. Zandi^73^, Lea Zillich^81^, 23andMe Research Team^186^, Estonian Biobank research team^186^, Genoplan Research Team^186^, HUNT All-In Psychiatry^186^, PGC-FG Single cell working group^186^, Genomic Psychiatry Cohort (GPC) Investigators^186^, Rolf Adolfsson^187^, Martin Alda^84,188^, Lars Alfredsson^189^, Lena Backlund^122,123^, Bernhard T. Baune^190,191,192^, Frank Bellivier^193,194^, Susanne Bengesser^71^, Wade H. Berrettini^195^, Joanna M. Biernacka^63,68^, Michael Boehnke^196^, Anders D. Børglum^25,89,90^, Gerome Breen^12,13^, Vaughan J. Carr^171^, Stanley Catts^197^, Sven Cichon^8,99,100,147^, Aiden Corvin^198^, Nicholas Craddock^10^, Udo Dannlowski^95^, Dimitris Dikeos^199^, Bruno Etain^193,194^, Panagiotis Ferentinos^12,37^, Mark Frye^68^, Janice M. Fullerton^170,200^, Micha Gawlik^117^, Elliot S. Gershon^32,201^, Fernando S. Goes^73^, Melissa J. Green^170,171^, Maria Grigoroiu-Serbanescu^202^, Joanna Hauser^203^, Frans A. Henskens^204^, Jens Hjerling-Leffler^205^, David M. Hougaard^25,52^, Kristian Hveem^51,206^, Nakao Iwata^104^, Ian Jones^10^, Lisa A. Jones^87^, René S. Kahn^3,45^, John R. Kelsoe^88^, Tilo Kircher^9^, George Kirov^10^, Po-Hsiu Kuo^60,207^, Mikael Landén^19,107^, Marion Leboyer^105^, Qingqin S. Li^208,209^, Jolanta Lissowska^210^, Christine Lochner^211^, Carmel Loughland^212^, Jurjen J. Luykx^213,214^, Nicholas G. Martin^86,215^, Carol A. Mathews^216^, Fermin Mayoral^92^, Susan L. McElroy^217^, Andrew M. McIntosh^130^, Francis J. McMahon^166^, Sarah E. Medland^14,218,219^, Ingrid Melle^1,220^, Lili Milani^120^, Philip B. Mitchell^171^, Gunnar Morken^221,222^, Ole Mors^25,223^, Preben Bo Mortensen^25,224^, Bertram Müller-Myhsok^129,225,226^, Richard M. Myers^227^, Woojae Myung^93,94^, Benjamin M. Neale^127,128,228^, Caroline M. Nievergelt^88,132^, Merete Nordentoft^25,229^, Markus M. Nöthen^8^, John I. Nurnberger^230^, Michael C. O’Donovan^10^, Ketil J. Oedegaard^231,232^, Tomas Olsson^233^, Michael J. Owen^10^, Sara A. Paciga^234^, Christos Pantelis^192,235,236^, Carlos N. Pato^237^, Michele T. Pato^237^, George P. Patrinos^238,239,240,241^, Joanna M. Pawlak^203^, Josep Antoni Ramos-Quiroga^29,30,31,57^, Andreas Reif^50^, Eva Z. Reininghaus^71^, Marta Ribasés^29,30,31,163^, Marcella Rietschel^81^, Stephan Ripke^127,128,149^, Guy A. Rouleau^242,243^, Panos Roussos^3,5,20,21,244^, Takeo Saito^104^, Ulrich Schall^245,246^, Martin Schalling^122,123^, Peter R. Schofield^170,200^, Thomas G. Schulze^22,73,81,247,248^, Laura J. Scott^196^, Rodney J. Scott^249,250^, Alessandro Serretti^251,252,253^, Jordan W. Smoller^128,254,255^, Alessio Squassina^150^, Eli A. Stahl^3,5,228^, Hreinn Stefansson^169^, Kari Stefansson^169,256^, Eystein Stordal^257,258^, Fabian Streit^81,142,259^, Patrick F. Sullivan^19,260,261^, Gustavo Turecki^262^, Arne E. Vaaler^263^, Eduard Vieta^264^, John B. Vincent^110^, Irwin D. Waldman^265^, Cynthia S. Weickert^170,171,266^, Thomas W. Weickert^170,171,266^, Thomas Werge^25,267,268,269^, David C. Whiteman^148^, John-Anker Zwart^47,48,181^, Howard J. Edenberg^270,271^, Andrew McQuillin^6,274^, Andreas J. Forstner^8,147,272,274^, Niamh Mullins^3,4,5,274^, Arianna Di Florio^10,261,274^, Roel A. Ophoff^7,82,83,274^, Ole A. Andreassen^1,2,274,*^, for the Bipolar Disorder Working Group of the Psychiatric Genomics Consortium

### Affiliations

^1^Division of Mental Health and Addiction, Oslo University Hospital, Oslo, Norway. ^2^NORMENT, University of Oslo, Oslo, Norway. ^3^Department of Psychiatry, Icahn School of Medicine at Mount Sinai, New York, NY, USA. ^4^Charles Bronfman Institute for Personalized Medicine, Icahn School of Medicine at Mount Sinai, New York, NY, USA. ^5^Department of Genetics and Genomic Sciences, Icahn School of Medicine at Mount Sinai, New York, NY, USA. ^6^Division of Psychiatry, University College London, London, UK. ^7^Department of Human Genetics, David Geffen School of Medicine, University of California Los Angeles, Los Angeles, CA, USA. ^8^Institute of Human Genetics, University of Bonn, School of Medicine and University Hospital Bonn, Bonn, Germany. ^9^Department of Psychiatry and Psychotherapy, University of Marburg, Marburg, Germany. ^10^Centre for Neuropsychiatric Genetics and Genomics, Division of Psychological Medicine and Clinical Neurosciences, Cardiff University, Cardiff, UK. ^11^23andMe, Inc., Sunnyvale, CA, USA. ^12^Social, Genetic and Developmental Psychiatry Centre, King’s College London, London, UK. ^13^NIHR Maudsley BRC, King’s College London, London, UK. ^14^Mental Health and Neuroscience, QIMR Berghofer Medical Research Institute, Brisbane, QLD, Australia. ^15^School of Biomedical Sciences and Faculty of Medicine, The University of Queensland, Brisbane, QLD, Australia. ^16^New York University, New York, NY, USA. ^17^Flatiron Institute, New York, NY, USA. ^18^School of Biomedical Sciences, Queensland University of Technology, Brisbane, QLD, Australia. ^19^Department of Medical Epidemiology and Biostatistics, Karolinska Institutet, Stockholm, Sweden. ^20^Center for Disease Neurogenomics, Icahn School of Medicine at Mount Sinai, New York, NY, USA. ^21^Friedman Brain Institute, Icahn School of Medicine at Mount Sinai, New York, NY, USA. ^22^Institute of Psychiatric Phenomics and Genomics (IPPG), LMU University Hospital, LMU Munich, Munich, Germany. ^23^Department of Psychiatry and Psychotherapy, University Hospital, LMU Munich, Munich, Germany. ^24^University Hospital of Psychiatry and Psychotherapy, University of Bern, Switzerland. ^25^iPSYCH, The Lundbeck Foundation Initiative for Integrative Psychiatric Research, Denmark. ^26^National Centre for Register-Based Research, Aarhus University, Aarhus, Denmark. ^27^Centre for Integrated Register-based Research, Aarhus University, Aarhus, Denmark. ^28^Department of Neuroscience, Istituto Di Ricerche Farmacologiche Mario Negri IRCCS, Milano, Italy. ^29^Instituto de Salud Carlos III, Biomedical Network Research Centre on Mental Health (CIBERSAM), Madrid, Spain. ^30^Department of Psychiatry, Hospital Universitari Vall d´Hebron, Barcelona, Spain. ^31^Psychiatric Genetics Unit, Group of Psychiatry Mental Health and Addictions, Vall d´Hebron Research Institut (VHIR), Universitat Autònoma de Barcelona, Barcelona, Spain. ^32^Department of Psychiatry and Behavioral Neuroscience, University of Chicago, Chicago, IL, USA. ^33^Northwestern University, Chicago, IL, USA. ^34^iSEQ, Center for Integrative Sequencing, Aarhus University, Aarhus, Denmark. ^35^Department of Biomedicine - Human Genetics, Aarhus University, Aarhus, Denmark. ^36^Department of Neurology, Klinikum rechts der Isar, School of Medicine, Technical University of Munich, Munich, Germany. ^37^National and Kapodistrian University of Athens, 2nd Department of Psychiatry, Attikon General Hospital, Athens, Greece. ^38^PsychGen Centre for Genetic Epidemiology and Mental Health, Norwegian Institute of Public Health, Oslo, Norway. ^39^PROMENTA Research Centre, Department of Psychology, University of Oslo, Norway. ^40^Department of Psychiatry and Psychotherapy, University Hospital Carl Gustav Carus, Technische Universität Dresden, Dresden, Germany. ^41^Department of Psychiatry and Behavioral Sciences, SUNY Downstate Health Sciences University, Brooklyn, NY, USA. ^42^VA NY Harbor Healthcare System, Brooklyn, NY, USA. ^43^Institute for Genomics in Health, SUNY Downstate Health Sciences University, Brooklyn, NY, USA. ^44^Department of Epidemiology and Biostatistics, School of Public Health, SUNY Downstate Health Sciences University, Brooklyn, NY, USA. ^45^Psychiatry, Brain Center UMC Utrecht, Utrecht, The Netherlands. ^46^Research and Communication Unit for Musculoskeletal Health, Division of Clinical Neuroscience, Oslo University Hospital, Ullevål, Oslo, Norway. ^47^Institute of Clinical Medicine, University of Oslo, Oslo, Norway. ^48^HUNT Center for Molecular and Clinical Epidemiology, Department of Public Health and Nursing, Faculty of Medicine and Health Sciences, Norwegian University of Science and Technology, Trondheim, Norway. ^49^Programa SJD MIND Escoles, Hospital Sant Joan de Déu, Institut de Recerca Sant Joan de Déu, Esplugues de Llobregat, Spain. ^50^Department of Psychiatry, Psychosomatic Medicine and Psychotherapy, University Hospital Frankfurt, Frankfurt am Main, Germany. ^51^K. G. Jebsen Center for Genetic Epidemiology, Department of Public Health and Nursing, Faculty of Medicine and Health Sciences, Norwegian University of Science and Technology, Trondheim, Norway. ^52^Center for Neonatal Screening, Department for Congenital Disorders, Statens Serum Institut, Copenhagen, Denmark. ^53^Psychiatry, University of California San Francisco, San Francisco, CA, USA. ^54^School of Biomedical Sciences and Pharmacy, The University of Newcastle, Callaghan, NSW, Australia. ^55^Precision Medicine Research Program, Hunter Medical Research Institute, New Lambton, NSW, Australia. ^56^Section of Psychiatry, Department of Medical Sciences and Public Health, University of Cagliari, Italy. ^57^Department of Psychiatry and Forensic Medicine, Universitat Autònoma de Barcelona, Barcelona, Spain. ^58^Fundació Privada d’Investigació Sant Pau (FISP), Barcelona, Spain. ^59^Department of Psychiatry, Mood Disorders Program, McGill University Health Center, Montreal, QC, Canada. ^60^Department of Psychiatry, National Taiwan University Hospital, Taipei, Taiwan. ^61^Department of Psychiatry, College of Medicine, National Taiwan University, Taipei, Taiwan. ^62^Division of Psychiatry, University of Edinburgh, Edinburgh, UK. ^63^Department of Quantitative Health Sciences Research, Mayo Clinic, Rochester, MN, USA. ^64^Nic Waals Institute, Lovisenberg Diaconal Hospital, Oslo, Norway. ^65^Department of Genetics and Bioinformatics, Norwegian Institute of Public Health, Oslo, Norway. ^66^Department of Physiology and Pharmacology, Karolinska Institutet, Stockholm, Sweden. ^67^Department of Psychiatry, Universidad Autonoma de Nuevo Leon, Monterrey, Mexico. ^68^Department of Psychiatry and Psychology, Mayo Clinic, Rochester, MN, USA. ^69^Department of Psychiatry, Laboratory of Psychiatric Genetics, Poznan University of Medical Sciences, Poznan, Poland. ^70^Center for Multimodal Imaging and Genetics, Departments of Neurosciences, Radiology, and Psychiatry, University of California, San Diego, CA, USA. ^71^Medical University of Graz, Division of Psychiatry and Psychotherapeutic Medicine, Graz, Austria. ^72^Department of Child and Adolescent Psychiatry, Psychosomatics and Psychotherapy, University Hospital Essen, University of Duisburg-Essen, Duisburg, Germany. ^73^Department of Psychiatry and Behavioral Sciences, Johns Hopkins University School of Medicine, Baltimore, MD, USA. ^74^Department of Medical Genetics, Oslo University Hospital Ullevål, Oslo, Norway. ^75^Department of Clinical Science, University of Bergen, Bergen, Norway. ^76^Department of Psychiatry, Sørlandet hospital, Kristiansand/Arendal, Norway. ^77^Department of Psychiatry, University of Arizona College of Medicine-Phoenix, Phoenix, AZ, USA. ^78^Carl T. Hayden Veterans Affairs Medical Center, Phoenix, AZ, USA. ^79^Banner-University Medical Center, Phoenix, AZ, USA. ^80^Academic Psychiatry, Newcastle University, Newcastle upon Tyne, UK. ^81^Department of Genetic Epidemiology in Psychiatry, Central Institute of Mental Health, Medical Faculty Mannheim, Heidelberg University, Mannheim, Germany. ^82^Center for Neurobehavioral Genetics, Semel Institute for Neuroscience and Human Behavior, Los Angeles, CA, USA. ^83^Department of Psychiatry and Biobehavioral Science, Semel Institute, David Geffen School of Medicine, University of California, Los Angeles, Los Angeles, CA, USA. ^84^Department of Psychiatry, Dalhousie University, Halifax, NS, Canada. ^85^Department of Psychological Sciences, University of Missouri, Columbia, MO, USA. ^86^Genetics and Computational Biology, QIMR Berghofer Medical Research Institute, Brisbane, QLD, Australia. ^87^Psychological Medicine, University of Worcester, Worcester, UK. ^88^Department of Psychiatry, University of California San Diego, La Jolla, CA, USA. ^89^Department of Biomedicine and the iSEQ Center, Aarhus University, Aarhus, Denmark. ^90^Center for Genomics and Personalized Medicine, CGPM, Aarhus, Denmark. ^91^Bioinformatics Research Centre, Aarhus University, Aarhus, Denmark. ^92^Mental Health Department, University Regional Hospital, Biomedicine Institute (IBIMA), Málaga, Spain. ^93^Department of Neuropsychiatry, Seoul National University Bundang Hospital, Seongnam, Republic of Korea. ^94^Department of Neuropsychiatry, Seoul National University College of Medicine, Seoul, Republic of Korea. ^95^Institute for Translational Psychiatry, University of Münster, Münster, Germany. ^96^Faculty of Medicine, Department of Psychiatry, School of Health Sciences, University of Iceland, Reykjavik, Iceland. ^97^Landspitali University Hospital, Reykjavik, Iceland. ^98^Department of Psychology, Eberhard Karls Universität Tübingen, Tubingen, Germany. ^99^Department of Biomedicine, University of Basel, Basel, Switzerland. ^100^Institute of Medical Genetics and Pathology, University Hospital Basel, Basel, Switzerland. ^101^Brain and Mind Centre, The University of Sydney, Sydney, NSW, Australia. ^102^Department of Psychiatry, Taipei City Psychiatric Center, Taipei City Hospital, Taipei, Taiwan. ^103^Department of Psychiatry, Nagoya University Graduate School of Medicine, Nagoya, Japan. ^104^Department of Psychiatry, Fujita Health University School of Medicine, Toyoake, Japan. ^105^Univ Paris Est Créteil, INSERM, IMRB, Translational Neuropsychiatry, Créteil, France. ^106^Department of Psychiatry, UNC Chapel Hill School of Medicine, University of North Carolina at Chapel Hill, Chapel Hill, NC, USA. ^107^Institute of Neuroscience and Physiology, University of Gothenburg, Gothenburg, Sweden. ^108^Laboratory of Complex Trait Genomics, Department of Computational Biology and Medical Sciences, Graduate School of Frontier Sciences, The University of Tokyo, Tokyo, Japan. ^109^Laboratory for Statistical and Translational Genetics, RIKEN Center for Integrative Medical Sciences, Yokohama, Japan. ^110^Campbell Family Mental Health Research Institute, Centre for Addiction and Mental Health, Toronto, ON, Canada. ^111^Neurogenetics Section, Centre for Addiction and Mental Health, Toronto, ON, Canada. ^112^Department of Psychiatry, University of Toronto, Toronto, ON, Canada. ^113^Institute of Medical Sciences, University of Toronto, Toronto, ON, Canada. ^114^Department of Brain and Cognitive Sciences, Seoul National University College of Natural Sciences, Seoul, Republic of Korea. ^115^Samsung Advanced Institute for Health Sciences and Technology (SAIHST), Sungkyunkwan University, Samsung Medical Center, Seoul, Republic of Korea. ^116^Department of Psychiatry and Neurobehavioral Science, University College Cork, Cork, Ireland. ^117^Department of Psychiatry, Psychosomatics and Psychotherapy, Center of Mental Health, University Hospital Würzburg, Würzburg, Germany. ^118^Human Genetics Institute of New Jersey, Rutgers University, Piscataway, NJ, USA. ^119^ISGlobal, Barcelona, Spain. ^120^Estonian Genome Centre, Institute of Genomics, University of Tartu, Tartu, Estonia. ^121^Department of Psychiatry, Erasmus MC, University Medical Center Rotterdam, Rotterdam, The Netherlands. ^122^Translational Psychiatry, Department of Molecular Medicine and Surgery, Karolinska Institutet, Stockholm, Sweden. ^123^Center for Molecular Medicine, Karolinska University Hospital, Stockholm, Sweden. ^124^Psychiatry, North East London NHS Foundation Trust, Ilford, UK. ^125^Clinic for Psychiatry and Psychotherapy, University Hospital Cologne, Cologne, Germany. ^126^Department of Psychiatry, Korea University College of Medicine, Seoul, Republic of Korea. ^127^Analytic and Translational Genetics Unit, Massachusetts General Hospital, Boston, MA, USA. ^128^Stanley Center for Psychiatric Research, Broad Institute, Cambridge, MA, USA. ^129^Department of Translational Research in Psychiatry, Max Planck Institute of Psychiatry, Munich, Germany. ^130^Division of Psychiatry, Centre for Clinical Brain Sciences, The University of Edinburgh, Edinburgh, UK. ^131^Department of Psychiatry and Psychotherapy, University of Bonn, School of Medicine and University Hospital Bonn, Bonn, Germany. ^132^Research/Psychiatry, Veterans Affairs San Diego Healthcare System, San Diego, CA, USA. ^133^Unit of Clinical Psychiatry, University Hospital Agency of Cagliari, Cagliari, Italy. ^134^National and Kapodistrian University of Athens, Medical School, Clinical Biochemistry Laboratory, Attikon General Hospital, Athens, Greece. ^135^Department of Clinical Neuroscience, Karolinska Institutet, Stockholm, Sweden. ^136^Centre for Psychiatry Research, SLSO Region Stockholm, Sweden. ^137^Department of Clinical Neuroscience, Centre for Psychiatry Research, Karolinska Institutet, Stockholm, Sweden. ^138^Human and Systems Genetics Working Group, Department of Genetics, Stellenbosch University, Stellenbosch, South Africa. ^139^Department of Psychiatry, University of Michigan, Ann Arbor, MI, USA. ^140^Genetic Cancer Susceptibility Group, International Agency for Research on Cancer, Lyon, France. ^141^Institute for Genomic Health, SUNY Downstate Medical Center College of Medicine, Brooklyn, NY, USA. ^142^Department of Psychiatry and Psychotherapy, Central Institute of Mental Health, Medical Faculty Mannheim, University of Heidelberg, Mannheim, Germany. ^143^German Centre for Mental Health (DZPG), Germany. ^144^Department of Psychiatry and Psychotherapy, Clinical Division of General Psychiatry, Medical University of Vienna, Austria. ^145^Comprehensive Center for Clinical Neurosciences and Mental Health, Medical University of Vienna, Vienna, Austria. ^146^Centre for Neuroimaging and Cognitive Genomics (NICOG), School of Biological and Chemical Sciences, University of Galway, Galway, Ireland. ^147^Institute of Neuroscience and Medicine (INM-1), Research Centre Jülich, Jülich, Germany. ^148^Population Health, QIMR Berghofer Medical Research Institute, Brisbane, QLD, Australia. ^149^Department of Psychiatry and Psychotherapy, Charité - Universitätsmedizin, Berlin, Germany. ^150^Department of Biomedical Sciences, University of Cagliari, Italy. ^151^Oxford Health NHS Foundation Trust, Warneford Hospital, Oxford, UK. ^152^Department of Psychiatry, University of Oxford, Warneford Hospital, Oxford, UK. ^153^Department of Psychiatry and Behavioral Sciences, Emory University School of Medicine, Atlanta, GA, USA. ^154^Outpatient Clinic for Bipolar Disorder, Altrecht, Utrecht, The Netherlands. ^155^Department of Psychiatry, Washington University in Saint Louis, Saint Louis, MO, USA. ^156^Department of Biochemistry and Molecular Biology II, Faculty of Pharmacy, University of Granada, Granada, Spain. ^157^Institute of Neurosciences ´Federico Olóriz´, Biomedical Research Center (CIBM), University of Granada, Granada, Spain. ^158^Instituto de Investigación Biosanitaria ibs.GRANADA, Granada, Spain. ^159^KG Jebsen Centre for Neurodevelopmental disorders, University of Oslo, Oslo, Norway. ^160^Faculty of Medicine, University of Queensland, Brisbane, QLD, Australia. ^161^Division of Psychiatry, Centre for Clinical Brain Sciences, University of Edinburgh, Edinburgh, UK. ^162^Psychiatry and the Behavioral Sciences, University of Southern California, Los Angeles, CA, USA. ^163^Department of Genetics, Microbiology, and Statistics, Faculty of Biology, Universitat de Barcelona, Barcelona, Spain. ^164^SAMRC Unit on Risk and Resilience in Mental Disorders, Dept of Psychiatry and Neuroscience Institute, University of Cape Town, Cape Town, South Africa. ^165^Virginia Institute for Psychiatric and Behavioral Genetics, Virginia Commonwealth University, Richmond, VA, USA. ^166^Human Genetics Branch, Intramural Research Program, National Institute of Mental Health, Bethesda, MD, USA. ^167^Department of Environmental Epidemiology, Nofer Institute of Occupational Medicine, Lodz, Poland. ^168^Department of Mental Disorders, Norwegian Institute of Public Health, Oslo, Norway. ^169^deCODE Genetics / Amgen, Reykjavik, Iceland. ^170^Neuroscience Research Australia, Sydney, NSW, Australia. ^171^Discipline of Psychiatry and Mental Health, School of Clinical Medicine, Faculty of Medicine and Health, University of New South Wales, Sydney, NSW, Australia. ^172^Centro de Biología Molecular Severo Ochoa, Universidad Autónoma de Madrid and CSIC, Madrid, Spain. ^173^Department of Psychiatry, Harvard Medical School, Boston, MA, USA. ^174^School of Biomedical Science and Pharmacy, University of Newcastle, Newcastle, NSW, Australia. ^175^Department of Psychiatry, Taipei Veterans General Hospital, Taipei, Taiwan. ^176^Division of Psychiatry, National Yang Ming Chiao Tung University, Taipei, Taiwan. ^177^Department of Psychiatry and Human Behavior, School of Medicine, University of California, Irvine, CA, USA. ^178^Psychiatry, Psychiatrisches Zentrum Nordbaden, Wiesloch, Germany. ^179^Department of Child and Adolescent Psychiatry/Psychology, Erasmus MC Sophia Children Hospital, Erasmus University, Rotterdam, The Netherlands. ^180^Department of Psychology Education and Child Studies, Erasmus School of Social and Behavioral Sciences, Erasmus University Rotterdam, The Netherlands. ^181^Department of Research, Innovation and Education, Division of Clinical Neuroscience, Oslo University Hospital, Oslo, Norway. ^182^Department of Neurology, Oslo University Hospital, Oslo, Norway. ^183^Samsung Genome Institute, Samsung Medical Center, Sungkyunkwan University School of Medicine, Seoul, Republic of Korea. ^184^Department of Psychological Medicine, Institute of Psychiatry, Psychology and Neuroscience, King’s College London, London, UK. ^185^South London and Maudsley NHS Foundation Trust, Bethlem Royal Hospital, Monks Orchard Road, Beckenham, Kent, UK. ^186^A list of members and affiliations appears in the Supplementary Note. ^187^Department of Clinical Sciences, Psychiatry, Umeå University Medical Faculty, Umeå, Sweden. ^188^National Institute of Mental Health, Klecany, Czech Republic. ^189^Institute of Environmental Medicine, Karolinska Institutet, Stockholm, Sweden. ^190^Department of Psychiatry, University of Münster, Münster, Germany. ^191^Department of Psychiatry, Melbourne Medical School, The University of Melbourne, Melbourne, VIC, Australia. ^192^The Florey Institute of Neuroscience and Mental Health, The University of Melbourne, Parkville, VIC, Australia. ^193^Université Paris Cité, INSERM, Optimisation Thérapeutique en Neuropsychopharmacologie, UMRS-1144, Paris, France. ^194^APHP Nord, DMU Neurosciences, GHU Saint Louis-Lariboisière-Fernand Widal, Département de Psychiatrie et de Médecine Addictologique, Paris, France. ^195^Psychiatry, University of Pennsylvania, Philadelphia, PA, USA. ^196^Center for Statistical Genetics and Department of Biostatistics, University of Michigan, Ann Arbor, MI, USA. ^197^University of Queensland, Brisbane, QLD, Australia. ^198^Neuropsychiatric Genetics Research Group, Dept of Psychiatry and Trinity Translational Medicine Institute, Trinity College Dublin, Dublin, Ireland. ^199^National and Kapodistrian University of Athens, 1st Department of Psychiatry, Eginition Hospital, Athens, Greece. ^200^School of Biomedical Sciences, Faculty of Medicine and Health, University of New South Wales, Sydney, NSW, Australia. ^201^Department of Human Genetics, University of Chicago, Chicago, IL, USA. ^202^Biometric Psychiatric Genetics Research Unit, Alexandru Obregia Clinical Psychiatric Hospital, Bucharest, Romania. ^203^Department of Psychiatry, Department of Psychiatric Genetics, Poznan University of Medical Sciences, Poznan, Poland. ^204^School of Medicine and Public Health, University of Newcastle, Newcastle, NSW, Australia. ^205^Department of Medical Biochemistry and Biophysics, Karolinska Institutet, Stockholm, Sweden. ^206^HUNT Research Center, Department of Public Health and Nursing, Faculty of Medicine and Health Sciences, Norwegian University of Science and Technology, Trondheim, Norway. ^207^Department of Public Health and Institute of Epidemiology and Preventive Medicine, College of Public Health, National Taiwan University, Taipei, Taiwan. ^208^Neuroscience Therapeutic Area, Janssen Research and Development, LLC, Titusville, NJ, USA. ^209^JRD Data Science, Janssen Research and Development, LLC, Titusville, NJ, USA. ^210^Cancer Epidemiology and Prevention, M. Sklodowska-Curie National Research Institute of Oncology, Warsaw, Poland. ^211^SA MRC Unit on Risk and Resilience in Mental Disorders, Dept of Psychiatry, Stellenbosch University, Stellenbosch, South Africa. ^212^University of Newcastle, Newcastle, NSW, Australia. ^213^Department of Psychiatry, Amsterdam University Medical Center, Amsterdam, The Netherlands. ^214^Department of Psychiatry and Neuropsychology, School for Mental Health and Neuroscience, Maastricht University Medical Center, Maastricht, The Netherlands. ^215^School of Psychology, The University of Queensland, Brisbane, QLD, Australia. ^216^Department of Psychiatry and Genetics Institute, University of Florida, Gainesville, FL, USA. ^217^Research Institute, Lindner Center of HOPE, Mason, OH, USA. ^218^School of Psychology and Faculty of Medicine, The University of Queensland, Brisbane, QLD, Australia. ^219^School of Psychology and Counselling, Queensland University of Technology, Brisbane, QLD, Australia. ^220^Division of Mental Health and Addiction, University of Oslo, Institute of Clinical Medicine, Oslo, Norway. ^221^Department of Mental Health, Faculty of Medicine and Health Sciences, Norwegian University of Science and Technology (NTNU), Trondheim, Norway. ^222^Psychiatry, St Olavs University Hospital, Trondheim, Norway. ^223^Psychosis Research Unit, Aarhus University Hospital - Psychiatry, Risskov, Denmark. ^224^NCRR and CIRRAU, Aarhus BSS, Aarhus University, Aarhus, Denmark. ^225^Munich Cluster for Systems Neurology (SyNergy), Munich, Germany. ^226^University of Liverpool, Liverpool, UK. ^227^HudsonAlpha Institute for Biotechnology, Huntsville, AL, USA. ^228^Medical and Population Genetics, Broad Institute, Cambridge, MA, USA. ^229^Mental Health Services in the Capital Region of Denmark, Mental Health Center Copenhagen, University of Copenhagen, Copenhagen, Denmark. ^230^Psychiatry, Indiana University School of Medicine, Indianapolis, IN, USA. ^231^Division of Psychiatry, Haukeland Universitetssjukehus, Bergen, Norway. ^232^Faculty of Medicine and Dentistry, University of Bergen, Bergen, Norway. ^233^Department of Clinical Neuroscience and Center for Molecular Medicine, Karolinska Institutet at Karolinska University Hospital, Solna, Sweden. ^234^Human Genetics and Computational Biomedicine, Pfizer Global Research and Development, Groton, CT, USA. ^235^Melbourne Neuropsychiatry Centre, Department of Psychiatry, The University of Melbourne, VIC, Australia. ^236^Monash Institute of Pharmaceutical Sciences (MIPS), Monash University, Parkville, VIC, Australia. ^237^Rutgers Health, Rutgers University, Piscataway, New Jersey, USA. ^238^University of Patras, School of Health Sciences, Department of Pharmacy, Laboratory of Pharmacogenomics and Individualized Therapy, Patras, Greece. ^239^United Arab Emirates University, College of Medicine and Health Sciences, Department of Genetics and Genomics, Al-Ain, United Arab Emirates. ^240^United Arab Emirates University, Zayed Center for Health Sciences, Al-Ain, United Arab Emirates. ^241^Erasmus University Medical Center Rotterdam, Faculty of Medicine and Health Sciences, Department of Pathology, Clinical Bioinformatics Unit, Rotterdam, The Netherlands. ^242^Department of Neurology and Neurosurgery, McGill University, Faculty of Medicine, Montreal, QC, Canada. ^243^Montreal Neurological Institute and Hospital, McGill University, Montréal, QC, Canada. ^244^Center for Precision Medicine and Translational Therapeutics, James J. Peters VA Medical Center, Bronx, NY, USA. ^245^Centre for Brain and Mental Health Research, The University of Newcastle, Newcastle, NSW, Australia. ^246^Hunter Medical Research Institute, New Lambtion Heights, NSW, Australia. ^247^Department of Psychiatry and Psychotherapy, University Medical Center Göttingen, Göttingen, Germany. ^248^Department of Psychiatry and Behavioral Sciences, SUNY Upstate Medical University, Syracuse, NY, USA. ^249^The School of Biomedical Sciences and Pharmacy, Faculty of Medicine, Health and Wellbeing, University of Newcastle, Newcastle, NSW, Australia. ^250^Cancer Detection and Therapies Program, Hunter Medical Research Institute, University of Newcastle, Newcastle, NSW, Australia. ^251^Department of Medicine and Surgery, Kore University of Enna, Enna, Italy. ^252^Department of Biomedical and Neuromotor Sciences, University of Bologna, Bologna, Italy. ^253^Oasi Research Institute-IRCCS, Troina, Italy. ^254^Department of Psychiatry, Massachusetts General Hospital, Boston, MA, USA. ^255^Psychiatric and Neurodevelopmental Genetics Unit (PNGU), Massachusetts General Hospital, Boston, MA, USA. ^256^Faculty of Medicine, University of Iceland, Reykjavik, Iceland. ^257^Department of Psychiatry, Hospital Namsos, Namsos, Norway. ^258^Department of Neuroscience, Norges Teknisk Naturvitenskapelige Universitet Fakultet for naturvitenskap og teknologi, Trondheim, Norway. ^259^Hector Institute for Artificial Intelligence in Psychiatry, Central Institute of Mental Health, Medical Faculty Mannheim, Heidelberg University, Mannheim, Germany. ^260^Department of Genetics, University of North Carolina at Chapel Hill, Chapel Hill, NC, USA. ^261^Department of Psychiatry, University of North Carolina at Chapel Hill, Chapel Hill, NC, USA. ^262^Department of Psychiatry, McGill University, Montreal, QC, Canada. ^263^Dept of Psychiatry, Sankt Olavs Hospital Universitetssykehuset i Trondheim, Trondheim, Norway. ^264^Clinical Institute of Neuroscience, Hospital Clinic, University of Barcelona, IDIBAPS, CIBERSAM, Barcelona, Spain. ^265^Department of Psychology, Emory University, Atlanta, GA, USA. ^266^Department of Neuroscience, SUNY Upstate Medical University, Syracuse, NY, USA. ^267^Institute of Biological Psychiatry, Mental Health Services, Copenhagen University Hospital, Copenhagen, Denmark. ^268^Department of Clinical Medicine, University of Copenhagen, Copenhagen, Denmark. ^269^Center for GeoGenetics, GLOBE Institute, University of Copenhagen, Copenhagen, Denmark. ^270^Biochemistry and Molecular Biology, Indiana University School of Medicine, Indianapolis, IN, USA. ^271^Department of Medical and Molecular Genetics, Indiana University, Indianapolis, IN, USA. ^272^Centre for Human Genetics, University of Marburg, Marburg, Germany. ^273^These authors contributed equally as second authors: Maria Koromina, Tracey van der Veen, Toni Boltz, Friederike S. David, Jessica Mei Kay Yang, Keng-Han Lin, Xin Wang, Jonathan R. I. Coleman, Brittany L. Mitchell, Caroline C. McGrouther, Aaditya V. Rangan, Penelope A. Lind, Elise Koch, Arvid Harder, Nadine Parker, Jaroslav Bendl. ^274^These authors jointly supervised this work: Andrew McQuillin, Andreas J. Forstner, Niamh Mullins, Arianna Di Florio, Roel A. Ophoff, Ole A. Andreassen. *Corresponding authors.
