## Supplementary Note for "Genomics yields biological and phenotypic insights into bipolar disorder"

**CONTENTS**

Supplementary Note	2

Detailed description of TWAS, FOCUS and isoTWAS eQTL analyses	2

Full acknowledgements	3

Funding Sources	9

Consortium/Group authors and affiliations	18

Competing Interests	24

### **Supplementary Note**

#### Detailed description of TWAS, FOCUS and isoTWAS eQTL analyses

As additional eQTL integration analysis, transcriptome-wide association studies (TWAS) were performed using the FUSION software^1^ with precomputed gene expression weights from the PsychENCODE dataset,^2^ available online at <http://resource.psychencode.org/>. This data is derived from 1,321 postmortem human brain samples and comprises 14,750 genes with significant *cis*-SNP heritability. Monomorphic and rare (MAF < 0.01) variants were excluded from the HRC reference panels via filtering with Plink 2^3^. We also ran colocalization^4^ tests on any gene-trait associations with a TWAS p-value less than 0.05 (--coloc_P 0.05 flag in FUSION). After excluding the 119 MHC locus genes due to complex LD structure and genes that were skipped by FUSION due to technical reasons such as an insufficient overlap between eQTLs and GWAS SNPs, TWAS p-values were Bonferroni-corrected at a p-value threshold of 3 × 10^−6^, derived by taking the nominal alpha level of 0.05 and correcting for the 14,773 genes tested. Gene-trait associations with an adjusted p-value < 0.05 were filtered for COLOC posterior probability (PP4) >= 0.8 to obtain gene-trait associations with a shared causal variant between GWAS and eQTL effect.

We performed fine-mapping of TWAS results using FOCUS^5,6^ to model the correlation among the TWAS signals and prioritise the most likely causal gene(s) in each region. For the multi-ancestry summary statistics, we used the multi-ancestry LD blocks (37:EUR-EAS-AFR). We also applied the MA-FOCUS^5,6^ software implementing the joint analysis of each of the ancestry-specific summary statistics, again using the multi-ancestry LD blocks and ancestry-matched LD reference panels from the HRC, though for the gene expression weights we used the same PsychENCODE database (EUR-specific) for all ancestries. We then kept all associations that were in the credible set and had a posterior inclusion probability (PIP) >= 0.8.

We also applied the recently developed isoform-level TWAS method (isoTWAS)^7^, using precomputed weights per isoform derived from adult PsychENCODE data, as provided by the isoTWAS developers (<https://zenodo.org/record/6795947#.Y8mi2-zMLBI>). In total we tested the 7,530 genes, each with varying numbers of isoforms, that had positive heritability with a p-value < 0.05 within the PsychENCODE data. We subset the provided PsychENOCDE SNP-isoform weights and GWAS summary statistics SNPs to those that intersect with the HRC reference panel SNPs, then perform the isoTWAS burden test to generate z-scores for each SNP-isoform pair. We then performed probabilistic fine-mapping, filtering the resulting SNP-isoform statistics to those isoforms with a burden z-test p-value < 0.05 and a permutation p-value < 0.05. The permutation p-value was generated by performing 100 random shuffles of SNP-to-isoform weights to generate a null distribution. Credible sets were defined as SNPs passing these two p-value thresholds and having a posterior inclusion probability (PIP) >= 0.8.

#### Full acknowledgements

We thank the participants who donated their time, life experiences and DNA to this research, and the clinical and scientific teams that worked with them. We are deeply indebted to the investigators who comprise the PGC. The PGC has received major funding from the US National Institute of Mental Health (PGC4: R01 MH124839; PGC3: U01 MH109528; PGC2: U01 MH094421; PGC1: U01 MH085520). Statistical analyses were carried out on the NL Genetic Cluster Computer (http://www.geneticcluster.org) hosted by SURFsara. The content is solely the responsibility of the authors and does not necessarily represent the official views of the National Institutes of Health. Niamh Mullins and Maria Koromina received funding from the Baszucki Brain Research Fund via the Milken Institute Center for Strategic Philanthropy.

**Cohort acknowledgements:**

AMQ1: Participant recruitment received support from, the Neuroscience Research Charitable Trust, the Central London NHS (National Health Service) Blood Transfusion Service, the Camden and Islington NHS Foundation Trust, a research lectureship from the Priory Hospitals, the National Institutes for Health Research (NIHR) Mental Health Research Network (MHRN), the Medical Research Council (MRC) grant number G1000708, the Stanley Medical Research Institute, and the NIHR UCLH Biomedical Research Centre (‘BRC’). Genotyping of cases and controls was performed at the Broad Institute.

GBP: Data collection was funded and data analysis was supported by the Australian National Health and Medical Research Council (No. APP1138514) to SEM. DJS is supported by a National Health and Medical Research Council Investigator Grant (No. APP1194635). NGM is supported by a National Health and Medical Research Council Investigator Grant (No. APP 1172990). SEM is supported by a National Health and Medical Research Council Investigator Grant (No. APP1172917). BLM is supported by a National Health and Medical Research Council Investigator Grant (No. APP2017176). We thank the participants for giving their time and support for this project. We acknowledge and thank M. Steffens for her generous donations and fundraising support.

BACCS: This work was supported in part by the NIHR Maudsley Biomedical Research Centre (‘BRC’) hosted at King’s College London and South London and Maudsley NHS Foundation Trust, and funded by the National Institute for Health Research under its Biomedical Research Centres funding initiative. The views expressed are those of the authors and not necessarily those of the BRC, the NHS, the NIHR or the Department of Health or King’s College London. We gratefully acknowledge capital equipment funding from the Maudsley Charity (Grant Reference 980) and Guy’s and St Thomas’s Charity (Grant Reference STR130505).

BACCS-Canada: This work was supported through funding from the Canadian Institutes of Health Research, MOP-172013, to JBV at the Centre for Addiction & Mental Health, Toronto. The ascertainment of the case control cohorts was also supported by funding from GlaxoSmithKline to JBV.

BD_TRS: This work was funded by the German Research Foundation (DFG, grant FOR2107 DA1151/5-1 to UD; SFB-TRR58, Project C09 to UD) and the Interdisciplinary Center for Clinical Research (IZKF) of the medical faculty of Münster (grant Dan3/012/17 to UD).

BiGS, GAIN: FJM and HS were supported by the NIMH Intramural Research Program, NIH, DHHS (ZIA MH002843). BOMA-Australia: This work was supported by the Australian National Health and Medical Research Council, grant numbers: 1037196 (PBM, PRS), 1177991 (PBM), 1066177 (JMF, JIN), 1063960 (JMF, PRS), 1176716 (PRS); and the Lansdowne Foundation. BOMA-Germany I, BOMA-Germany II, BOMA-Germany III, PsyCourse: This work was supported by the German Ministry for Education and Research (BMBF) through the Integrated Network IntegraMent (Integrated Understanding of Causes and Mechanisms in Mental Disorders), under the auspices of the e:Med program (grant 01ZX1314A/01ZX1614A to MMN and SC, grant 01ZX1314G/01ZX1614G to MR, grant 01ZX1314K to TGS). This work was supported by the German Ministry for Education and Research (BMBF) grants NGFNplus MooDS (Systematic Investigation of the Molecular Causes of Major Mood Disorders and Schizophrenia; grant 01GS08144 to MMN and SC, grant 01GS08147 to MR). This work was also supported by the Deutsche Forschungsgemeinschaft (DFG), grant NO246/10-1 to MMN (FOR 2107), grant RI 908/11-1 to MR (FOR 2107), grant WI 3429/3-1 to SHW, grants SCHU 1603/4-1, SCHU 1603/5-1 (KFO 241) and SCHU 1603/7-1 (PsyCourse) to TGS. This work was supported by the Swiss National Science Foundation (SNSF, grant 156791 to SC). MMN is supported through the Excellence Cluster ImmunoSensation. TGS is supported by an unrestricted grant from the Dr. Lisa-Oehler Foundation. AJF received support from the BONFOR Programme of the University of Bonn, Germany. MH was supported by the Deutsche Forschungsgemeinschaft.

FOR2107: The FOR 2107 consortium is supported by the German Research Council (Deutsche Forschungsgemeinschaft, DFG, Grant nos. KI 588/14-1, KI 588/14-2, KR 3822/7-1, KR 3822/7-2, NE 2254/1-2, DA 1151/5-1, DA 1151/5-2, SCHW 559/14-1, 545/7-2, RI 908/11-2, WI 3439/3-2, NO 246/10-2, DE 1614/3-2, HA 7070/2-2, JA 1890/7-1, JA 1890/7-2, MU 1315/8-2, RE 737/20-2, KI 588/17-1). Ethics approval was obtained from the ethics committees of the Medical Schools of the Universities of Marburg (approval identifier Studie 07/2014) and Münster, respectively, in accordance with the Declaration of Helsinki. All subjects volunteered to participate in the study and provided written informed consent.

Fran (France): This research was supported by Assistance Publique des Hôpitaux de Paris (APHP Grant PHRC GAN12), by Institut National de la Santé et de la Recherche Médicale (INSERM grant C0829), by the Fondation FondaMental and by the Investissements d’Avenir Programs managed by the Agence nationale pour la Recherche (references ANR-11-IDEX-0004-02 and ANR-10-COHO-10-01).

GREAT: This study using Taiwanese cohort was supported by the National Science and Technology Council (MOST 105-2628-B-002- 028-MY3, MOST 108-2314-B-002- 136-MY3), the National Health Research Institutes Project (NHRI-EX106-10627NI), and the National Taiwan University Career Development Project (109L7860).

Halifax: Halifax data were obtained with support from the Canadian Institutes of Health Research (grant #166098), Genome Canada, Research Nova Scotia, and from Dalhousie Medical Research Foundation.

HUNT: The Trøndelag Health Study (HUNT) is a collaboration between HUNT Research Centre (Faculty of Medicine and Health Sciences, Norwegian University of Science and Technology NTNU), Trøndelag County Council, Central Norway Regional Health Authority, and the Norwegian Institute of Public Health. The genotyping was financed by the National Institute of health (NIH), University of Michigan, The Norwegian Research council, and Central Norway Regional Health Authority and the Faculty of Medicine and Health Sciences, Norwegian University of Science and Technology (NTNU). The genotype quality control and imputation has been conducted by the K.G. Jebsen center for genetic epidemiology, Department of public health and nursing, Faculty of medicine and health sciences, Norwegian University of Science and Technology (NTNU).

IMAGE: This project was supported by grants from National Institutes of Health (grant R01MH62873) for initial sample recruitment, and from the Dutch NWO Large Investment Program (grant 1750102007010). Controls from this study collected in The Netherlands were included here.

iPSYCH BP group: ADB and the iPSYCH team acknowledges funding from The Lundbeck Foundation (R102-A9118, R155-2014-1724, and R248-2017-2003), the Stanley Medical Research Institute, NIH/NIMH (1U01MH109514-01 and 1R01MH124851-01 to A.D.B.) and the Universities and University Hospitals of Aarhus and Copenhagen. The Danish National Biobank resource was supported by the Novo Nordisk Foundation. High-performance computer capacity for handling and statistical analysis of iPSYCH data on the GenomeDK HPC facility was provided by the Center for Genomics and Personalized Medicine and the Centre for Integrative Sequencing, iSEQ, Aarhus University, Denmark (grant to A.D.B.). A.D.B. was also supported by the EU’s HORIZON-HLTH-2021-STAYHLTH-01 programme, project number 101057385: Risk and Resilience in Developmental Diversity and Mental Health ( R2D2-MH).

The analysis of Italian samples (UNICA Psychiatry Unit, UNICA Clinical pharmacology, and Lucio Bini center) was funded through the NARSAD Young investigator (2017) Brain and Behaviour foundation, the NIMH intramural research program and the ERA PerMed program (grant PLOT-BD).

Korean Cohort: This study was conducted with bioresources from National Biobank of Korea, the Korea Disease Control and Prevention Agency, Republic of Korea (KBN-2021-030).

The Mayo Bipolar Disorder Biobank was funded by the Marriot Foundation and the Mayo Clinic Center for Individualized Medicine.

Michigan (NIMH/Pritzker Neuropsychiatric Disorders Research Consortium): We thank the participants who donated their time and DNA to make this study possible. We thank members of the NIMH Human Genetics Initiative and the University of Michigan Prechter Bipolar DNA Repository for generously providing phenotype data and DNA samples. Some of the authors (L.J.S, M.B.,R.M.M) were or are members of the Pritzker Neuropsychiatric Disorders Research Consortium which is supported by the Pritzker Neuropsychiatric Disorders Research Fund L.L.C. A shared intellectual property agreement exists between this philanthropic fund and the University of Michigan, Stanford University, the Weill Medical College of Cornell University, HudsonAlpha Institute of Biotechnology, the Universities of California at Davis, and at Irvine, to encourage the development of appropriate findings for research and clinical applications. We thank Pablo Gejman for collection of the National Institute of Mental Health (NIMH) Bipolar Disorder Genetics Initiative control samples.

Data and biomaterials for bipolar cases were collected as part of 10 projects that participated in the National Institute of Mental Health (NIMH) Bipolar Disorder Genetics Initiative. From 199903, the Principal Investigators and Co-Investigators were: Indiana University, Indianapolis, IN, R01 MH59545, John Nurnberger, M.D., Ph.D., Marvin J. Miller, M.D., Elizabeth S. Bowman, M.D., N. Leela Rau, M.D., P. Ryan Moe, M.D., Nalini Samavedy, M.D., Rif El-Mallakh, M.D. (at University of Louisville), Husseini Manji, M.D. (at Wayne State University), Debra A. Glitz, M.D. (at Wayne State University), Eric T. Meyer, M.S., Carrie Smiley, R.N., Tatiana Foroud, Ph.D., Leah Flury, M.S., Danielle M. Dick, Ph.D., Howard Edenberg, Ph.D.; Washington University, St. Louis, MO, R01 MH059534, John Rice, Ph.D, Theodore Reich, M.D., Allison Goate, Ph.D., Laura Bierut, M.D.; Johns Hopkins University, Baltimore, MD, R01 MH59533, Melvin McInnis M.D., J. Raymond DePaulo, Jr., M.D., Dean F. MacKinnon, M.D., Francis M. Mondimore, M.D., James B. Potash, M.D., Peter P. Zandi, Ph.D, Dimitrios Avramopoulos, and Jennifer Payne; University of Pennsylvania, PA, R01 MH59553, Wade Berrettini M.D., Ph.D.; University of California at Irvine, CA, R01 MH60068, William Byerley M.D., and Mark Vawter M.D.; University of Iowa, IA, R01 MH059548, William Coryell M.D., and Raymond Crowe M.D.; University of Chicago, IL, R01 MH59535, Elliot Gershon, M.D., Judith Badner Ph.D., Francis McMahon M.D., Chunyu Liu Ph.D., Alan Sanders M.D., Maria Caserta, Steven Dinwiddie M.D., Tu Nguyen, Donna Harakal; University of California at San Diego, CA, R01 MH59567, John Kelsoe, M.D., Rebecca McKinney, B.A.; Rush University, IL, R01 MH059556, William Scheftner M.D., Howard M. Kravitz, D.O., M.P.H., Diana Marta, B.S., Annette Vaughn-Brown, M.S.N., R.N., and Lauri Bederow, M.A.; NIMH Intramural Research Program, Bethesda, MD, 1Z01MH002810, NCT00001174, Francis J. McMahon, M.D., Layla Kassem, PsyD., Sevilla Detera-Wadleigh, Ph.D., Dennis L. Murphy, M.D. This work was funded by NIMH, R01 MH09414501A1 (MB, RMM), NIMH, MH105653 (MB), and NIMH U01 MH085513-01 (subcontract PI, LJS).

MoBa is supported by the Norwegian Ministry of Health and Care Services and the Ministry of Education and Research. We are grateful to all the participating families in Norway who take part in this on-going cohort study. We thank the Norwegian Institute of Public Health for generating high-quality genomic data. This research is part of the HARVEST collaboration, supported by the Research Council of Norway (RCN) (#229624). We also thank the NORMENT Centre for providing genotype data, funded by the RCN (#223273), South-Eastern Norway Health Authority (SENHA) and KG Jebsen Stiftelsen. We further thank the Center for Diabetes Research, the University of Bergen for providing genotype data and performing quality control and imputation of the data funded by the ERC AdG project SELECTionPREDISPOSED, Stiftelsen Kristian Gerhard Jebsen, Trond Mohn Foundation, the RCN, the Novo Nordisk Foundation, the University of Bergen, and the Western Norway health Authorities. We thank deCODE genetics for providing genotypes for the majority of the sample. This work was performed on the TSD (Tjeneste for Sensitive Data) facilities, owned by the University of Oslo, operated and developed by the TSD service group at the University of Oslo, IT Department (USIT). The analyses were performed on resources provided by Sigma2 - the National Infrastructure for High-Performance Computing and Data Storage in Norway. Disclaimer. Data from the Norwegian Patient Registry has been used in this publication. The interpretation and reporting of these data are the sole responsibility of the authors, and no endorsement by the Norwegian Patient Registry is intended nor should be inferred.

NIH (NIMH)

Neuc1 (NeuRA-BDHR-Australia): This work was supported by the Australian National Health and Medical Research Council, grant numbers: 1037196 (PBM, PRS), 1066177 (JMF, JIN), 1063960 (JMF, PRS), 1200428 (JMF, PRS, MJG, CT), 1176716 (PRS), 1177991 (PBM); and the Lansdowne Foundation. JMF would like to thank Janette M. O'Neil and Betty C. Lynch OAM (dec) for their support. APP1200428

Neuc1 (NeuRA-CASSI-Australia): This work was funded by the NSW Ministry of Health, Office of Health and Medical Research. CSW was a recipient of National Health and Medical Research Council (Australia) Fellowships (1117079, 1021970).

Neuc1 (NeuRA-IGP-Australia): This work was supported by the Australian National Health and Medical Research Council, grant numbers: 630471 (MJG), 1081603 (MJG); MJG was supported by a NHMRC Career Development Fellowship (1061875).

Neuc1/ASGC1/ASRB: This study used samples and data from the Australian Schizophrenia Research Bank (ASRB), which is supported by the National Health and Medical Research Council of Australia (grant ID: 628386), the Pratt Foundation, Ramsay Health Care, the Viertel Charitable Foundation and the Schizophrenia Research Institute. We thank and acknowledge the contribution of the ASRB Chief Investigators: V. Carr, U. Schall, R. Scott, A. Jablensky, B. Mowry, P. Michie, S. Catts, F. Henskens and C. Pantelis.

QSkin: The QSkin Sun and Health Study (QSkin) is supported by grants from the National Health and Medical Research Council of Australia (NHMRC) (APP1185416; APP1063061; APP1073898). David C Whiteman is supported by an NHMRC Research Fellowship (APP1155413).

ROM4 (ROMANIA): The work was supported by UEFISCDI, Romania (grant number 203/2021 and the Alexandru Obregia Clinical Psychiatric Hospital. Bucharest, Romania.

Span2: This investigation was supported by Instituto de Salud Carlos III (PI18/01788, PI19/01224, PI20/00041, PI22/00464, PI23/00404, PI23/00026), and cofinanced by the European Regional Development Fund (ERDF), la Marató de TV3 (202228-30 and 202228-31), the Agència de Gestió d’Ajuts Universitaris i de Recerca-AGAUR, Generalitat de Catalunya (2021SGR-00840), the Instituto de Salud Carlos III; Instituto de Salud Carlos III and NEURON BIOPHARMA, S.A grant Nº JTC2021; Fundació la Caixa, Diputació de Barcelona, Pla Estratègic de Recerca i Innovació en Salut (PERISSLT006/17/285); Fundació Privada d'Investigació Sant Pau (FISP); and Ministry of Health of Generalitat de Catalunya. SA and MSA acknowledge their Miguel Servet contracts (CP22/00026 and CP22/00128, respectively) awarded by the Instituto de Salud Carlos III and co-funded by the European Union Found: Fondo Social Europeo Plus, FSE +.Thanks also for the support of the European Union Horizon 2020 research and innovation program (Eat2beNICE Grant Nº 728018 ; Timespan Grant Nº 965381). These samples are included in the spsp3 cohort.

SWEBIC: We are deeply grateful for the participation of all subjects contributing to this research, and to the collection team that worked to recruit them. We also wish to thank the Swedish National Quality Register for Bipolar Disorders: BipoläR. Funding support for the cohort collection was provided by the Stanley Center for Psychiatric Research, Broad Institute from a grant from Stanley Medical Research Institute. The research is further supported by grants from the Swedish Research Council (2022-01643), the Swedish foundation for Strategic Research (KF10-0039), and the Swedish Brain foundation (Hjärnfonden FO2022-0217).

Sweden: This work was funded by the Swedish Research Council (M. Schalling, C. Lavebratt,T Olsson), the Stockholm County Council (M. Schalling, C. Lavebratt, L. Backlund, L. Frisén, U. Ösby, T Olsson) and the Söderström Foundation (L. Backlund) and the Swedish Brain Foundation (T. Olsson).

UF: The University of Florida (UF) samples were collected through funding from the UF Center for OCD, Anxiety and Related Disorders (UF COARD) and genotyped through funding from the PGC.

UK - BDRN: BDRN would like to acknowledge funding from the Wellcome Trust and Stanley Medical Research Institute, and especially the research participants who continue to give their time to participate in our research. ADF and JY are funded by the European Research Council (project number 947763) and ADF is further funded by the UK Medical Research Council (grant MR/W004658/1).

UNIBO / University of Barcelona, Hospital Clinic, IDIBAPS, CIBERSAM: EV thanks the support of the Spanish Ministry of Economy and Competitiveness (PI15/00283) integrated into the Plan Nacional de I+D+I y cofinanciado por el ISCIII-Subdirección General de Evaluación y el Fondo Europeo de Desarrollo Regional (FEDER); CIBERSAM; and the Comissionat per a Universitats i Recerca del DIUE de la Generalitat de Catalunya to the Bipolar Disorders Group (2014 SGR 398).

WTCCC: The principal funder of this project was the Wellcome Trust. For the 1958 Birth Cohort, venous blood collection was funded by the UK Medical Research Council.

EstBB: This study has been supported by grants from the Estonian Research Council (grant numbers PRG184), and the CoMorMent project within the EU H2020 Framework Program (agreement no. 847776). The activities of the EstBB are regulated by the Human Genes Research Act, which was adopted in 2000 specifically for the operations of the EstBB. Individual level data analysis in the EstBB was carried out under ethical approval (1.1-12/624) from the Estonian Committee on Bioethics and Human Research (Estonian Ministry of Social Affairs). Data analysis was carried out in part in the High-Performance Computing Center of University of Tartu.

The GPC was supported by grants R01 MH085548, R01 MH104964, R01 R01MH104964, and R01MH123451 from the National Institute of Mental Health (NIMH), and genotyping of samples was provided by the Stanley Center for Psychiatric Research at Broad Institute. Funding support for the Whole Genome Association Study of Bipolar Disorder and the Genome-Wide Association of Schizophrenia Study was provided by the NIMH (R01 MH67257, R01 MH59588, R01 MH59571, R01 MH59565, R01 MH59587, R01 MH60870, R01 MH59566, R01 MH59586, R01 MH61675, R01 MH60879, R01 MH81800, U01 MH46276, U01 MH46289, U01 MH46318, U01 MH79469, and U01 MH79470) and the genotyping of samples was provided through the Genetic Association Information Network (GAIN).

This work was funded in part by a NARSAD Young Investigator award to EAS. AHY is funded by the National Institute for Health Research (NIHR) Biomedical Research Centre at South London and Maudsley NHS Foundation Trust and King’s College London. The views expressed are those of the authors and not necessarily those of the NHS, the NIHR, or the Department of Health.

This work was supported in part by the Million Veteran Program Merit Review Award #I01 BX003341 from the U.S. Department of Veterans Affairs, Biomedical Laboratory Research and Development Service and the VISN 4 Mental Illness Research, Education and Clinical Center. The views expressed in this article are those of the authors and do not necessarily represent the position or policy of the Department of Veterans Affairs or the U.S. Government.

#### Funding Sources

| **Study** | **Lead investigator** | **Country, Funder, Award number** |
| --- | --- | --- |
| PGC | PF Sullivan; EA Stahl | USA, NIMH MH109528; NIMH U01 MH109536 |
| PGC | D Posthuma | Netherlands, Scientific Organization Netherlands, 480-05-003 |
| PGC | D Posthuma | Dutch Brain Foundation and the VU University Amsterdam Netherlands |
| UK - BDRN | N Craddock  I Jones  L Jones  MJ Owen | Medical Research Council (MRC) Centre (MR/P005748/1) and Program Grants (MR/P005748/1) |
| Analysis | NR Wray | NHMRC 1078901,108788 |
| ASRB | V Carr | Australia, National Health and Medical Research Council, grant number: (86500). |
| ASRB | M Cairns | Australia, National Health and Medical Research Council, grant numbers: (1121474, 1147644). |
| ASRB | C Pantelis | Australia, National Health and Medical Research Council of Australia (grants IDs: 1196508, 1150083). |
| BACCS | G Breen | GB, JRIC, HG, CL were supported in part by the NIHR Maudsley Biomedical Research Centre (‘BRC’) hosted at King’s College London and South London and Maudsley NHS Foundation Trust, and funded by the National Institute for Health Research under its Biomedical Research Centres funding initiative. |
| BD_TRS | U Dannlowski | Germany, DFG, Grant FOR2107 DA1151/5-1; Grant SFB-TRR58, Project C09 |
| BiGS, Uchicago | ES Gershon | R01 MH103368 |
| BiGS, NIMH | FJ McMahon | US, NIMH, R01 MH061613, ZIA MH002843 |
| BiGS, GAIN, UCSD | J Kelsoe | US, NIMH, MH078151, MH081804, MH59567 |
| BOMA-Australia | JM Fullerton | Australia, National Health and Medical Research Council, grant numbers: 1066177; 1063960 |
| BOMA-Australia | SE Medland | Australia, National Health and Medical Research Council, grant numbers: 1103623 |
| BOMA-Australia | PB Mitchell | Australia, National Health and Medical Research Council, grant numbers: 1037196 and 1177991 |
| BOMA-Australia | GW Montgomery | Australia, National Health and Medical Research Council, grant numbers: 1078399 |
| BOMA-Australia | PR Schofield | Australia, National Health and Medical Research Council, grant numbers: 1037196; 1063960; 1176716 |
| ROMANIA  rom4_eur  (BOMA-Romania;  Bip_rom3_eur,  bmrom) | M Grigoroiu-Serbanescu | ROMANIA, UEFISCDI, Romania, Grant nr. 203/2021 (code PN-III-P4-ID-PCE-2020-2269) |
| BOMA-Germany I, II, III | S Cichon | Germany, BMBF Integrament, 01ZX1314A/01ZX1614A |
| BOMA-Germany I, II, III | S Cichon | Germany, BMBF NGFNplus MooDS, 01GS08144 |
| BOMA-Germany I, II, III | S Cichon | Switzerland, SNSF, 156791 |
| BOMA-Germany I, II, III | S Cichon | Switzerland, SNSF, 182731 |
| BOMA-Germany I, II, III | MM Nöthen | Germany, BMBF Integrament, 01ZX1314A/01ZX1614A |
| BOMA-Germany I, II, III | MM Nöthen | Germany, BMBF NGFNplus MooDS, 01GS08144 |
| BOMA-Germany I, II, III | MM Nöthen | Germany, Deutsche Forschungsgemeinschaft, Excellence Cluster ImmunoSensation |
| BOMA-Germany I, II, III | MM Nöthen | Germany, Deutsche Forschungsgemeinschaft, NO246/10-1 |
| BOMA-Germany I, II, III | SH Witt | Germany, Deutsche Forschungsgemeinschaft, WI 3429/3-2 |
| BOMA-Germany I, II, III, BOMA-Spain | M Rietschel | Germany, Deutsche Forschungsgemeinschaft, RI 908/11-1 |
| BOMA-Germany I, II, III, BOMA-Spain | M Rietschel | Germany, BMBF, ERA-Net Neuron “EMBED”, 01EW1904 |
| BOMA-Germany I, II, III, BOMA-Spain | M Rietschel | Germany, BMBF, ERA-Net Neuron “Synschiz”, 01EW1810 |
| BOMA-Germany I, II, III, PsyCourse, BiGS | TG Schulze | Germany, BMBF Integrament, 01ZX1314K |
| BOMA-Germany I, II, III, PsyCourse, BiGS | TG Schulze | Germany, DFG, SCHU 1603/4-1, SCHU 1603/5-1, SCHU 1603/7-1 |
| BOMA-Germany I, II, III, PsyCourse, BiGS | TG Schulze | Germany, Dr. Lisa-Oehler Foundation (Kassel, Germany) |
| Bulgarian Trios (Cardiff) | G Kirov  MJ Owen | The recruitment was funded by the Janssen Research Foundation. Genotyping was funded by multiple grants to the Stanley Center for Psychiatric Research at the Broad Institute from the Stanley Medical Research Institute, The Merck Genome Research Foundation, and the Herman Foundation. |
| EstBB | L Milani  K Krebs | Estonia, Estonian Research Council (grant numbers PRG184), European Commission EU H2020 (CoMorMent project, agreement no. 847776). |
| FOR2107 | T Kircher, U Dannlowski | German Research Foundation (Deutsche Forschungsgemeinschaft, DFG grant nos. KI 588/14-1, KI 588/14-2, KR 3822/7-1, KR 3822/7-2, NE 2254/1-2, DA 1151/5-1, DA 1151/5-2, SCHW 559/14-1, 545/7-2, RI 908/11-2, WI 3439/3-2, NO 246/10-2, DE 1614/3-2, HA 7070/2-2, JA 1890/7-1, JA 1890/7-2, MU 1315/8-2, RE 737/20-2, KI 588/17-1) |
| France | M Leboyer, F Bellivier, B Etain, S Jamain | France, APHP, INSERM, ANR, Fondation Fondamental |
| GREAT | PH Kuo | Taiwan, National Science and Technology Council, MOST 105-2628-B-002-028-MY3, MOST 108-2314-B-002- 136-MY3; National Health Research Institutes (NHRI-EX106-10627NI); National Taiwan University (Career Development Project 109L7860) |
| Halifax | M Alda | CIHR grant #166098, Research Nova Scotia, Genome Canada, and Dalhousie Medical Research Foundation |
| IMAGE | B Franke | National Institutes of Health (grant R01MH62873), Dutch NWO Large Investment Program (grant 1750102007010) |
| iPSYCH BP group | AD Børglum | Denmark, Lundbeck Foundation, R102-A9118 and R155-2014-1724, and R248-2017-2003 (iPSYCH) |
| iPSYCH BP group | AD Børglum | Denmark, Aarhus University, iSEQ, CGPM, and CIRRAU |
| iPSYCH BP group | AD Børglum | USA, NIMH 1U01MH109514-01 and 1R01MH124851-01 (sub-contract PI) |
| iPSYCH BP group | AD Børglum | EU, Horizon Europe, grant agreement no. 101057385 |
| Italy/UNICA | M Manchia, B Carpiniello | Brain and Behaviour Foundation 2017 Young investigator Award |
| Italy/Clinical Pharmacology | A Squassina, C Pisanu | National Institute of Mental Health Intramural Research Program. |
| Italy/Bini Centre | L Tondo, M Manchia | ERA PerMed Program (Grant PLOT-BD) |
| Mayo Bipolar Disorder Biobank | JM Biernacka, MA Frye | Marriot Foundation and the Mayo Clinic Center for Individualized Medicine |
| Michigan | M Boehnke, RM Myers | US, NIMH, R01 MH09414501A1 |
| Michigan | M Boehnke | US, NIMH, MH105653 |
| Michigan | L Scott | US, NIMH U01 MH085513-01 (sub-contract PI) |
| Million Veteran Program | H Kranzler | I01 BX003341 from the U.S. Department of Veterans Affairs, Biomedical Laboratory Research and Development Service and the VISN 4 Mental Illness Research, Education, and Clinical Center. |
| Mount Sinai | EA Stahl | NARSAD Young Investigator Award |
| Mount Sinai, STEP-BD, FAST | P Sklar, EA Stahl | US NIH R01MH106531, R01MH109536 |
| NeuRA-CASSI-Australia (neuc1) | C Shannon Weickert | Australia, National Health and Medical Research Council, grant number: 568807 |
| NeuRA-CASSI-Australia (neuc1) | TW Weickert | Australia, National Health and Medical Research Council, grant number: 568807 |
| NeuRA-IGP-Australia (neuc1) | MJ Green | Australia, National Health and Medical Research Council, grant numbers: 630471, 1081603 |
| NeuRA-BDHR-Australia (neuc1) | PB Mitchell, PR Schofield, JM Fullerton | Australia, National Health and Medical Research Council, grant numbers: 1037196; 1066177; 1063960, 1200428, 1176716, 1177991 & The Lansdowne Foundation |
| Norway | OA Andreassen | Norway, Research Council of Norway (#223273, #248778, #249711, #273291, #296030,#324499 #324252), KG Jebsen Stiftelsen, The South-East Norway Regional Health Authority (#2017-004, #2022-073, #2023-031)  EU’s H2020 RIA grant # 964874 REALMENT |
| Norway | KS O’Connell | US NIH 5R01MH124839-02, Research Council of Norway (#334920) |
| Norway | S Djurovic | Research Council of Norway (#223273, #248778), KG Jebsen Stiftelsen |
| Norway | OB Smeland | European Union’s Horizon 2020 Research and Innovation Programme (Grant No. 847776; CoMorMent and Grant No. 964874; RealMent) |
| Norway | E Koch | Research Council of Norway (RCN) #296030 |
| Norway | O Frei | South-Eastern Norway Regional Health Authority (#2022073); Research Council of Norway (#324499) |
| Norway | A Shadrin | Research Council of Norway (#326813); EEA and Norway grant (#EEA-RO-NO-2018-0573) |
| Norway | M Tesfaye | Research Council of Norway (RCN) (#273291, #273446) |
| Norway | I Melle | Research Council of Norway (#223273, #181831,#287714), South-Eastern Norway Regional Health Authority (#2006233, #2006258, #2011085, #2014102, #2015088, # 2021067). |
| MoBa | A Havdahl | Research Council of Norway (#274611; #336085); South-Eastern Norway Regional Health Authority (#2020022; #202283; #2019097); European Union’s Horizon 2020 Research and Innovation Programme (FAMILY; Grant No. 101057529). |
| MoBa | E.C. Corfield | Research Council of Norway (#274611); South-Eastern Norway Regional Health Authority (#2021045) |
| MoBa | H Ask | Research Council of Norway (#274611); |
| Span2 | M Ribasés, JA Ramos-Quiroga | Instituto de Salud Carlos III (PI18/01788PI19/01224, PI20/00041, PI22/00464), “la Marató de TV3” (202228-30 and 202228-31), the Agència de Gestió d’Ajuts Universitaris i de Recerca-AGAUR, Generalitat de Catalunya (2021SGR-00840), ERANA-NET Neuron Nº JTC2021,Fundació ‘la Caixa, Diputació de Barcelona, Pla Estratègic de Recerca i Innovació en Salut (PERISSLT006/17/285) and Fundació Privada d'Investigació Sant Pau(FISP), European Union Horizon 2020 (Grant Nº 728018 ; Grant Nº 965381). |
| State University of New York, Downstate Medical Center (SUNY DMC) | C Pato, MT Pato, JA Knowles, H Medeiros | US, National Institutes of Health, R01MH085542 |
| SWEBIC | M Landén | The Stanley Center for Psychiatric Research, Broad Institute from a grant from Stanley Medical Research Institute, the Swedish Research Council (2022-01643), the Swedish foundation for Strategic Research (KF10-0039), and the Swedish Brain foundation (FO2022-0217). |
| UCL | A McQuillin | Medical Research Council (MRC) - G1000708 |
| UCLA-Utrecht (Los Angeles) | RA Ophoff | US, National Institutes of Health, R01MH090553, R01MH115676 |
| UK - BDRN (Cardiff) | MC O'Donovan | Medical Research Council (MRC) Centre (MR/L010305/1 and Program Grants (MR/P005748/1) |
| UK - BDRN (Cardiff) | MJ Owen | Medical Research Council (MRC) Centre (MR/L010305/1) and Program Grants (MR/P005748/1) |
| UK - BDRN (Cardiff and Worcester) | N Craddock, I Jones, LA Jones | UK, Wellcome Trust, 078901; USA, Stanley Medical Research Institute, 5710002223-01 |
| UK - BDRN (Cardiff) | A Di Florio | European Commission Marie Curie Fellowship, grant number 623932  European Research Council; project number 947763  Medical research council (MR/W004658/1) |
| UNIBO / University of Barcelona, Hospital Clinic, IDIBAPS, CIBERSAM | E Vieta | Grants PI15/00283 (Spain) and 2021 SGR 01358 (Catalonia) |
| University of Pittsburgh | V Nimgaonkar | US, NIMH MH63480 |
| USC | JL Sobell | USA, National Institutes of Health, R01MH085542 |
| WTCCC | N Craddock; AH Young | Wellcome Trust. For the 1958 Birth Cohort, venous blood collection was funded by the UK Medical Research Council. AHY was funded by NIMH (USA); CIHR (Canada); NARSAD (USA); Stanley Medical Research Institute (USA); MRC (UK); Wellcome Trust (UK); Royal College of Physicians (Edin); BMA (UK); UBC-VGH Foundation (Canada); WEDC (Canada); CCS Depression Research Fund (Canada); MSFHR (Canada); NIHR (UK); Janssen (UK) |
| Greece | G.P. Patrinos | European Commission (H2020-668353; Ubiquitous Pharmacogenomics (U-PGx); European Commission (Horizon Europe-101057639; SafePolyMed); Greek General Secretariat of Research and Technology (MIS 5002550) |
| University of California, Irvine | MP Vawter | USA, National Institutes of Health, MH113177, MH074307, MH085801, MH099440, RR000827, MH060068, MH060870.  Pritzker Neuropsychiatric Disorders Research Fund L.L.C. |
| Korea | HJ Lee,  W Myung, H-H Won | National Research Foundation of Korea (2019R1A2C2084158 and 2017M3A9F1031220 to HJ Lee; NRF-2021R1A2C4001779 to WM;NRF-2022R1A2C2009998 to HHW). Ministry of Health & Welfare of Korea (HM14C2606) |
| BIPGRAZ, Austria | EZ Reininghaus | Funding from the government of Styria, Austria. |
| BIPGRAZ, Austria | SA Bengesser | MEFO funding of the Medical University of Graz. |
| Japan | N Iwata, M Ikeda, C Terao | Japan Agency for Medical Research and Development  (JP21wm0425008, JP21ek0109555, JP21tm0424220, JP21ck0106642, JP21wm0525024, JP23ek0410114 and JP23tm0424225)  Japan Society for the Promotion of Science (JSPS) KAKENHI grant (JP21K07490, JP22K15774, JP22H03003, JP21H02854 and JP20H00462) |
| FAST-STEP | JW Smoller | USA, NIH, R01MH063445 |
| ICCBD | JW Smoller, P Sklar | USA, NIH, R01MH085542 |
| Edinburgh, Scotland | A McIntosh | Wellcome funding, reference 220857/Z/20/Z |
| GBP | SE Medland | National Health and Medical Research Council, grant numbers: APP1138514 & APP1172917 |
| QSkin | DC Whiteman | National Health and Medical Research Council, grant numbers: APP1185416, APP1063061, & APP1073898 |

#### Consortium/Group authors and affiliations

Named authors and their affiliations appear in the main manuscript.

*HUNT All-In Psychiatry*

Amy Martinsen^1,2,14^, Anne Heidi Skogholt^2,3^, Bendik S Winsvold^2,4^, Børge Sivertsen^5,6,7^, Cristen Willer^8,9,10,^ Eystein Stordal^6,11^, Grete Dyb^12^, Gunnar Morken^6,13^, Håvard Kallestad^6,13^, Ingrid Heuch^4^, John-Anker Zwart^2,4,14^, Jonas Bille Nielsen^8,^ Katrine Kveli Fjukstad^15,16^, Lars Fritsche^8,^ Laurent Thomas^2^, Linda M Pedersen^4^, Maiken Elvestad Gabrielsen^2^, Marianne Bakke Johnsen^1,14^, Marit Skrove^17^, Marit Sæbø Indredavik^3^, Ole Kristian Drange^6,13^, Ottar Bjerkeset^6,18^, Sigrid Børte^1,2,14^, Synne Øien Stensland^1,12^, Torunn Stene Nøvik^19^, Wei Zhou^9,10^

1 Research and Communication Unit for Musculoskeletal Health (FORMI), Department of Research, Innovation and Education, Division of Clinical Neuroscience, Oslo University Hospital, Oslo, Norway.

2 K. G. Jebsen Center for Genetic Epidemiology, Department of Public Health and Nursing, Faculty of Medicine and Health Sciences, Norwegian University of Science and Technology, Trondheim, Norway.

3 Department of Clinical and Molecular Medicine, Norwegian University of Science and Technology, Trondheim, Norway.

4 Department of Research, Innovation and Education, Division of Clinical Neuroscience, Oslo University Hospital, Oslo, Norway.

5 Department of Health Promotion, Norwegian Institute of Public Health, Bergen, Norway.

6 Department of Mental Health, Faculty of Medicine and Health Sciences, Norwegian University of Science and Technology, Trondheim, Norway.

7 Department of Research and Innovation, Helse-Fonna HF, Haugesund, Norway.

8 Department of Internal Medicine, Division of Cardiovascular Medicine, University of Michigan, Ann Arbor, 48109, MI, USA.

9 Department of Computational Medicine and Bioinformatics, University of Michigan, Ann Arbor, MI, USA.

10 Department of Human Genetics, University of Michigan, Ann Arbor, MI 48109, USA; Center for Statistical Genetics, University of Michigan, Ann Arbor, MI 48109, USA.

11 Department of Psychiatry, Hospital Namsos, Nord-Trøndelag Health Trust, Namsos, Norway.

12 NKVTS, Norwegian Centre for Violence and Traumatic Stress Studies.

13 Division of Mental Health Care, St. Olavs Hospital, Trondheim University Hospital, Trondheim, Norway.

14 Institute of Clinical Medicine, Faculty of Medicine, University of Oslo, Oslo, Norway.

15 Department of Psychiatry, Nord-Trøndelag Hospital Trust, Levanger Hospital, Norway.

16 Department of Laboratory Medicine, Children’s and Women’s Health, Norwegian University of Science and Technology, Trondheim, Norway.

17 Regional Centre for Child and Youth Mental Health and Child Welfare, Department of Mental Health, Faculty of Medicine and Health Sciences, NTNU – Norwegian University of Science and Technology.

18 Faculty of Nursing and Health Sciences, NORD University, Levanger, Norway.

19 Department of Child and Adolescent Psychiatry, St. Olavs Hospital, Trondheim University Hospital, Trondheim, Norway.

Maria Grigoroiu-Serbanescu, Alexandru Obregia Clinical Psychiatric Hospital, Psychiatric Genetics Research Unit, Bucharest, Romania

*Genoplan Research Team*

Byung-Chul Lee, PhD^1,2^, Ji-Woong Kim, MS^1,2^, Young Kee Lee, MS^1,2^, Joon Ho Kang, BS^1,2^, Myeong Jae Cheon, BS^1,2^, Dong Jun Kim, MS^1,2^

^1^ Genoplan RnD Division, Genoplan Korea, Seoul, Republic of Korea

^2^ Genoplan RnD Division, Genoplan Japan, Fukuoka, Japan

*Genomic Psychiatry Cohort (GPC) Investigators*

Michele T Pato MD ^44^, Carlos N Pato, MD, PhD ^44^, Tim B Bigdeli, PhD ^1,2,3^, Ayman H Fanous, MD ^45,46,47^, Steven A McCarroll, PhD ^4,5^, Peter F Buckley, MD ^6^, Mark J. Daly ^7,8,9,5^, James A Knowles MD, PhD ^2,10^, Douglas S Lehrer, MD ^11^, Dolores Malaspina, MD, MSPH ^12,13^, Mark H Rapaport, MD ^14^, Jeffrey J Rakofsky, MD ^14^, Janet L Sobell, PhD ^15^, Giulio Genovese, PhD ^4,5^, Penelope Georgakopoulos, DrPH ^2^, Jacquelyn L Meyers, PhD ^1^, Roseann E Peterson, PhD ^6^, Helena Medeiros, MSW ^2^, Jorge Valderrama, PhD ^1,2^, Eric D Achtyes, MD ^16^, Roman Kotov, PhD ^17^, Colony Abbott, MPH ^16^, Maria Helena Azevedo, PhD ^18^, Richard A Belliveau, Jr, BA ^4^, Elizabeth Bevilacqua, BS ^19^, Evelyn J Bromet, PhD ^17^, William Byerley, MD ^20^, Celia Barreto Carvalho, PhD ^21^, Sinéad B Chapman, MS ^4^, Lynn E DeLisi, MD ^22,23^, Ashley L Dumont, BASc ^4^, Colm O'Dushlaine, PhD ^4^, Laura J Fochtmann, MD ^17^, Diane Gage ^4^, James L Kennedy, MD ^24^, Becky Kinkead, PhD ^14^, Antonio Macedo, PhD ^18^, Jennifer L Moran, PhD ^4^, Christopher P Morley, PhD ^25-27^, Mantosh J Dewan, MD ^27^, James Nemesh ^4^, Diana O Perkins, MD, MPH ^28^, Shaun M Purcell, PhD ^4,29^, Edward M Scolnick, MD ^4^, Brooke M Sklar, MA ^15^, Pamela Sklar, MD, PhD ^12,13^, Jordan W Smoller, MD, ScD ^4,23,30,31^, Patrick F Sullivan, MD, FRANZCP ^28,32^, Humberto Nicolini, MD ^33^, Conrad O Iyegbe, PhD ^34^, Fabio Macciardi, MD, PhD ^35^, Stephen R Marder, MD ^36,37^, Michael A Escamilla, MD ^38^, Ruben C Gur, PhD ^39-41^, Raquel E Gur, MD, PhD ^39-41^, Tiffany A Greenwood, PhD ^42^, David L Braff, MD ^42,43^, Marquis P Vawter, PhD, MA, MS ^35^, Chris Chatzinakos, PhD ^1,2^

^1^ Department of Psychiatry and Behavioral Sciences and ^2^ Institute for Genomics in Health, SUNY Downstate Medical Center, Brooklyn, NY, USA; ^3^ VA New York Harbor Healthcare System, Brooklyn, NY, USA; ^4^ Stanley Center for Psychiatric Research, Broad Institute of MIT and Harvard, Cambridge, MA, USA; ^5^ Department of Genetics, Harvard Medical School, Boston, MA, USA; ^6^ School of Medicine, Virginia Commonwealth University, Richmond, VA, USA; ^7^ Institute for Molecular Medicine Finland (FIMM), University of Helsinki, Helsinki, Finland; ^8^ Analytic and Translational Genetics Unit, Massachusetts General Hospital, Boston, MA, USA; ^9^ Program in Medical and Population Genetics, Broad Institute of Harvard and MIT, Cambridge, MA, USA; ^10^ Department of Cell Biology, SUNY Downstate Medical Center, Brooklyn, NY, USA; ^11^ Department of Psychiatry, Wright State University, Dayton, OH, USA; ^12^ Departments of Psychiatry and ^13^ Genetics & Genomics, Icahn School of Medicine at Mount Sinai, NY, USA; ^14^ Department of Psychiatry and Behavioral Sciences, Emory University, Atlanta, GA, USA; ^15^ Department of Psychiatry & Behavioral Sciences, University of Southern California, Los Angeles, CA, USA; ^16^ Cherry Health and Michigan State University College of Human Medicine, Grand Rapids, MI, USA; ^17^ Department of Psychiatry, Stony Brook University, Stony Brook, NY, USA; ^18^ Institute of Medical Psychology, Faculty of Medicine, University of Coimbra, Coimbra, PT; ^19^ Beacon Health Options, Boston, MA, USA; ^20^ Department of Psychiatry, University of California, San Francisco, CA, USA; ^21^ Faculty of Social and Human Sciences, University of Azores, PT; ^22^ VA Boston Healthcare System, Brockton, MA, USA; ^23^ Department of Psychiatry, Harvard Medical School, Boston, MA, USA; ^24^ Neurogenetics Laboratory, Campbell Family Mental Health Research Institute, Centre for Addiction and Mental Health; Department of Psychiatry, University of Toronto, ON, CA; ^25^ Departments of Public Health and Preventive Medicine, ^26^ Family Medicine, and ^27^ Psychiatry and Behavioral Sciences, State University of New York, Upstate Medical University, Syracuse, NY, USA; ^28^ Department of Psychiatry, University of North Carolina, Chapel Hill, NC, USA; ^29^ Department of Psychiatry, Brigham and Women’s Hospital, Boston, MA, USA; ^30^ Department of Psychiatry, Massachusetts General Hospital, Boston, MA, USA; ^31^ Department of Epidemiology, Harvard T.H. Chan School of Public Health, Boston, MA, USA; ^32^ Medical Epidemiology and Biostatistics, Karolinska Institutet, Solna, SE; ^33^ Carracci Medical Group, Mexico City, MX; ^34^ Department of Psychosis Studies, King’s College London, London, UK; ^35^ Department of Psychiatry and Human Behavior, University of California, Irvine, CA, USA; ^36^ Department of Psychiatry and Biobehavioral Sciences and ^37^ Semel Institute for Neuroscience and Human Behavior, Geffen School of Medicine, University of California Los Angeles, Los Angeles, CA, USA; ^38^ Department of Psychiatry, University of Texas Rio Grande Valley School of Medicine; ^39^ Departments of Psychiatry and ^40^ Child & Adolescent Psychiatry and ^41^ Lifespan Brain Institute, University of Pennsylvania Perelman School of Medicine and Children's Hospital of Philadelphia, Philadelphia, PA, USA; ^42^ Department of Psychiatry, University of California, La Jolla, San Diego, CA, USA; ^43^ VISN-22 Mental Illness, Research, Education and Clinical Center (MIRECC), VA San Diego Healthcare System, San Diego, CA, USA

*The CSP #572 study team*

Planning Committee: M. Aslan, M. Antonelli, M. de Asis, M. S. Bauer, M. Brophy, J. Concato, F. Cunningham, R. Freedman, M. Gaziano, T. Gleason, P. D. Harvey, G. Huang, J. Kelsoe, T. Kosten, T. Lehner, J. B. Lohr, S. R. Marder, P. Miller, T.J. O’Leary, T. Patterson, P. Peduzzi, R. Przygodzki, L. Siever, P. Sklar, S. Strakowski, H. Zhao.

Executive Committee: M. Brophy, J. Concato, A.H. Fanous, W. Farwell, M. Gaziano, P.D. Harvey, T. Kosten, A. Malhotra, S. Mane, P. Palacios, P. Sklar, L. Siever, H. Zhao.

Study Chairs’ Offices: VA Healthcare System, Bronx, NY, included L. Siever (Study Co- Chair), M. Corsey, L. Zaluda. VA Healthcare System, Miami, FL, included P.D. Harvey (Study Co- Chair), J. Johnson.

CSP Epidemiology Centers: VA Clinical Epidemiology Research Center (CERC), VA Connecticut Healthcare System, West Haven, CT, included J. Concato (Director, Methodological Co-Principal Proponent), M. Aslan, D. Cavaliere, V. Jeanpaul, A. Maffucci, L. Mancini; the Massachusetts Veterans Epidemiology Research and Information Center (MAVERIC), VA Boston Healthcare System, Jamaica Plain, MA, included M. Gaziano (Director, Methodological Co-Principal Proponent), J. Deen, G. Muldoon, S. Whitbourne.

Study Sites: *Albuquerque*: J. Canive, L. Adamson, L. Calais, G. Fuldauer, R. Kushner, G. Toney, M. Lackey, A. Mank, N. Mahdavi, G. Villarreal. *Atlanta*: E. C. Muly, F. Amin, M. Dent, J. Wold. *Baltimore*: B. Fischer, A. Elliott, C. Felix, G. Gill. *Birmingham*: P. E. Parker, C. Logan, J. McAlpine. *Brockton*: L.E. DeLisi, S. G Reece. *Charleston*: M.B. Hammer, D. Agbor-Tabie, W. Goodson. *Cincinnati*: M. Aslam, M. Grainger, Neil Richtand, Alexander Rybalsky. *Houston*: R. Al Jurdi, E. Boeckman, T. Natividad, D. Smith, M. Stewart, S. Torres, Z. Zhao. *Indianapolis*: A. Mayeda, A. Green, J. Hofstetter, S. Ngombu, M. K. Scott, A. Strasburger, J. Sumner. *Little Rock*: G. Paschall, J. Mucciarelli, R. Owen, S. Theus, D. Tompkins. *Long Beach*: S.G. Potkin, C. Reist, M. Novin, S. Khalaghizadeh. *Miami*: R. Douyon, J. Johnson, N. Kumar, B. Martinez. *Minneapolis*: S.R. Sponheim, T.L. Bender, H.L. Lucas, A.M. Lyon, M.P. Marggraf, L.H. Sorensen, C.R. Surerus. *Montrose*: C. Sison, J. Amato, D.R. Johnson, N. Pagan-Howard. *New York Harbor*: L.A. Adler, S. Alperin, T. Leon. *Northampton*: K.M. Mattocks, N. Araeva, J.C. Sullivan. *Palo Alto*: T. Suppes, K. Bratcher, L. Drag, E.G. Fischer, L. Fujitani, S. Gill, D. Grimm, J. Hoblyn, T. Nguyen, E. Nikolaev, L. Shere, R. Relova, A. Vicencio, M. Yip. *Philadelphia*: I. Hurford, S. Acheampong, G. Carfagno. *Pittsburgh*: G.L. Haas, C. Appelt, E. Brown, B. Chakraborty, E. Kelly, G. Klima, S. Steinhauer. *Salisbury*: R.A. Hurley, R. Belle, D. Eknoyan, K. Johnson, J. Lamotte. *San Diego*: E. Granholm, K. Bradshaw, J. Holden, R. H. Jones, T. Le, I.G. Molina, M. Peyton, I. Ruiz, L. Sally. *Tacoma*: A. Tapp, S. Devroy, V. Jain, N. Kilzieh, L. Maus, K. Miller, H. Pope, A. Wood. *Temple*: E. Meyer, P. Givens, P. B. Hicks, S. Justice, K. McNair, J.L. Pena, D.F. Tharp. *Tuscaloosa*: L. Davis, M. Ban, L. Cheatum, P. Darr, W. Grayson, J. Munford, D. Smith, B. Whitfield, E. Wilson. *Washington DC*: A.H. Fanous, S.E. Melnikoff, B.L Schwartz, M.A. Tureson. *West Haven*: D. D’Souza, K. Forselius, M. Ranganathan, L. Rispoli.

Albuquerque, NM, CSP Coordinating Center (for monitoring): M. Sather (Director), C. Colling, C. Haakenson, D. Krueger.

VA Office of Research and Development: T. O’Leary (Chief Research and Development Officer), G. Huang (Director, Cooperative Studies Program), T. Gleason (Director, Clinical Science Research and Development Service), R. Przygodzki (Associate Director for Genomic Medicine, and Acting Director of Biomedical Laboratory Research and Development Service), S. Muralidhar (Senior Scientific Program Manager Genomic Medicine Program, Biomedical and Clinical R&D Services).

*Million Veteran Program: Consortium Acknowledgement*

MVP Executive Committee

- Co-Chair: J. Michael Gaziano, M.D., M.P.H.

- Co-Chair: Rachel Ramoni, D.M.D., Sc.D.

- Jim Breeling, M.D. (ex-officio)

- Kyong-Mi Chang, M.D.

- Grant Huang, Ph.D.

- Sumitra Muralidhar, Ph.D.

- Christopher J. O’Donnell, M.D., M.P.H.

- Philip S. Tsao, Ph.D.

MVP Program Office

- Sumitra Muralidhar, Ph.D.

- Jennifer Moser, Ph.D.

MVP Recruitment/Enrollment

- Recruitment/Enrollment Director/Deputy Director, Boston

- Stacey B. Whitbourne, Ph.D.; Jessica V. Brewer, M.P.H.

- MVP Coordinating Centers

- Clinical Epidemiology Research Center (CERC), West Haven – John Concato, M.D., M.P.H.

- Cooperative Studies Program Clinical Research Pharmacy Coordinating Center, Albuquerque - Stuart Warren, J.D., Pharm D.; Dean P. Argyres, M.S.

- Genomics Coordinating Center, Palo Alto – Philip S. Tsao, Ph.D.

- Massachusetts Veterans Epidemiology Research Information Center (MAVERIC), Boston - J. Michael Gaziano, M.D., M.P.H.

- MVP Information Center, Canandaigua – Brady Stephens, M.S.

- Core Biorepository, Boston – Mary T. Brophy M.D., M.P.H.; Donald E. Humphries, Ph.D.

- MVP Informatics, Boston – Nhan Do, M.D.; Shahpoor Shayan

- Data Operations/Analytics, Boston – Xuan-Mai T. Nguyen, Ph.D.

MVP Science

- Genomics - Christopher J. O’Donnell, M.D., M.P.H.; Saiju Pyarajan Ph.D.; Philip S. Tsao, Ph.D.

- Phenomics - Kelly Cho, M.P.H, Ph.D.

- Data and Computational Sciences – Saiju Pyarajan, Ph.D.

- Statistical Genetics – Elizabeth Hauser, Ph.D.; Yan Sun, Ph.D.; Hongyu Zhao, Ph.D.

MVP Local Site Investigators

- Atlanta VA Medical Center (Peter Wilson) - Bay Pines VA Healthcare System (Rachel McArdle)

- Birmingham VA Medical Center (Louis Dellitalia)

- Cincinnati VA Medical Center (John Harley)

- Clement J. Zablocki VA Medical Center (Jeffrey Whittle)

- Durham VA Medical Center (Jean Beckham)

- Edith Nourse Rogers Memorial Veterans Hospital (John Wells)

- Edward Hines, Jr. VA Medical Center (Salvador Gutierrez)

- Fayetteville VA Medical Center (Gretchen Gibson)

- VA Health Care Upstate New York (Laurence Kaminsky)

- New Mexico VA Health Care System (Gerardo Villareal)

- VA Boston Healthcare System (Scott Kinlay)

- VA Western New York Healthcare System (Junzhe Xu)

- Ralph H. Johnson VA Medical Center (Mark Hamner)

- Wm. Jennings Bryan Dorn VA Medical Center (Kathlyn Sue Haddock)

- VA North Texas Health Care System (Sujata Bhushan)

- Hampton VA Medical Center (Pran Iruvanti)

- Hunter Holmes McGuire VA Medical Center (Michael Godschalk)

- Iowa City VA Health Care System (Zuhair Ballas)

- Jack C. Montgomery VA Medical Center (Malcolm Buford)

- James A. Haley Veterans’ Hospital (Stephen Mastorides)

- Louisville VA Medical Center (Jon Klein)

- Manchester VA Medical Center (Nora Ratcliffe)

- Miami VA Health Care System (Hermes Florez)

- Michael E. DeBakey VA Medical Center (Alan Swann)

- Minneapolis VA Health Care System (Maureen Murdoch)

- N. FL/S. GA Veterans Health System (Peruvemba Sriram)

- Northport VA Medical Center (Shing Shing Yeh)

- Overton Brooks VA Medical Center (Ronald Washburn)

- Philadelphia VA Medical Center (Darshana Jhala)

- Phoenix VA Health Care System (Samuel Aguayo)

- Portland VA Medical Center (David Cohen)

- Providence VA Medical Center (Satish Sharma)

- Richard Roudebush VA Medical Center (John Callaghan)

- Salem VA Medical Center (Kris Ann Oursler)

- San Francisco VA Health Care System (Mary Whooley)

- South Texas Veterans Health Care System (Sunil Ahuja)

- Southeast Louisiana Veterans Health Care System (Amparo Gutierrez)

- Southern Arizona VA Health Care System (Ronald Schifman)

- Sioux Falls VA Health Care System (Jennifer Greco)

- St. Louis VA Health Care System (Michael Rauchman)

- Syracuse VA Medical Center (Richard Servatius)

- VA Eastern Kansas Health Care System (Mary Oehlert)

- VA Greater Los Angeles Health Care System (Agnes Wallbom)

- VA Loma Linda Healthcare System (Ronald Fernando)

- VA Long Beach Healthcare System (Timothy Morgan)

- VA Maine Healthcare System (Todd Stapley)

- VA New York Harbor Healthcare System (Scott Sherman)

- VA Pacific Islands Health Care System (Gwenevere Anderson)

- VA Palo Alto Health Care System (Philip Tsao)

- VA Pittsburgh Health Care System (Elif Sonel)

- VA Puget Sound Health Care System (Edward Boyko)

- VA Salt Lake City Health Care System (Laurence Meyer)

- VA San Diego Healthcare System (Samir Gupta)

- VA Southern Nevada Healthcare System (Joseph Fayad)

- VA Tennessee Valley Healthcare System (Adriana Hung)

- Washington DC VA Medical Center (Jack Lichy)

- W.G. (Bill) Hefner VA Medical Center (Robin Hurley)

- White River Junction VA Medical Center (Brooks Robey)

- William S. Middleton Memorial Veterans Hospital (Robert Striker)

#### PGC-FG Single cell working group

#### Named authors and their affiliations appear in the main manuscript.

#### Nicholask Bray^1^, Howard Edenberg^2,3^, Mary-Ellen Lynall^4^, Mahesh Gouda^5^, Lei Hou^6^, Piotr Jaholkowski^7,8^, Gustavo Kajitani^9^, Chunyu Liu^10^, Yunlong Liu^11^, Janitza Montalvo-Ortiz^12,13,14^, Jennifer Mulle^15^, Graham Murray^16^, Nadine Parker^17^, Dalila Pinto^18,19,20^, Eva Schulte^21,22^, Erik Smedler^23,24,25^, Vladimir Vladimirov^26^,

#### Wesley Wilson^27^, Jareth Wolfe^28^, Shuyang Yao^29^,

#### Michael Ziller^30,31^.

#### 1 Centre for Neuropsychiatric Genetics & Genomics, Cardiff University, Cardiff, CF24 4HQ, UK

#### 2 Department of Medical and Molecular Genetics, School of Medicine, Indiana University, Indianapolis, IN, USA.

#### 3 Department of Biochemistry and Molecular Biology, School of Medicine, Indiana University, Indianapolis, IN, USA.

#### 4 Department of Psychiatry, Cambridge University, Herchel Smith Building, Forvie Site, Cambridge Biomedical Campus, CB2 0SZ, UK

#### 5 Department of Psychiatry and Psychotherapy, University Hospital Bonn, Medical Faculty University of Bonn, Bonn, Germany.

#### 6 Department of Medicine, Biomedical Genetics Section, Boston University School of Medicine, Boston MA, USA.

#### 7 Center for Precision Psychiatry, Division of Mental Health and Addiction, Oslo University Hospital and Institute of Clinical Medicine, University of Oslo, Oslo, Norway.

#### 8 NORMENT, Centre for Mental Disorders Research, Division of Mental Health and Addiction, Oslo University Hospital, Institute of Clinical Medicine, University of Oslo, Oslo, Norway.

#### 9 Departamento de Microbiologia, Instituto de Ciências Biomédicas, Universidade de São Paulo, São Paulo, Brazil.

#### 10 Department of Psychiatry, SUNY Upstate Medical University, Syracuse, NY 13250, USADepartment of Biostatistics, Boston University School of Public Health, Boston, MA02118, USA.

#### 11 Center for Computational Biology and Bioinformatics, Indiana University School of Medicine, Indianapolis, IN 46202, USA.

#### 12 Division of Human Genetics, Department of Psychiatry, Yale School of Medicine, New Haven, CT, USA.

#### 13 Department of Psychiatry, Yale School of Medicine, New Haven, CT, USA.

#### 14 Department of Veterans Affairs National Center for Posttraumatic Stress Disorder, Clinical Neurosciences Division, VA Connecticut Healthcare System, West Haven, CT, USA.

#### 15 Department of Psychiatry, Rutgers Robert Wood Johnson Medical School, Piscataway, NJ, USA; Center for Advanced Biotechnology and Medicine, Rutgers Robert Wood Johnson Medical School, Piscataway, NJ, USA.

#### 16 Department of Psychiatry, University of Cambridge, Cambridge, UK.

#### 17 NORMENT Centre, Division of Mental Health and Addiction, Oslo University Hospital, Institute of Clinical Medicine, University of Oslo, Building 48, Oslo, Norway.

#### 18 Department of Psychiatry, and Seaver Autism Center, Icahn School of Medicine at Mount Sinai, New York, NY, USA

#### 19 Department of Genetics and Genomic Sciences, and Icahn Genomics Institute, Icahn School of Medicine at Mount Sinai, New York, NY, USA

#### 20 The Mindich Child Health and Development Institute, Icahn School of Medicine at Mount Sinai, New York, NY, USA

#### 21 Institute of Psychiatric Phenomics and Genomics, University Hospital, Ludwig Maximilian University of Munich, Munich, Germany.

#### 22 Department of Psychiatry and Psychotherapy, University Hospital, Ludwig Maximilian University of Munich, Munich, Germany.

#### 23 Department of Psychiatry and Neurochemistry, Institute of Neuroscience and Physiology, Sahlgrenska Academy at University of Gothenburg, Gothenburg, Sweden.

#### 24 The Wallenberg Centre for Molecular and Translational Medicine, Gothenburg, Sweden.

#### 25 Department of Clinical Chemistry, Sahlgrenska University Hospital, Gothenburg, Sweden.

#### 26 University of Arizona College of Medicine, Phoenix, AZ, USA.

#### 27 University of Pennsylvania, Philadelphia, PA, USA.

#### 28 Centre for Clinical Brain Sciences, The University of Edinburgh, Edinburgh, Scotland, EH16 4SB, UK.

#### 29 Department of Medical Epidemiology and Biostatistics, Karolinska Institutet, Stockholm, Sweden.

#### 30 Department of Translational Research in Psychiatry, Max Planck Institute of Psychiatry, Munich, Germany.

#### 31 Department of Psychiatry, University of Muenster, Muenster 48149, Germany.

Estonian Biobank Research Team^1^

Andres Metspalu, Tõnu Esko, Reedik Mägi, Mari Nelis, Georgi Hudjashov

1 Estonian Genome Centre, Institute of Genomics, University of Tartu

#### Competing Interests

T.E.T., H.S. and K.S. are employed by deCODE Genetics/Amgen. E.A.S. is an employee of Regeneron Genetics Center and owns stocks of Regeneron Pharmaceutical Co. K-H.L. and X.W. are employed by 23andMe Inc.

Multiple additional authors work for pharmaceutical or biotechnology companies in a manner directly analogous to academic coauthors and collaborators.

A.H.Y. has given paid lectures and served on advisory boards relating to drugs used in affective and related disorders for several companies (AstraZeneca, Eli Lilly, Lundbeck, Sunovion, Servier, Livanova, Janssen, Allergan, Bionomics and Sumitomo Dainippon Pharma), was Lead Investigator for Embolden Study (AstraZeneca), BCI Neuroplasticity study and Aripiprazole Mania Study, and is an investigator for Janssen, Lundbeck, Livanova and Compass.

J.I.N. is an investigator for Janssen.

P.F.S. reports the following potentially competing financial interests: Neumora Therapeutics (advisory committee and shareholder).

G. Breen reports consultancy and speaker fees from Eli Lilly and Illumina and grant funding from Eli Lilly.

M. Landén has received speaker fees from Lundbeck.

O.A.A. has served as a speaker for Janssen, Lundbeck, and Sunovion and as a consultant for Cortechs.ai.

A.M.D. is a founder of and holds equity interest in CorTechs Labs and serves on its scientific advisory board; he is a member of the scientific advisory board of Human Longevity and the Mohn Medical Imaging and Visualization Center (Bergen, Norway); and he has received research funding from General Electric Healthcare.

E.V. has received grants and served as a consultant, advisor or CME speaker for the following entities: AB-Biotics, Abbott, Allergan, Angelini, AstraZeneca, Bristol Myers Squibb, Dainippon Sumitomo Pharma, Farmindustria, Ferrer, Forest Research Institute, Gedeon Richter, GlaxoSmithKline, Janssen, Lundbeck, Otsuka, Pfizer, Roche, SAGE, Sanofi-Aventis, Servier, Shire, Sunovion, Takeda, the Brain and Behaviour Foundation, the Catalan Government (AGAUR and PERIS), the Spanish Ministry of Science, Innovation, and Universities (AES and CIBERSAM), the Seventh European Framework Programme and Horizon 2020 and the Stanley Medical Research Institute.

S.K.-S. received author’s, speaker’s and consultant honoraria from Janssen, Medice Arzneimittel Pütter GmbH and Takeda outside of the current work

A.S. is or has been a consultant/speaker for: Abbott, Abbvie, Angelini, AstraZeneca, Clinical Data, Boheringer, Bristol Myers Squibb, Eli Lilly, GlaxoSmithKline, Innovapharma, Italfarmaco, Janssen, Lundbeck, Naurex, Pfizer, Polifarma, Sanofi, Servier.

J.R.D. has served as an unpaid consultant to Myriad – Neuroscience (formerly Assurex Health) in 2017 and 2019 and owns stock in CVS Health.

B.M.N. is a member of the scientific advisory board at Deep Genomics and consultant for Camp4 Therapeutics, Takeda Pharmaceutical and Biogen.

B.-C.L, J.-W.K, Y.K.L, J.H.K, M.J.C, and D.J.K are employed by Genoplan inc.

I.B.H. is the Co-Director of Health and Policy at the Brain and Mind Centre (BMC) University of Sydney. The BMC operates an early-intervention youth services at Camperdown under contract to Headspace. He is the Chief Scientific Advisor to, and a 3.2% equity shareholder in, InnoWell Pty Ltd. InnoWell was formed by the University of Sydney (45% equity) and PwC (Australia; 45% equity) to deliver the $30 M Australian Government-funded Project Synergy (2017-20; a three-year program for the transformation of mental health services) and to lead transformation of mental health services internationally through the use of innovative technologies.

MJO and MCO’D have received funding from Takeda Pharmaceuticals and Akrivia Health outside the scope of the current work. PBM has received remuneration from Janssen (Australia) and Sanofi (Hangzhou) for lectures or advisory board membership.

All other authors declare no financial interests or potential conflicts of interest.

J.A.R.Q was on the speakers’ bureau and/or acted as consultant for Biogen, Idorsia, Casen-Recordati, Janssen-Cilag, Novartis, Takeda, Bial, Sincrolab, Neuraxpharm, Novartis, BMS, Medice, Rubió, Uriach, Technofarma and Raffo in the last 3 years. He also received travel awards (air tickets + hotel) for taking part in psychiatric meetings from Idorsia, Janssen-Cilag, Rubió, Takeda, Bial and Medice. The Department of Psychiatry chaired by him received unrestricted educational and research support from the following companies in the last 3 years: Exeltis, Idorsia, Janssen- Cilag, Neuraxpharm, Oryzon, Roche, Probitas and Rubió.
